# Randomised Controlled Trial of Intravenous Nafamostat Mesylate in COVID pneumonitis: Phase 1b/2a Experimental Study to Investigate Safety, Pharmacokinetics and Pharmacodynamics

**DOI:** 10.1101/2021.10.06.21264648

**Authors:** Tom M. Quinn, Erin E. Gaughan, Annya Bruce, Jean Antonelli, Richard O’Connor, Feng Li, Sarah McNamara, Oliver Koch, Claire MacIntosh, David Dockrell, Timothy Walsh, Kevin G. Blyth, Colin Church, Jürgen Schwarze, Cecilia Boz, Asta Valanciute, Matthew Burgess, Philip Emanuel, Bethany Mills, Giulia Rinaldi, Gareth Hardisty, Ross Mills, Emily Findlay, Sunny Jabbal, Andrew Duncan, Sinéad Plant, Adam D. L. Marshall, Irene Young, Kay Russell, Emma Scholefield, Alastair F. Nimmo, Islom B. Nazarov, Grant C. Churchill, James S.O. McCullagh, Kourosh H. Ebrahimi, Colin Ferrett, Kate Templeton, Steve Rannard, Andrew Owen, Anne Moore, Keith Finlayson, Manu Shankar-Hari, John Norrie, Richard A. Parker, Ahsan R. Akram, Daniel C. Anthony, James W. Dear, Nik Hirani, Kevin Dhaliwal

**Affiliations:** Centre for Inflammation Research, Queen’s Medical Research Institute, BioQuarter, University of Edinburgh, Edinburgh, UK; Royal Infirmary of Edinburgh, BioQuarter, Little France, Edinburgh; Regional Infectious Disease Unit, NHS Lothian; Institute of Cancer Sciences, University of Glasgow; Department of Respiratory Medicine, Queen Elizabeth University Hospital, NHS Greater Glasgow and Clyde Health Board, Glasgow, UK; Latus Therapeutics, Oxford, UK; Department of Pharmacology, University of Oxford, Oxford, UK; Department of Radiology, Oxford University Hospitals NHS Foundation Trust, Oxford, UK; Department of Chemistry, University of Oxford, Oxford, UK; Centre of Excellence for Long-acting Therapeutics, Materials Innovation Factory & Department of Pharmacology and Therapeutics, University of Liverpool; Edinburgh Clinical Trials Unit (ECTU), Usher Institute, University of Edinburgh, Edinburgh, UK; Centre for Cardiovascular Science, Queen’s Medical Research Institute, Bioquarter, University of Edinburgh, Edinburgh, UK; Institute of Pharmaceutical Science, King’s College London, UK

## Abstract

Despite the success of vaccines and selected repurposed treatments, COVID-19 is likely to remain a global health problem and further chemotherapeutics are required. Many repurposed drugs have progressed rapidly to Phase 2 and 3 trials without characterisation of Pharmacokinetics (PK)/Pharmacodynamics (PD) including safety in COVID-19. One such drug is Nafamostat Mesylate (Nafamostat), a synthetic serine protease inhibitor with anticoagulant and anti-inflammatory properties. Preclinical data has demonstrated that it is has potent antiviral activity against SARS-CoV-2 by directly inhibiting the transmembrane protease serine 2 (TMPRSS2) dependent stage of host cell entry.

**Methods:** We present the findings of a phase Ib/II open label, platform randomised controlled trial (RCT), exploring the safety of intravenous Nafamostat in hospitalised patients with confirmed COVID-19 pneumonitis. Patients were assigned randomly to standard of care (SoC), Nafamostat or an alternative therapy. Secondary endpoints included clinical endpoints such as number of oxygen free days and clinical improvement/ deterioration, PK/PD, thromboelastometry, D Dimers, cytokines, immune cell flow cytometry and viral load.

**Results:** Data is reported from 42 patients, 21 of which were randomly assigned to receive intravenous Nafamostat. The Nafamostat group developed significantly higher plasma creatinine levels, more adverse events and a lower number of oxygen free days. There were no other statistically significant differences in the primary or secondary endpoints between Nafamostat and SoC. PK data demonstrated that intravenous Nafamostat was rapidly broken down to inactive metabolites. We observed an antifibrinolytic profile, and no significant anticoagulant effects in thromboelastometry. Participants in the Nafamostat group had higher D Dimers compared to SoC. There were no differences in cytokine profile and immune cell phenotype and viral loads between the groups.

**Conclusion:** In hospitalised patients with COVID-19, we did not observe evidence of anti-inflammatory, anticoagulant or antiviral activity with intravenous Nafamostat. Given the number of negative trials with repurposed drugs, our experimental medicine trial highlights the value of PK/PD studies prior to selecting drugs for efficacy trials. Given the mechanism of action, further evaluation of Nafamostat delivered via a different route may be warranted. This trial demonstrates the importance of experimental trials in new disease entities such as COVID-19 prior to selecting drugs for larger trials.

## INTRODUCTION

COVID-19, caused by the coronavirus SARS-CoV-2, was declared a global pandemic on the 11^th^ of March 2020 [1] and an ongoing global health, social and economic crisis has ensued. Vaccination programmes are at varying stages globally, with concerns in vaccinated populations regarding resistant strains ever-present. Identifying effective treatments for preventing clinical deterioration is therefore of paramount importance. At the time of writing, dexamethasone and interleukin-6 receptor antagonists [2] are the only effective treatments available for COVID-19 [3] [4], however, the mortality rate of unvaccinated COVID-19 in hospitalised patients remains high at 22.9% [2].

Further chemotherapeutics are therefore required, with the repurposing of pre-existing drugs, quicker and more cost-effective than the development of new medications.

Nafamostat Mesylate (Nafamostat) is a synthetic protease inhibitor and directly inhibits the transmembrane protease serine 2 (TMPRSS2) dependent stage of host cell entry of MERS-CoV, therefore, blocking human cell entry [5]. This method of cell entry is shared by other coronaviruses including SARS-CoV-2, and *in-vitro* studies have confirmed activity against SARS-CoV-2 [6, 7]. Nafamostat has shown to significantly reduce weight loss and lung tissue SARS-CoV-2 titres in murine models [8]. Nafamostat has a short half-life and poor oral bioavailability, which necessitates intravenous administration, limiting the potential use of the current formulation outside of a hospital setting. It has been used to treat disseminated intravascular coagulation (DIC), acute pancreatitis, and as an anticoagulant in extracorporeal hemofiltration and dialysis since the 1980s. In addition to the potential antiviral effects, Nafamostat inhibits platelet aggregation, inhibits thrombin, kallikrein, plasmin and other complement factors and reduces endothelial activation [9]. Given the prominent activation of thrombotic pathways and endothelial inflammation in COVID-19 immunopathogenesis, these are potentially beneficial attributes.

In this context Nafamostat is a drug highlighted as a potential target due to its antiviral, immunomodulatory and anticoagulant effects. Nine trials are ongoing without testing whether at the current recommended dose and route of administration, it has the expected PK/PD and safety profile. In this context, we report the first detailed assessment of safety, PK/PD; immunology and coagulation effects of the drug at the recommended dose and route, using a platform RCT.

## METHODS

### Trial design and participants

The DEFINE trial is a platform, multicentre, randomised controlled open label trial. Participants were recruited between September 2020 and February 2021 from two teaching hospitals in the UK: The Royal Infirmary of Edinburgh and The Western General Hospital, Edinburgh. Randomisation was computer generated and patients were assigned Nafamostat (alongside SoC), another candidate therapy, or SoC alone in a 1:1:1:n ratio, using a minimisation procedure based on sex, age, BMI, and a history of diabetes. Here we report the finding for the Nafamostat arm compared to SoC. This trial has been registered on ISRCTN (https://www.isrctn.com/) ISRCTN14212905, and Clinicaltrials.gov (https://www.clinicaltrials.gov/) NCT04473053. The DEFINE trial has received full ethical approval from Scotland A REC (20/SS/0066), the MHRA (EudraCT 2020-002230-32) and NHS Lothian. The trial protocol has been reported previously. [10] In brief, patients were eligible for inclusion if they were over the age of 16, had confirmed SARS-CoV-2 infection by reverse transcription-polymerase chain reaction test (RT-PCR), and had chest radiographic changes consistent with COVID-19, or an additional oxygen requirement. Written informed consent was provided prior to any trial procedures being performed. If a patient lacked capacity, consent could be provided by their next of kin. Exclusion criteria included pregnancy, lactation, and the inability to reliably take or tolerate modes of delivery of the treatment. Additionally, patients were excluded if they required anticoagulation, antiplatelet therapy or potassium sparing diuretics which could not reasonably be withheld. A current or recent history of severe uncontrolled cardiac disease, diabetes mellitus, renal impairment or hepatic impairment, anaemia, thrombocytopaenia, hyponatraemia or hyperkalaemia, were also exclusion criteria (see pre-print protocol with full inclusion/exclusion criteria [10]).

### Endpoints

The primary endpoint was to evaluate the safety and tolerability of intravenous Nafamostat as an add on therapy for patients hospitalised with COVID-19 pneumonitis. Secondary endpoints were to explore the Pharmacokinetics/Pharmacodynamics (PK/PD) of Nafamostat; assess the response of key biomarkers during the treatment period; evaluate SARS-CoV-2 viral load over time. Clinical secondary endpoints included oxygen free days; the change in the oxygen saturations and fraction of inspired oxygen concentration (SpO2/FiO2 ratio); time to discharge; the use of kidney replacement therapy.

### Interventions

Participants randomised to intravenous Nafamostat were administered the drug as a continuous infusion at a dose of 0.2 mg/kg/hr through a dedicated intravenous cannula. Each 5 mg vial was reconstituted with 5 mL of 5 % dextrose, then the total dose was added to 1000ml 5 % dextrose. The infusion was prepared as per local guidelines and changed every 24 hours for a total of 7 days, or until discharge or withdrawal. In the event of biochemical side effects, namely clinically significant hyperkalaemia or hyponatraemia, treatment was ceased. Treatment was also terminated if there was a clotting event requiring anticoagulation or antiplatelet therapy, but trial participants continued to provide daily bloods, ECG and clinical assessments despite no longer receiving a trial medication.

SoC included all appropriate supportive measures for SARS-CoV-2 and approved therapies as per national guidance at the time including dexamethasone, remdesivir and tocilizumab as per NHS guidelines.

### Clinical and laboratory monitoring

Nursing and medical staff visited patients daily until discharge, withdrawal, or day 16 of participation. A list of daily observations and blood parameters recorded are listed in Table 3 (supplementary material). Symptoms were elicited from patients and recorded as adverse events (AE) if indicated. All patients had a chest x-ray performed clinically prior to screening and the results of this were documented.

### Pharmacokinetics

In the treatment group, blood samples were taken for PK analysis on day one prior to starting the intravenous Nafamostat infusion then at 50 minutes, 2 hours, and 6 hours after commencing the infusion. For eight patients in the Nafamostat arm, this was modified to a pre-dose sample and a sample 6-36 hours after starting the infusion to allow a steady state. Samples were analysed using reverse-phase liquid chromatography-mass spectrometry (RP-MS). All separations and analyses were performed using an Acquity Ultra Performance LC system (Waters Milford, MA, USA) connected to a Xevo G2-XS QTOF mass spectrometer (Waters Milford, MA, USA). A CORTECs UPLC T13 C18 (1.6 μm, 2.1 × 100 mm) (Waters Milford, MA, USA) column at 40 °C was used. The mobile phase A was: water plus 0.1% formic acid and the mobile phase B was: methanol plus 0.1% formic acid. The linear gradient used was: 0 min, 5%A; 4 min, 50% B; 12 min 99.9% B; 15 min, 99.9% B; 15.10 min, 5% B; 18 min, 5% B. The flow rate was 0.3 ml/min. Spectra were recorded in positive ion mode and parameters were set as follow: mass range, 50-900 Da; scan time, 0.5 sec; data format, continuum; collision energy 6 V; Ramp collision energy, 20-40 V. The source parameters were capillary (kV), 3; sampling cone, 40; source offset, 80; temperature, 140 °C; desolvation, 500; cone gas (L/h), 50; desolvation gas (L/h), 900. Calibration took place on the day of measurement and mass accuracy was confirmed to be <5 ppm. Mass spectral peaks were analysed to assess whether Nafamostat was converted to its inactive metabolite 4-guanidinobenzoic acid (4-GBA). To assess whether the conversion of Nafamostat to 4-GBA could have happened *ex-vivo* prior to spinning, whole blood was ‘spiked’ with an aliquot of Nafamostat (50ng/ml), and samples were incubated for different periods (t0-t80min). Plasma was generated at the time points and the samples were snap-frozen before the same analysis was performed using RP-MS. A standard curve was prepared to quantify 4-GBA levels in each sample. The area under the total ion chromatogram for each compound in each sample was integrated to obtain the intensity. Sample processing and analysis was performed using MestReNova Software.

### Viral Load

Qualitative and quantitative polymerase chain reaction (PCR) of oropharyngeal/nasal measurements for SARS-CoV-2 were taken from the final 37 participants in the trial.

Nasopharyngeal samples were collected and added to viral transport media (Remel MicroTest M4RT). A volume of 110μL of eluate containing purified RNA was obtained following automated extraction carried out on the NucliSENS® easyMag® (bioMérieux) using an ‘off-board’ extraction where 200μL of the sample was added to 2ml of easyMAG lysis buffer. Saliva was pre-treated with proteinase K whereby 200μL of sample was mixed with 25μL of molecular grade proteinase K and then inactivated by heating at 95°C for 10mins prior to extraction. Nasopharygeal and saliva samples were then tested using the Altona RealTime PCR kit (Hamburg, Germany). E (envelope) and S(spike) gene results were obtained. A threshold cycle (Ct) of 40 or less was used as a cut off for positivity. Ct values were converted to copies per mL by relating values to a standard linearity panel with values in copies/mL derived by digital droplet PCR (Quality Control for Molecular Diagnostics, Glasgow). The threshold for a negative PCR (Ct) was 40, however negative results were also reported as ‘0’. In order to convert Ct values to copies/mL, Ct values of 0 were standardised to 45. **Thromboelastometry**

Blood was sampled daily to perform thromboelastometry (TEM) using a CLOTPRO® analyser (Haemonetics). This enabled point of care evaluation of whole blood coagulation [11]. Measurements of anticoagulation effect, clot strength and antifibrinolytic effect were executed. Six different tests were performed including the TPA-test which measures the effect of antifibrinolytic drugs [12, 13]. Clotting time, maximum lysis percentage, lysis time and maximum clot firmness (MCF) using six different reagent panels were measured.

### Cytokine analysis

Plasma samples were prepared by centrifuging EDTA-blood at 1,400g for 10 minutes at 4□. Samples were frozen on dry-ice in aliquots and stored at -80°C and assayed at the end of the trial using the ELLA platform (Simple protein, Bio-Techne, R&D, USA). Results beyond the limit of detection and intra-cartridge coefficient of variation (CV)% higher than 10% were removed from analysis, the inter-cartridge CV% were within 20%.

### Flow cytometry

All staining and processing were performed in an MSCII biosafety cabinet with centrifugation steps using capped tubes in the biosafety bucket which were loaded and unloaded within the MSCII Peripheral blood was taken for detailed immunophenotyping. Red blood cells were lysed using BD FACS lyse. 100 μl of whole blood/EDTA was added to FACS tubes containing 5 μl of Fc blocker TruStain FcX (Biolegend 422301) and 5 μl Monocyte blocker (Biolegend 0426102). 5 minutes later, 50 μl of antibody staining cocktail (prepared in FACS staining buffer (PBS 2% FCS (Gibco)) containing 10% Brilliant violet plus buffer (BD 566385)) was added to each tube and then incubated in the dark, at room temperature for 20 minutes and washed twice in staining buffer before fixation (Biolegend Fixation buffer). After 20 minutes, fixed samples were moved from the MSCII to cold storage. All samples were analysed on a 5 laser BD LSR Fortessa within 24 hrs of staining. Freshly prepared 8 peak calibration beads were run daily prior to sample collection. Gating strategies used to identify cellular subpopulations are illustrated in supplementary Figure 8A-E and a list of all the antibodies used is provided in Table 4 (supplementary material).

### Statistics

The analysis population consisted of (i) all patients randomised to Nafamostat who received any dose of the trial drug and (ii) all patients randomised to the control arm (SoC) who would also have been eligible to receive Nafamostat. Therefore, any patients who were randomised to Nafamostat but did not receive the trial drug were excluded.

Descriptive statistics were calculated for all primary endpoints, split by day of measurement (post-randomisation) and treatment group. Bayesian Generalised Linear Mixed Effects models (GLMM) were used to compare continuous outcomes between the Nafamostat and control arms, with a random effect for patient, and adjusting for baseline and day of measurement post-randomisation (as a factor variable). Posterior mean differences and highest posterior density (HPD) intervals were reported (Table 6). Non-informative flat priors were used.

For binary categorical outcomes, composite binary variables were derived at the patient level, combining outcome data across all timepoints, due to sparse binary data. For example, for “abnormal respiratory examination”, we recorded whether each patient had any abnormal respiratory examination across all time points. A Bayesian logistic Generalized Linear Model (GLM) was fitted to each composite binary outcome, and the trial arm was included as the only explanatory variable.

Bayesian Poisson GLMs were used to compare the rate of oxygen free days and number of AEs for each patient between trial arms. We also separately analysed the outcome of “at least one AE during follow-up” using a Bayesian logistic regression model, with trial arm as the only explanatory variable. All statistical models were fitted using SAS software (SAS Institute Inc., Cary, NC, USA).

For TEM, viral and flow cytometry data, one-way ANOVA, mixed-effects model and student’s t-test were used. The cytokine data were presented as means and 95% confidence intervals with best-fit line by linear regression and comparing intercepts and slopes. The analyses were performed using two-sided tests and the GraphPad Prism 8 program. All patients randomised to Nafamostat were included in the analysis, regardless of whether they remained on the infusion. Two subjects in Nafamostat arm and one subject in SoC arm were not included in the cytokine analysis as plasma samples were unobtainable.

## RESULTS

### Participants

Amongst 299 individuals screened, 66 met eligibility criteria and were randomised. 44 participants were enrolled to the Nafamostat vs SoC comparison reported here. 22 participants were randomised to a third arm. There were no baseline differences between Nafamostat and SoC groups (Table 1; Figure-1).

**Table 1:** Patient demographics at time of enrolment

|  | Randomisation Result |  |  |  |  |  | Total (n=42) |  |  |
| --- | --- | --- | --- | --- | --- | --- | --- | --- | --- |
|  | Standard of care (n=21) |  |  | Nafamostat (n=21) |  |  |  |  |  |
| Mean Age (years) | 65.0 [ range 27-87] |  |  | 62.3 [range 31-85] |  |  | 63.6 [range 27-87] |  |  |
| Mean BMI (kg/m2 ) | 32.4 [range 22.3- 44.6] |  |  | 32.5 [range 22.6- 47.5] |  |  | 32.4 [range 22.3- 47.5] |  |  |
| Mean number of days since symptom onset | 8.6 [range 3-15] |  |  | 8.5 [range 3-17] |  |  | 8.5 [range 3-17] |  |  |
| Sex | M 57.1%(n=12) |  | F 42.9% (n=9) | M 61.9% (=13) |  | F 38.1% (n=8) | M 59.5% (n=25) |  | F 40.5% (n=17) |
| Ethnicity | White 90.5% (n= 19) |  |  | White 90.5% (n= 19) |  |  | White 90.5% (n=38) |  |  |
|  | Asian 4.8% (n= 1) |  |  | Asian 9.5% (n=2) |  |  | Asian 7.1% (n= 3) |  |  |
|  | Black 4.8% (n=1) |  |  | Black 0% |  |  | Black 2.4% (n=1) |  |  |
| Smoking status | NS 52.4% (n=11) | S 0% | Ex-S 47.6% (n=10) | NS 61.9% (n=13) | S 4.8% (n= 1) | Ex-S 33.3% (n=7) | NS 57.1% (n=24) | S 2.4% (n= 1) | Ex-S 40.5% (n=17) |
| Diabetes | Y 14.3% (n=3) |  | N 85.7% (n=18) | Y 28.6% (n=6) |  | N 71.45 (n=15) | Y 21.4% (n=9) |  | N 78.6% (n=33) |
| Hypertension | Y 38.1% (n=8) |  | N 61.9% (n=13) | Y 38.1% (n=8) |  | N 61.9% (n= 13) | Y 38.1% (n=16) |  | N 61.9% (n=26) |
| Chronic Cardiac Disease | Y 23.8% (n=5) |  | N 76.2% (n=16) | Y 4.8% (n=1) |  | N 95.2% (n=20) | Y 14.3% (n=6) |  | N 85.7% (n=36) |
| Asthma | Y 23.8% (n= 3) |  | N 76.2% (n=18) | Y 4.8% (n=1) |  | N 95.2% (n= 20) | Y 9.5% (n= 4) |  | N 90.5% (n=38) |
| Chronic kidney disease | Y 4.8% (n=1) |  | N 95.2% (n=20) | Y 4.8% (n=1) |  | N 95.2% (n=20) | Y 4.8% (n=2) |  | N 95.2% (n= 40) |
| Chronic liver disease | Y 0% |  | N 100% (n= 21) | Y 0% |  | N 100% (n= 21) | Y 0% |  | N 100% (n= 42) |
| Malignancy | Y 14.3% (n=3) |  | N 85.7% (n= 18) | Y 4.8% (n=1) |  | N 95.2% (n=20) | Y 9.5% (n= 4) |  | N 90.5% (n=38) |
| Immunocompromised | Y 9.5% (n= 2) |  | N 90.5% (n=19) | Y 4.8% (n=1) |  | N 95.2% (n=20) | Y 7.1% (n=3) |  | N 92.9% (n= 39) |

**Figure 1:**
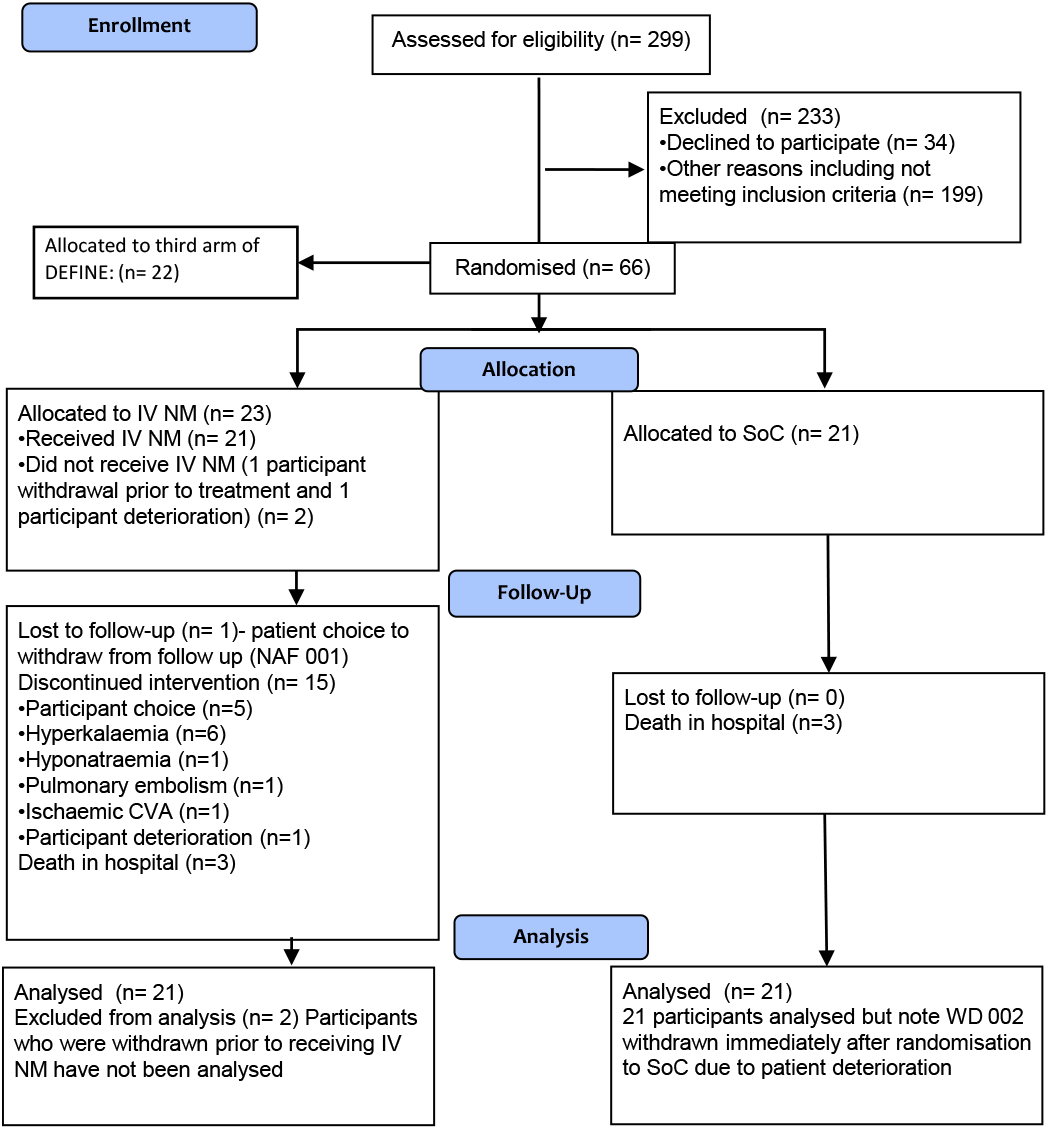
Consort flow diagram

### Adverse events

Patients’ clinical course, in-hospital AE occurrence and time in the trial are summarised for each arm in Figure 2A and Table 6.

**Figure 2:**
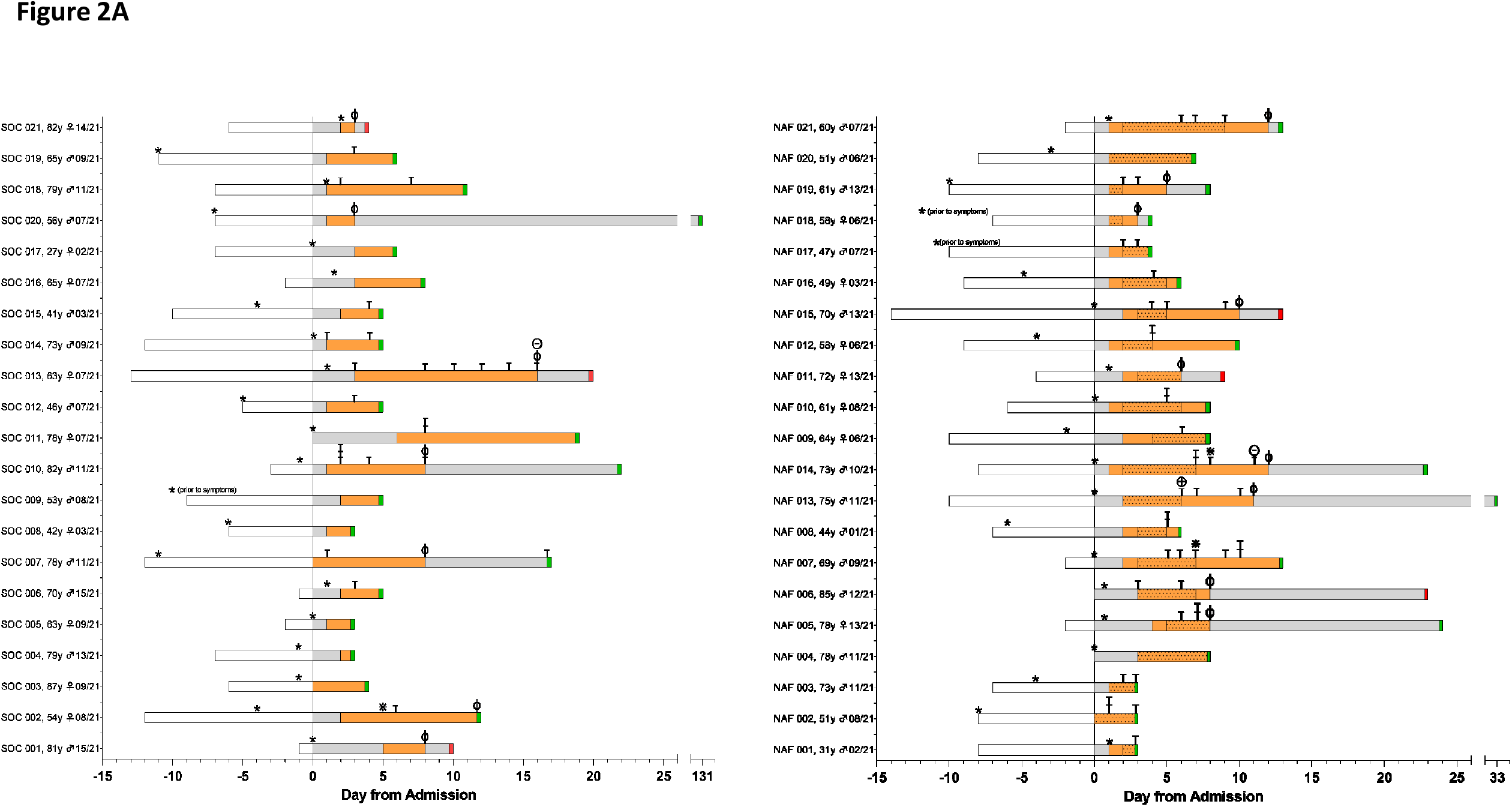
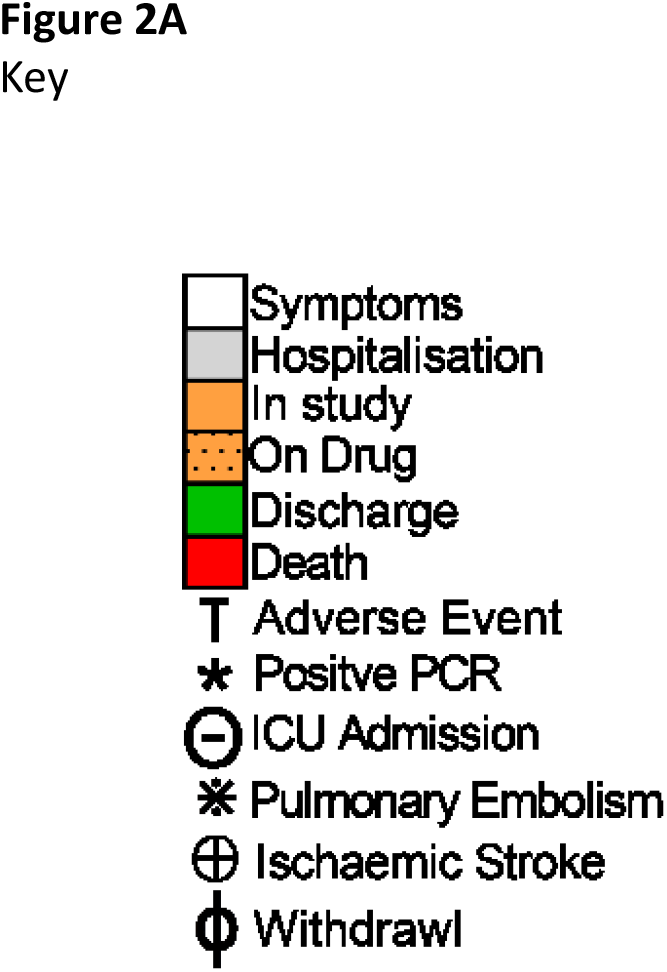
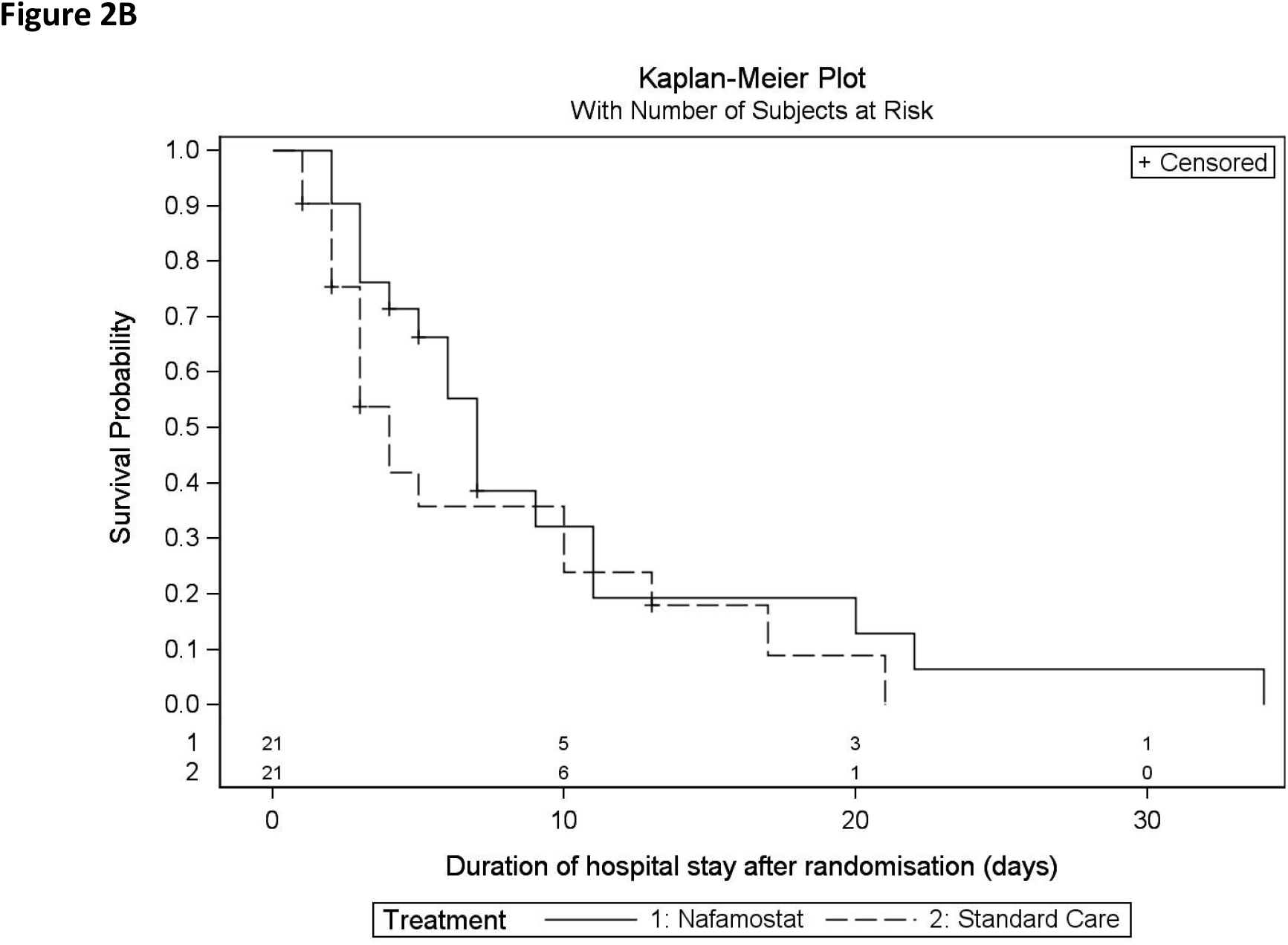
**A**: Clinical course for each patient from hospital admission to discharge or death. Study identifier code, age, gender and ISARIC score (score 0-21 points) given on y-axis with clinical course, time on trial as well as key clinical events reported. **B:** Kaplan-Meier plot reporting duration of hospital stay following randomisation.

The Nafamostat group experienced more AEs compared to SoC (n=50 vs n=35), with 78% of Nafamostat-treated patients experiencing at least one AE compared to 57% of the SoC group. There were no serious adverse events (SAEs) in either group. Of 21 patients who received Nafamostat, a total of 15 stopped prior to the 7 day planned course. (Figure 2A). Other than clinical deterioration, hyperkalaemia was the most common reason for early cessation (6/21), although there were no related complications. One patient developed a pulmonary embolism, and one patient suffered an ischaemic CVA whilst on Nafamostat (Figure 2A).

### Clinical endpoints

A Kaplan-Meier plot of duration of hospital stay is shown in Figure 2B, with an average longer hospital stay in Nafamostat patients (Table 2). Nafamostat-treated patients were on oxygen for a median of 2 days more than SoC patients (Table 2) and there were a significantly lower number of oxygen free days for those on Nafamostat (rate ratio 0.55-95% HPD interval 0.31-0.99). Serum creatinine was significantly higher following intravenous Nafamostat administration compared to SoC (posterior mean difference 10.57 micromol/L, 95% HPD interval 2.43 - 18.92). There were no other statistically significant differences regarding primary endpoints. Each group had three deaths, all attributed to worsening COVID infection. There were no changes noted in either group for other safety endpoints such as the daily physical exam or daily ECG (Table 6).

**Table 2:**
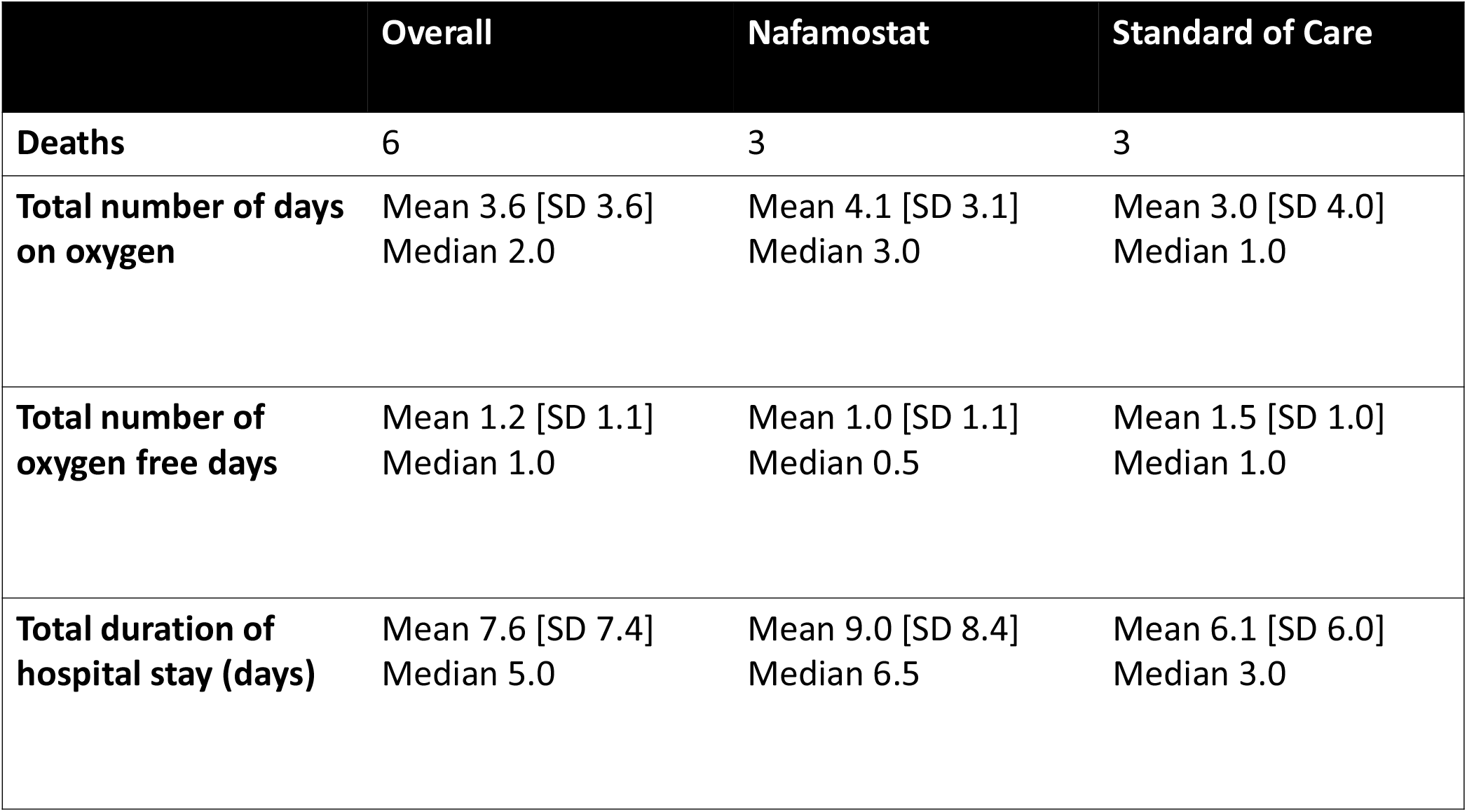
Hospital stay, days on oxygen and oxygen free days across the two groups. There was a significantly lower number for the rate of oxygen free days for those using IV NM when adjusted for the length of time the patient was in hospital and recording outcomes (rate ratio 0.552, 95% HPD 0.307-0.993).

### Pharmacokinetics

Blood samples for the analysis of plasma Nafamostat levels and its breakdown metabolite (4-GBA) were obtained. For each patient, the post/pre-infusion ratio of Nafamostat levels revealed almost undetectable levels Nafamostat (Figure 3A), with the exception of two participants who showed higher levels (NAF006 and 012). To determine Nafamostat breakdown products, levels of 4-GBA were measured pre-infusion (Figure 3B). 4-GBA was undetectable in the pre-infusion samples confirming no intrinsic 4-GBA. Following Nafamostat administration, 4-GBA was detected at elevated levels in plasma (Figure 3B and 3C). To ensure this was not an artefact of breakdown following blood draw, whole blood was spiked with Nafamostat over 80 minutes without observing breakdown products (Figure 3D). Taken together, this suggests in hospitalised patients with COVID-19 pneumonitis, there is rapid breakdown of intravenous Nafamostat to its inactive metabolite, 4-GBA, resulting in very low levels of circulating Nafamostat, and intravenous Nafamostat therefore had unfavourable PK characteristics in this cohort of patients.

**Figure 3.**
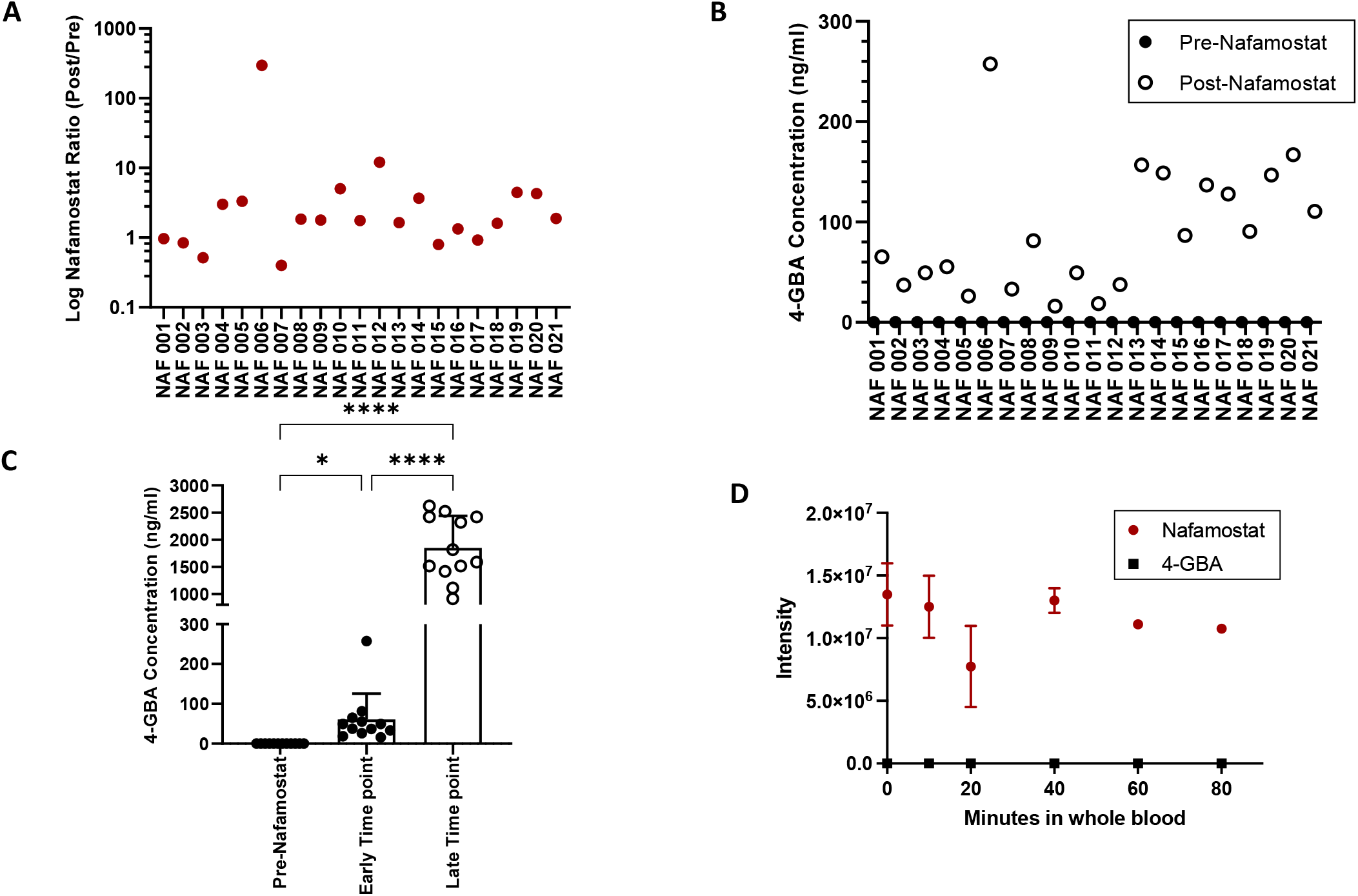
Pharmacokinetics of IV NM. **A**: Ratio of Post-NM to Pre-NM in each participant in the NM arm, measured by mass spectrometry with a detection m/z [M+H]+=348.1455. **B**: Assessment of NM metabolite 4-GBA in samples by mass spectrometry at detection of m/z [M+H]+= 180.0767. **C**: 4-GBA concentration in patients samples taken pre-infusion and at early and later time points (plotted for 12 patients in whom matched samples available). **** p value <0.0001, * p value = 0.02. Error bars: SD. **D**: To assess whether the conversion of NM to 4-GBA could have happened *ex-vivo* prior to spinning, whole blood was ‘spiked’ with an aliquot of NM (50ng/ml), and samples were incubated for different periods (t0-t80min). Plasma was generated at the time points and the samples were snap-frozen before analysis performed using RP-MS. A standard curve was prepared to quantify 4-GBA levels in each sample. The area under the total ion chromatogram for each compound in each sample was integrated to obtain the intensity. Error bars: SEM.

### Viral data

Nasopharyngeal and saliva samples were taken at baseline, day 3 and day 5 for RT PCR analysis. Viral load decreased over time in both groups, with no difference observed between the Nafamostat and SoC groups (Figure 4 A-D).

**Figure 4.**
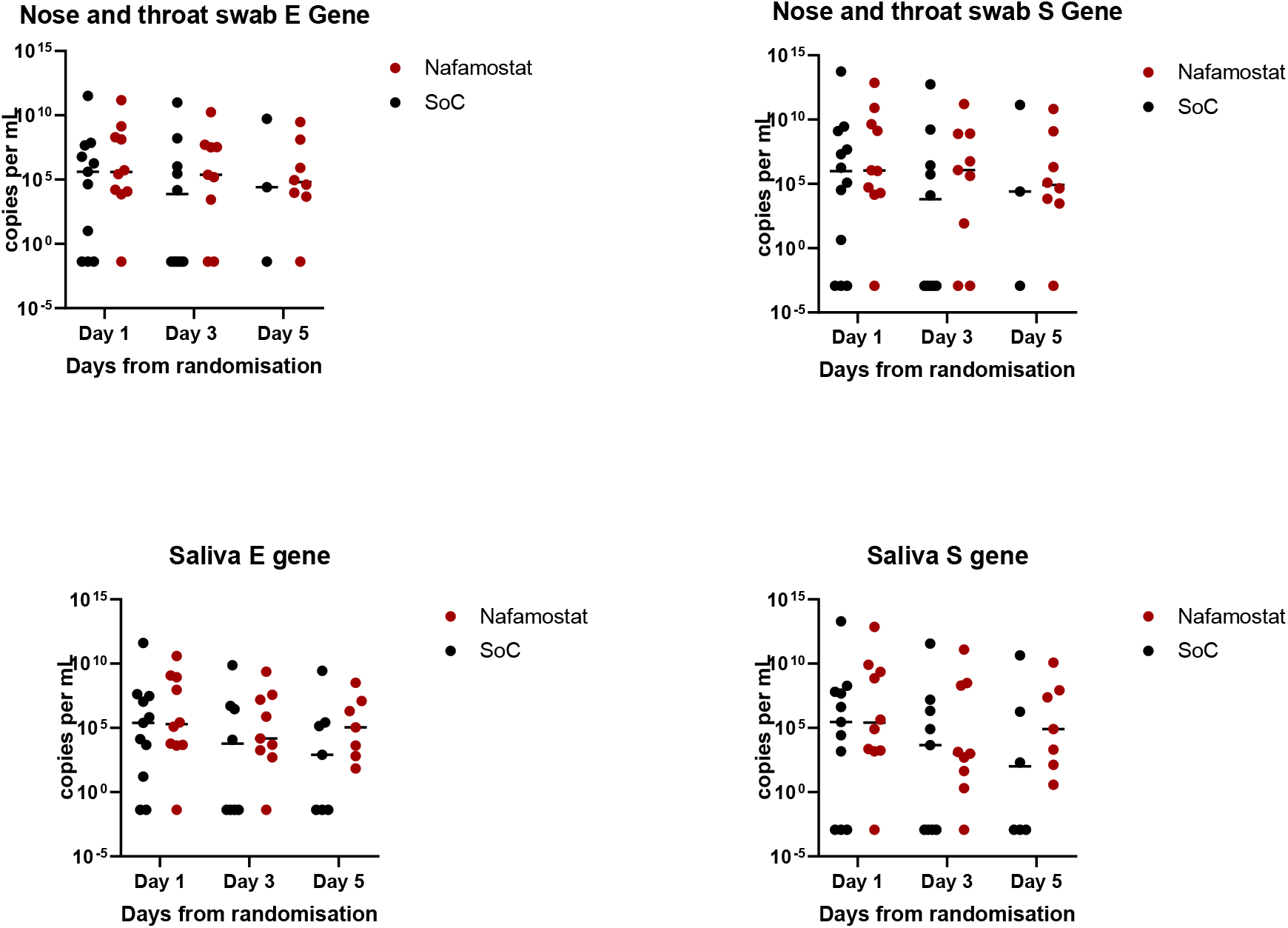
**A** and **B** – copies per mL of E and S gene respectively in nasopharyngeal and oropharyngeal swabs **C** and **D** – copies per mL of E and S gene respectively in saliva In both groups, the numbers of participants declined with time representing participant discharge or withdrawal (Nafamostat D1 n= 10, D3 n= 9, D5 n= 8. SOC D1 n=11, D3 n=10, D 5 n= 3).

### Thromboelastometry

In most patients receiving intravenous Nafamostat, little or no anticoagulant effect was evident (Figure 5A). However, an antifibrinolytic effect was seen in patients receiving Nafamostat (Figure 5B). When Nafamostat produces an antifibrinolytic effect with little or no anticoagulant effect, the overall effect is likely to be prothrombotic rather than anticoagulant.

**Figure 5:**
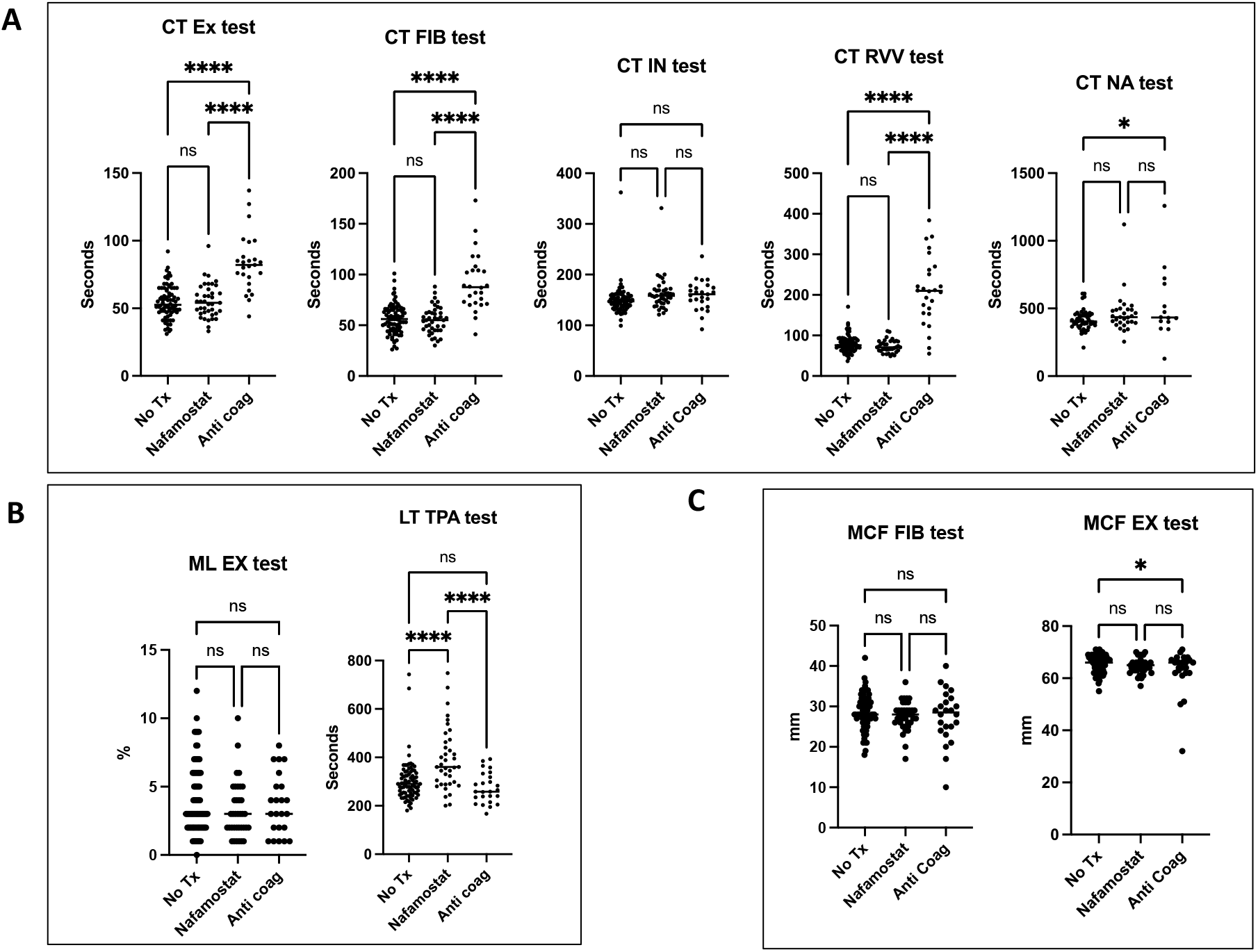
Thromboelastometry (ClotPro^®^). Blood tests performed daily on all patients. Presented here as pooled groups depending on treatment given at time of blood draw. Results pooled to three groups- no anticoagulant treatment (No Tx), on IV NM (Nafamostat), on other therapeutic anticoagulation e.g. apixaban/ dalteparin (Anti coag). Significance by one-way ANOVA and Student’s t-test. **A**: Panel evaluating anticoagulant activity (clotting time). **** p <0.001, * p=0.02 **B:** Panel evaluating antifibrinolytic activity (lysis time and maximum lysis percentage). **** p <0.0001 **C:** Panel evaluating clot strength (maximum clot firmness). * p = 0.04 Six ClotPro tests were performed on whole blood samples daily: EX-test: assessment of coagulation with extrinsic activation using recombinant tissue factor. FIB-test: assessment of coagulation with extrinsic activation but in the presence of inhibitors of platelet aggregation. Measures the fibrin contribution to clot strength. IN–test: assessment of coagulation with intrinsic activation using ellagic acid. RVV-test: screening test for direct FXa antagonists, also sensitive to thrombin antagonists. NA-test: non-activated test - assessment of coagulation without an activator of coagulation. TPA-test: assessment of coagulation with fibrinolysis activation (using recombinant tissue plasminogen activator). Measures the effect of anti-fibrinolytic drugs.

### Laboratory investigations, cytokines and biomarkers

A significantly higher creatinine level was observed in patients randomised to Nafamostat (posterior mean difference 10.57 micromol/L, 95% HPD interval 2.43 - 18.92) (Table 5 and Figure 6E, supplementary material). Otherwise, biochemical and haematological safety laboratory investigations were not significantly different between the groups (Figure 6, supplementary material).

**Figure 6:**
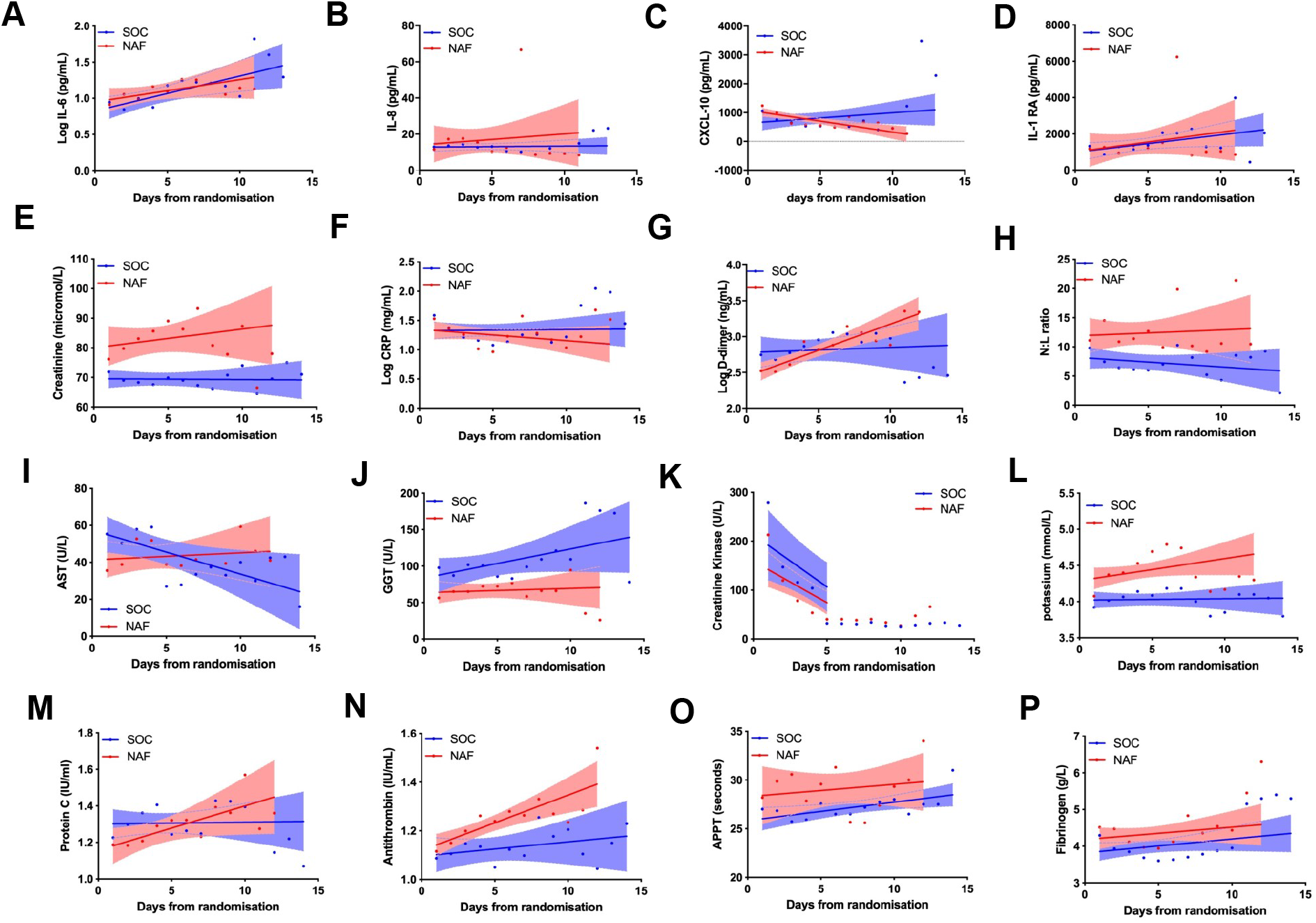
Biomarkers over time. SOC (blue)- standard of care. NAF (pink)- Nafamostat Mesylate **A-D**: Changes of COVID-19 cytokine storm related cytokines IL-6, IL-8, CXCL-10, IL-1RA. **E-L**: Clinical biomarkers : creatinine, CRP, D-dimer, neutrophil: lymphocyte ratio, AST, GGT, creatinine kinase and potassium. **M-P**: Biomarkers of coagulation such as protein C, antithrombin, APTT & fibrinogen. Data presented as mean with best-fit line and 95% confidence intervals by linear regression. All NM patients included in analysis including when off infusion.

Most COVID-19-related cytokine biomarkers showed similar trajectories between randomisation and 16-days, in both the Nafamostat and the SoC groups (Figure 6 A-D, supplementary material).

Clinical biomarkers showing disease severity were not improved by Nafamostat compared to SoC (Figure 6 & 7), on the contrary D-dimer levels rose when patients were treated with intravenous Nafamostat (Figure 6G). Nafamostat patients trended towards higher total white blood cell counts, neutrophils, monocytes, and lower lymphocytes counts over the course of treatment (Figure 8 J-M, supplementary material). Antithrombin levels increased throughout treatment in the Nafamostat group (Figure 6N). A similar trend was seen with Protein C (Figure 6M).

**Figure 7.**
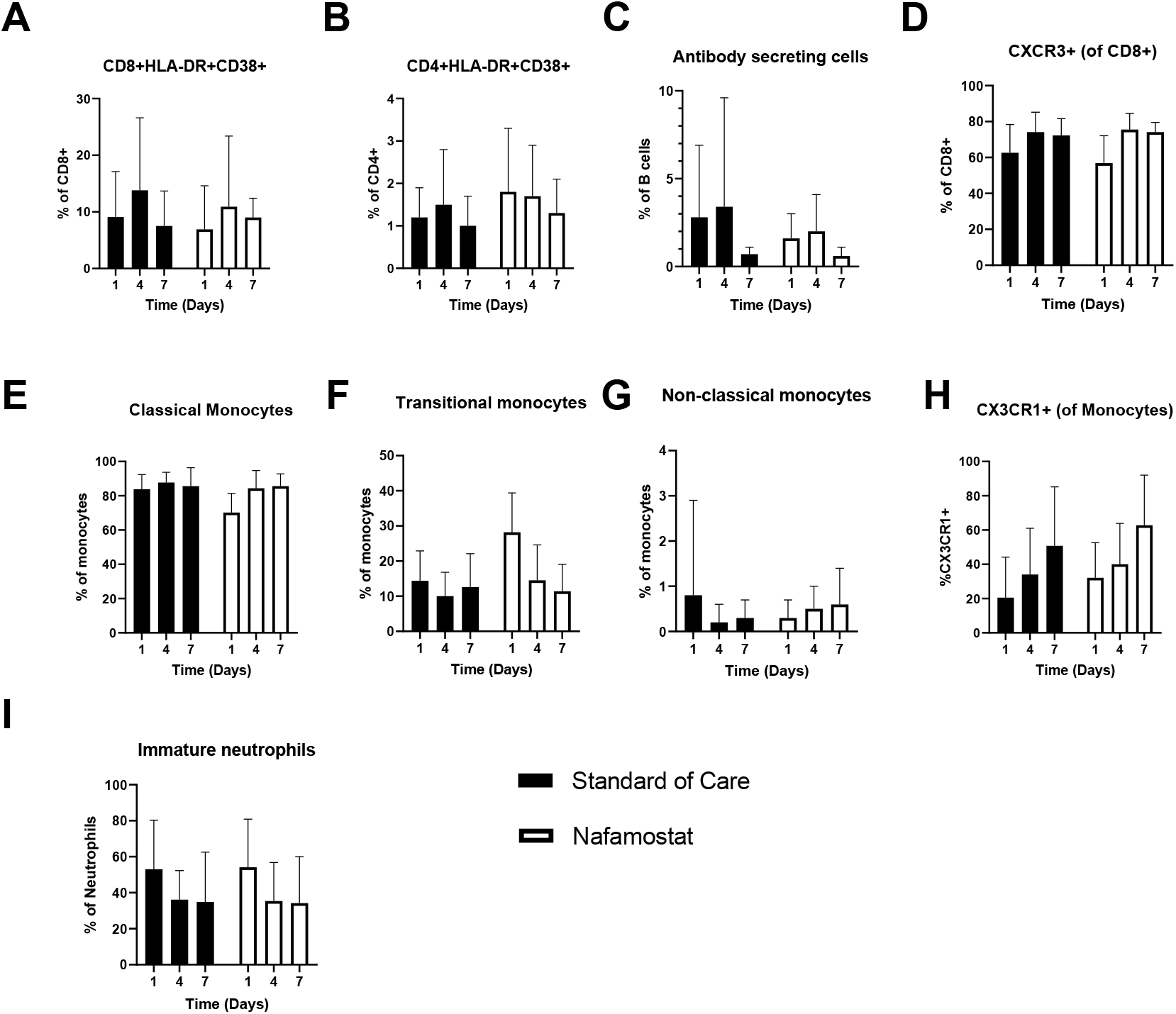

### Peripheral blood immunophenotyping

To assess immune perturbations associated with COVID-19 infection, and whether kinetic changes in the immune response correlated with treatment, flow cytometry was used to characterise peripheral blood leukocytes on entry to the trial (pre-treatment), and on day four and day seven whilst hospitalised. Evidence of activation of the adaptive immune response during COVID-19 infection was seen as the presence of HLA-DR+CD38+ activated T cells and CD19+CD27+CD38+ antibody secreting cells (ASC) (Figure 7 A-C). Nafamostat treatment did not alter the proportion of activated T cells or antibody secreting cells. Expression of the chemokine receptor CXCR3[14] was initially low in the CD8+ T cells of COVID-19 patients and increased during time on trial in both Nafamostat and SoC groups (Figure 7D). Within the monocyte population, there were low levels of non-classical monocytes, as has been described in COVID-19 patients[7] and this was not recovered during the time on trial (Figure 7 E-G). We observed low expression of the fractalkine receptor (CX3CR1) by monocytes from COVID-19 infected individuals at baseline which increased over time and was not influenced by Nafamostat treatment (Figure 7H). In summary, Nafamostat did not influence the rate of change in any immune parameters (Figure 7).

## DISCUSSION

Nafamostat was delivered at its recommended dose for its licenced indication. Intravenous Nafamostat treatment compared to SoC produced more AEs without any evidence of beneficial effects. Intravenous Nafamostat had an unfavourable PK profile when used in patients with COVID-19.

Compound screens have revealed that Nafamostat is one of the most potent inhibitors of SARS-CoV-2 viral entry into lung epithelial cells, with a greater than 600-fold potency over other antivirals [14]. As Nafamostat has anticoagulant and anti-inflammatory effects, the combination of these properties has led to intravenous Nafamostat being promoted as a promising therapeutic for COVID-19 [15, 16]. The intravenous formulation is currently under investigation in nine phase II/III clinical studies globally including the ASCOT trial, however to our knowledge none have fully reported results, or examined PK/PD[17]. Indeed, given the pressing nature of COVID-19, many therapeutics have progressed immediately to large phase II/III trials with no prior investigation of PK or safety in this population of unwell patients. Thus, to assess safety and PK of intravenous Nafamostat, we performed an incisive experimental medicine clinical trial in a small cohort of hospitalised patients with COVID-19 pneumonitis. In this cohort whilst there were no SAEs, there were a larger number of AEs within the intravenous Nafamostat treatment group and patients required oxygen for longer. The treatment course was discontinued early in 29% of patients indicating poor tolerability. We observed six cases of moderate hyperkalaemia in the intravenous Nafamostat arm, which prompted the end of treatment. Hyperkalaemia is a well described and widely reported side effect of intravenous Nafamostat owing to the effect of the metabolites on sodium conductance in the renal cortical collecting duct, thereby impairing urinary potassium excretion [18]. Notably, significantly higher creatinine levels were observed for patients randomised to intravenous Nafamostat compared to SoC.

Intravenous Nafamostat demonstrated a poor PK profile with undetectable levels in most patients with COVID-19 pneumonitis. Chemically, Nafamostat is an ester conjugate of p-guanidinobenzoic acid (GBA) and 6-amidino-2-naphthol. The ester site is described as the “reaction centre” as well as the site for the catabolic changes. Nafamostat has been reported to inhibit the activity of TMPRSS2 with an IC50 of between 5-55nM[7]. It is known that the plasma half-life (t1/2) of Nafamostat is short - between 23.1min and 1.84hr[19] and thus a continuous infusion is required to achieve a steady-state concentration sufficient to inhibit therapeutic targets. In this trial we used a dose of 0.2mg/kg/hr which is the current clinically licensed dose of Nafamostat for DIC and pancreatitis in Japan. The half-maximal inhibitory concentration (IC50) of Nafamostat in preventing infection of alveolar epithelial cells by SARS-CoV-2 is in the range of 5-10 nM[20]. The steady-state plasma concentrations of Nafamostat when infused to patients with DIC at 0.1 mg/kg/hr or 0.2 mg/kg/h is reported to be between 14-130 ng/mL, which exceeds the IC50 for TMPRSS2. PK analysis shows that with the exception of two patients, negligible levels of Nafamostat was circulating when administered by continuous intravenous infusion in COVID-19 patients. Detectable levels of the metabolite support a rapid breakdown *in vivo*. Human arylesterases and carboxylesterase 2 are predominantly responsible for the metabolism of Nafamostat in the blood and liver, respectively [21]. Whilst we did not measure activity of these pathways in our patients, it is possible that these may be modulated in COVID-19, either due to the intrinsic disease process or due to concomitant therapy with corticosteroids [22] which the majority of patients received as SoC. It should be noted however that corticosteroid doses used in COVID-19 are lower than those expected to elicit clinical drug interactions via other metabolic processes like CYP3A4 [23].

The trial was not powered to assess clinical efficacy, and we did not observe any trends suggestive of intravenous Nafamostat improving biomarker endpoints in our cohort of patients. It is well documented that COVID-19 patients are at risk of micro and macrovascular clotting events, with venous thromboembolic events occurring in approximately 25% - 30% of patients [24]. This is twice the incidence of other critically unwell patients [25] and is associated with poor outcomes [26]. The coagulopathy found in COVID-19 disease is characterised primarily by a raised D-dimer, with a normal to raised fibrinogen, and only mildly elevated or normal thrombin and platelet counts [27]. This, alongside the lack of bleeding diathesis is not typical of DIC or bacterial sepsis induced coagulopathy. A potential method to observe hypercoagulability in COVID-19 is TEM. It provides measurements of clot formation, clot strength and clot lysis by recording the elasticity of a blood clot from the formation of fibrin for a standardised period of time. Exploratory studies examining the use of TEM in COVID-19 have confirmed hypercoagulable profiles with derangement of fibrinogen and platelet function [28]. This was supported by the data we observed. In most patients receiving intravenous Nafamostat, little or no anticoagulant effect was evident, however an antifibrinolytic effect was observed. The antifibrinolytic effect could potentially have a procoagulant effect, though the size of the effect was small compared to that seen when an antifibrinolytic drug such as tranexamic acid is given [27,28]. These TEM results were supported by routine laboratory coagulation screening which demonstrated that with the exception of one patient, we did not observe a raised APTT in our treatment cohort.

Alongside coagulation profiles, biomarkers of COVID-19 disease severity were not improved with Nafamostat when compared to SoC. Elevated numbers of neutrophils and a higher frequency of immature neutrophils are seen in COVID-19 infection and associated with poor prognosis [29, 30]. An initially high frequency of immature neutrophils (expressing low levels of CD10) at baseline rebalanced in subsequent sampling but the rate of normalisation (with increasing MFI of CD10 and decreasing MFI of CD66b) was not impacted by intravenous Nafamostat. In summary, intravenous Nafamostat did not influence the rate of change in any immune or inflammatory parameters showing measurable alterations during the time on trial.

In conclusion, treatment of patients hospitalised with COVID-19 using continuous intravenous Nafamostat was poorly tolerated, demonstrating more AEs than the SoC group. There were undetectable plasma levels of Nafamostat in most patients. The small number of patients enrolled in the trial, makes it difficult to draw conclusions regarding potential efficacy of intravenous Nafamostat. However, there was no evidence to suggest a change in the clinical course and no difference between the key biochemical, haematological or cytokine trends between the Nafamostat or SoC groups. Furthermore, no significant anticoagulant or antiviral effect was observed. This experimental medicine trial with extensive phenotyping of PK and PD, does not support the use of intravenous Nafamostat in hospitalised COVID-19 patients. However, the antiviral mechanism of action as an entry inhibitor [31], coupled with knowledge of cell-cell transmission of SARS-CoV-2 once active infection is established [8, 32] may warrant further evaluation of Nafamostat via different routes of administration. For example, data recently suggested a putative role in pre-exposure prophylaxis and post-exposure early therapy via intranasal administration [8, 33]

## Data Availability

Ownership of the data arising from this study resides with the study team. Scientific publications and the sharing of clinical data generated as part of this trial is crucial to better understanding COVID-19 and developing new treatments. As such, the results will be submitted for publication in a peer-reviewed journal. Data will be shared with larger clinical trial networks, including RECOVERY.

## Acknowledgements and funding

We thank Nichi-Iko (Japan) for kindly supplying Futhan (intravenous Nafamostat) for the trial; The Clinical Research Facility and staff in the Royal Infirmary of Edinburgh; the EMERGE research team; the ACCORD team at NHS Lothian/ University of Edinburgh, the patients and their families for taking part; LifeArc (an independent medical research charity) for providing funding for the clinical trial under the STOPCOVID award to the University of Edinburgh. We also thank the Oxford University COVID-19 Research Response Fund (BRD00230).

