## supplementary material for "Randomised Controlled Trial of Intravenous Nafamostat Mesylate in COVID pneumonitis: Phase 1b/2a Experimental Study to Investigate Safety, Pharmacokinetics and Pharmacodynamics"

### Slide 1
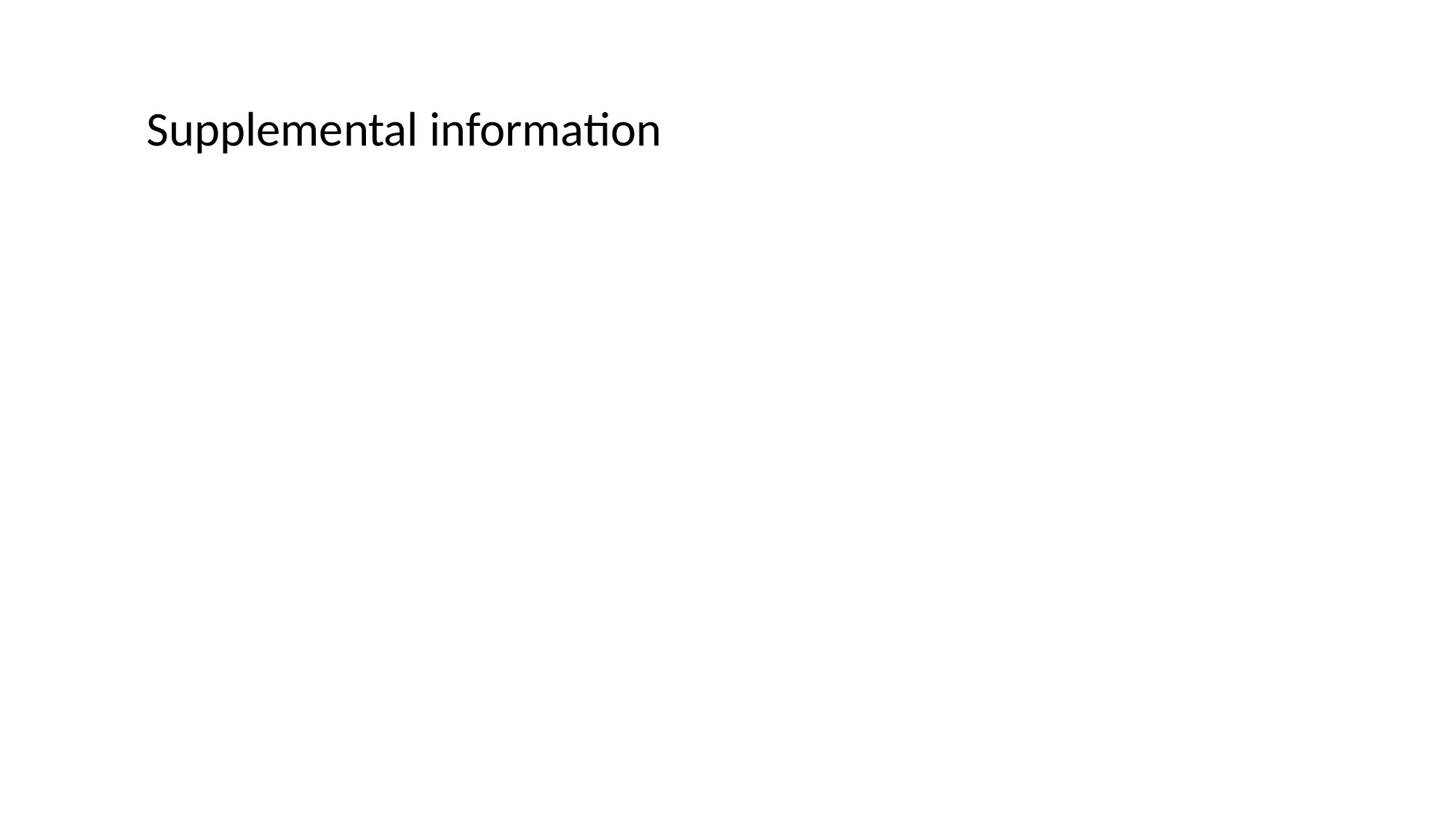

Supplemental information

### Slide 2
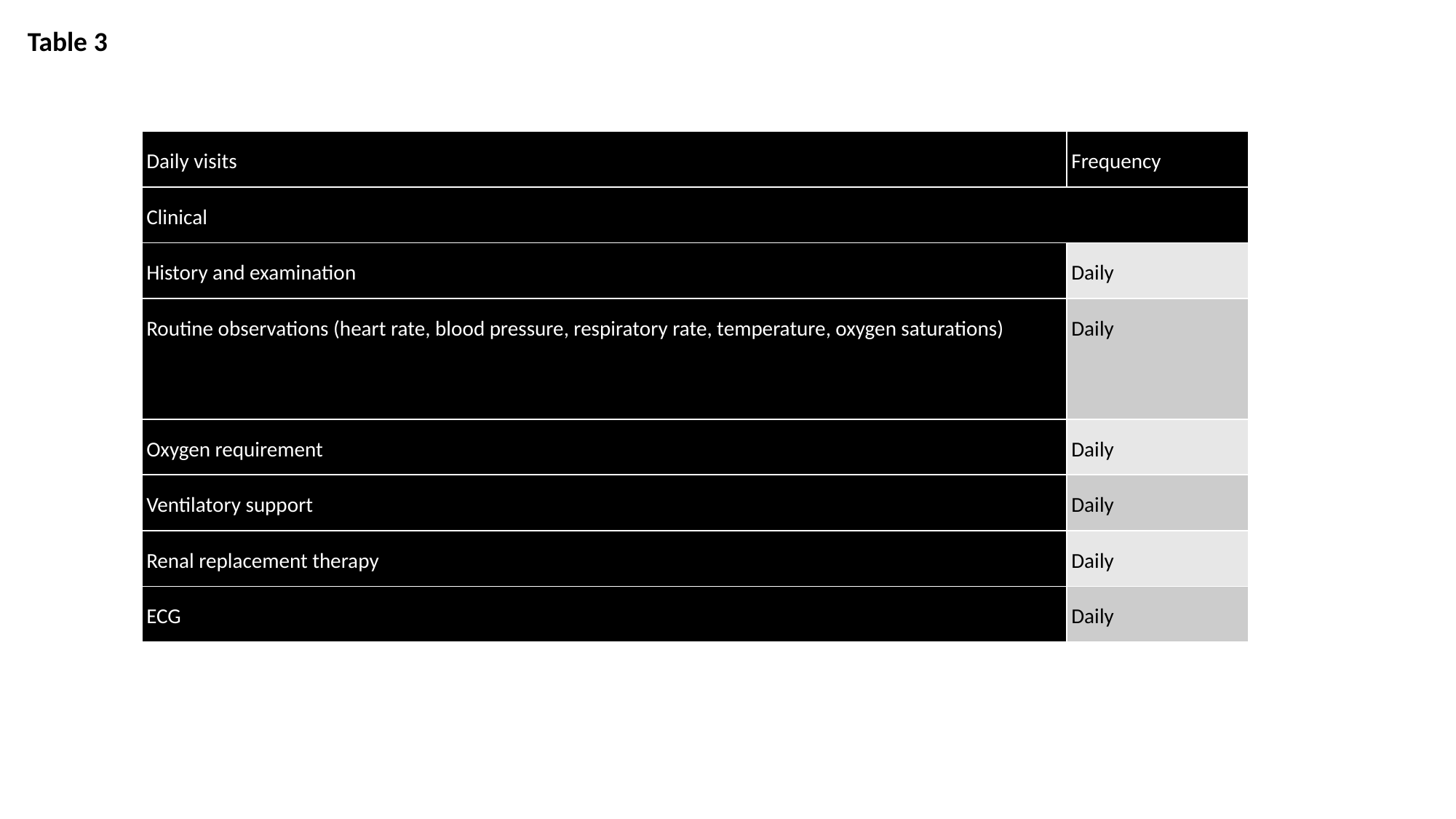

Table 3
| Daily visits | Frequency |
| --- | --- |
| Clinical | |
| History and examination | Daily |
| Routine observations (heart rate, blood pressure, respiratory rate, temperature, oxygen saturations) | Daily |
| Oxygen requirement | Daily |
| Ventilatory support | Daily |
| Renal replacement therapy | Daily |
| ECG | Daily |

### Slide 3
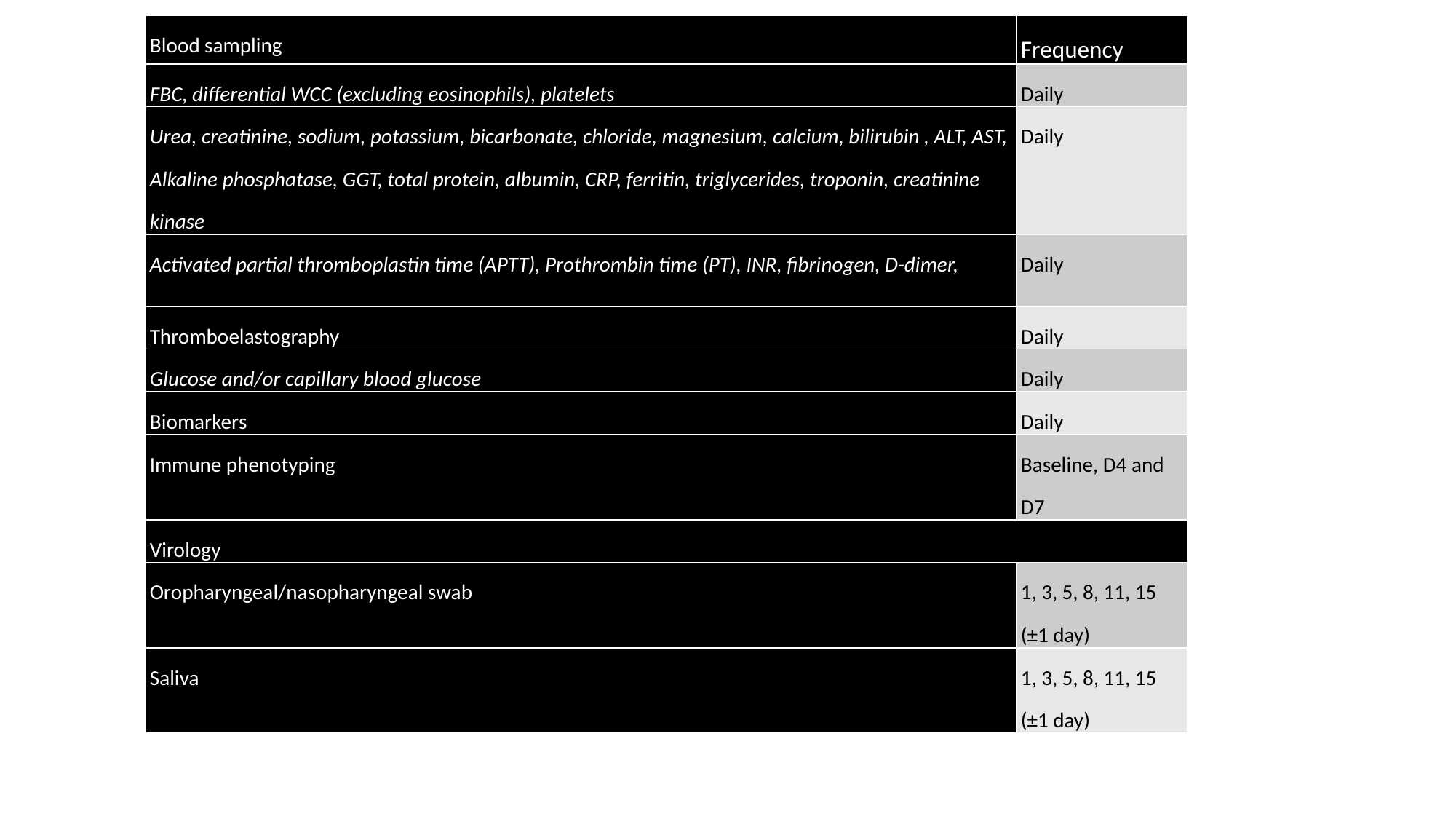

| Blood sampling | Frequency |
| --- | --- |
| FBC, differential WCC (excluding eosinophils), platelets | Daily |
| Urea, creatinine, sodium, potassium, bicarbonate, chloride, magnesium, calcium, bilirubin , ALT, AST, Alkaline phosphatase, GGT, total protein, albumin, CRP, ferritin, triglycerides, troponin, creatinine kinase | Daily |
| Activated partial thromboplastin time (APTT), Prothrombin time (PT), INR, fibrinogen, D-dimer, | Daily |
| Thromboelastography | Daily |
| Glucose and/or capillary blood glucose | Daily |
| Biomarkers | Daily |
| Immune phenotyping | Baseline, D4 and D7 |
| Virology | |
| Oropharyngeal/nasopharyngeal swab | 1, 3, 5, 8, 11, 15 (±1 day) |
| Saliva | 1, 3, 5, 8, 11, 15 (±1 day) |

### Slide 4
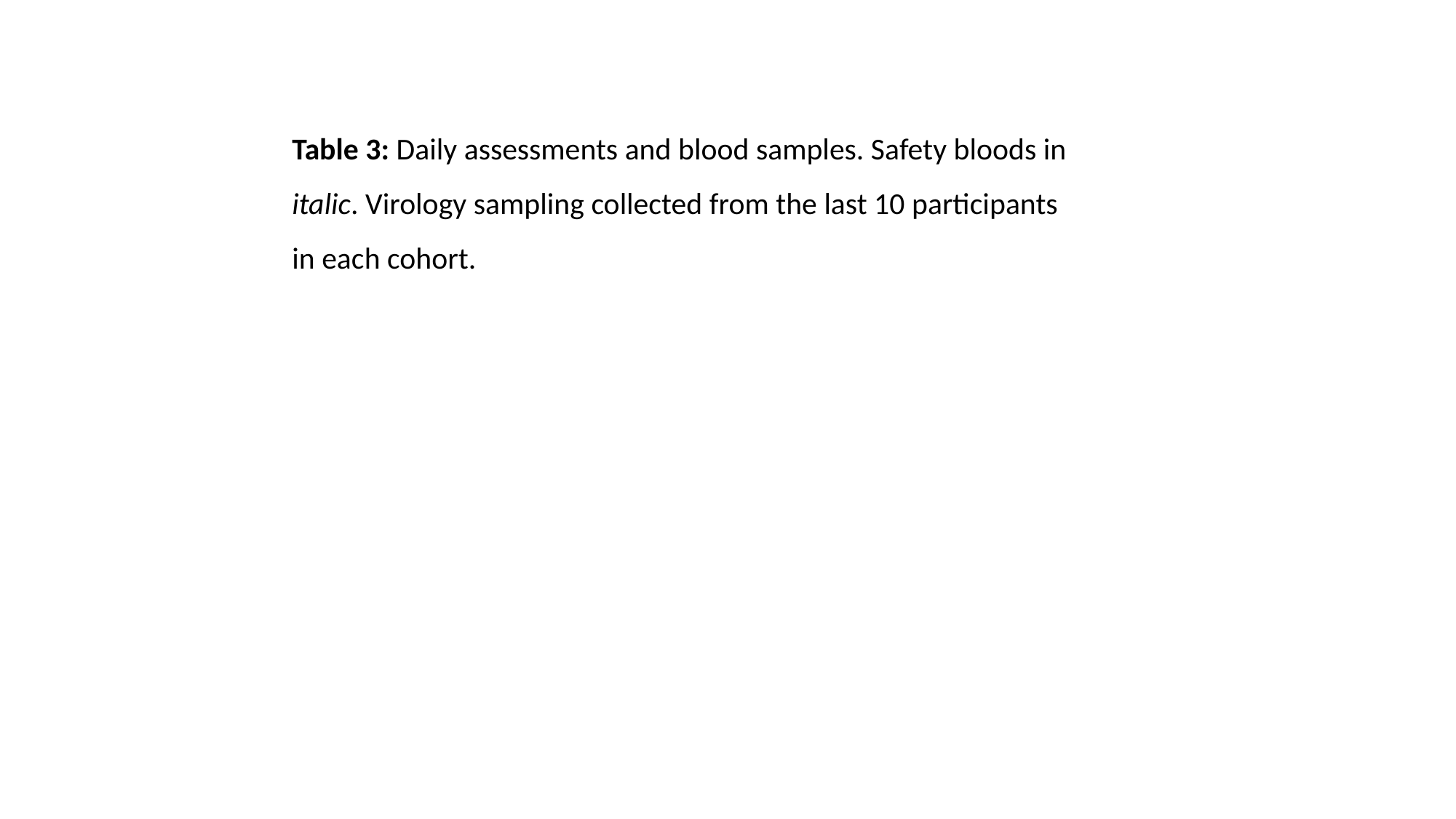

Table 3: Daily assessments and blood samples. Safety bloods in italic. Virology sampling collected from the last 10 participants in each cohort.

### Slide 5
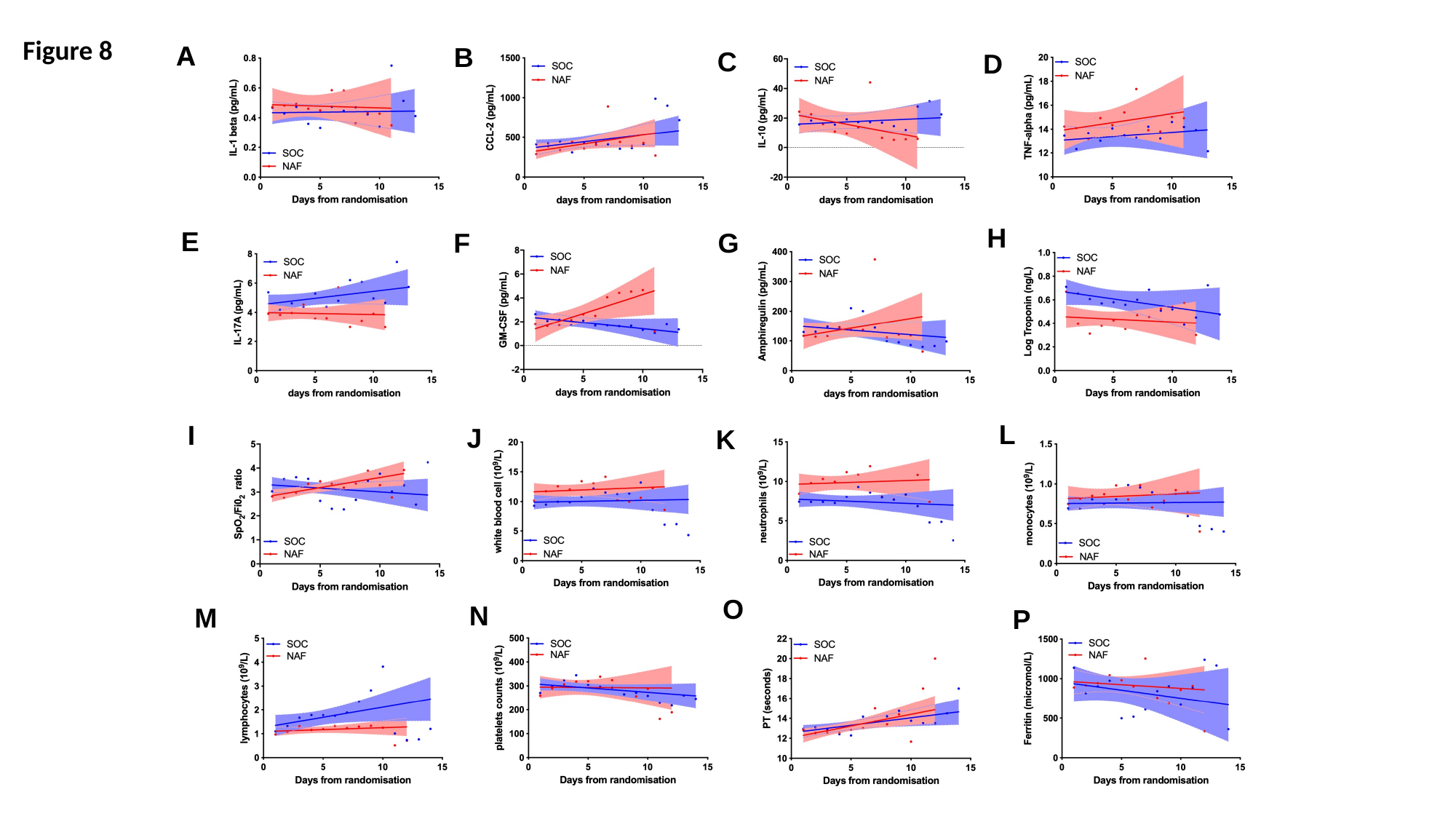

Figure 8
A
A
B
C
D
H
E
F
G
L
I
J
K
O
N
M
P

### Slide 6
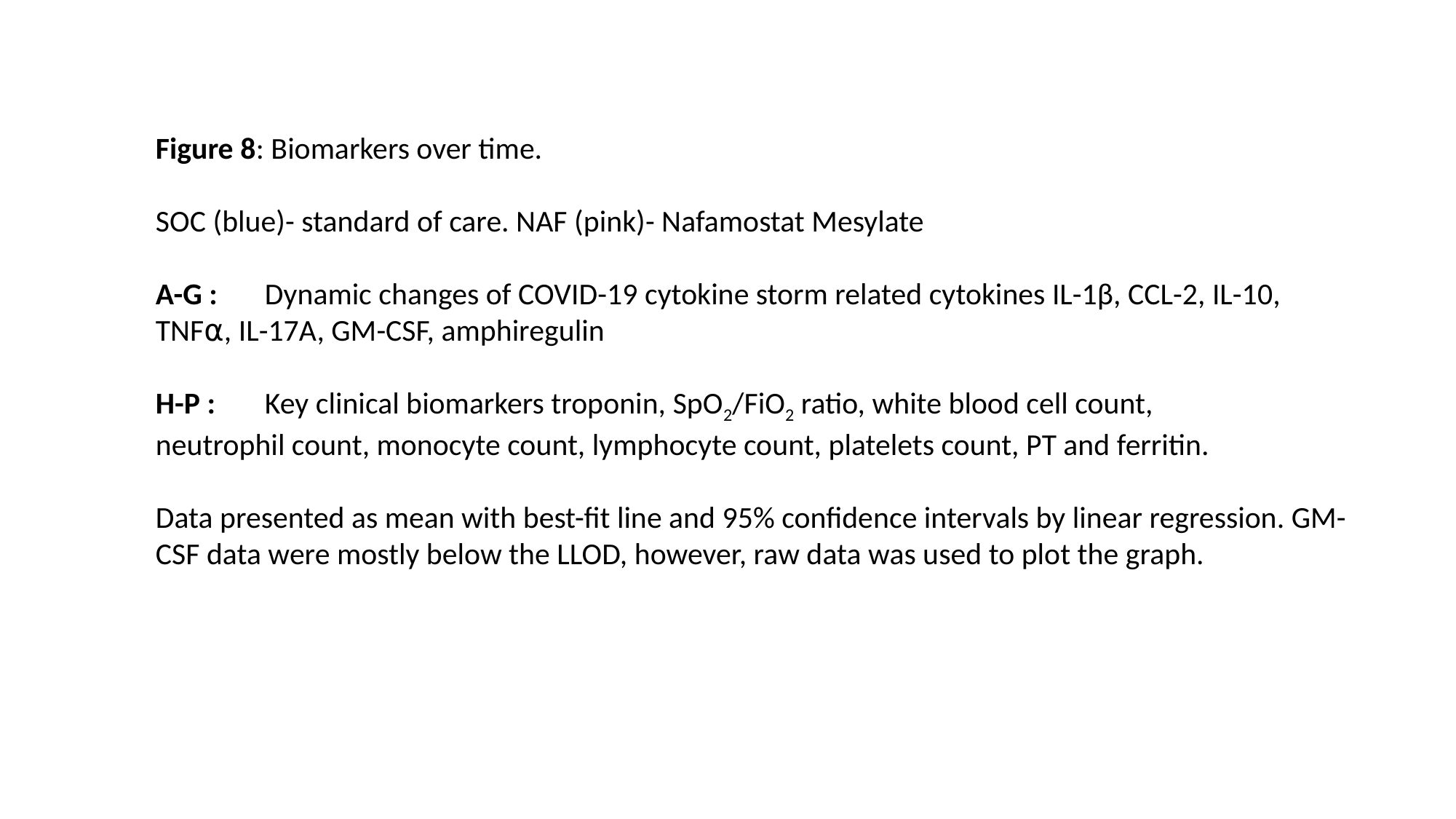

Figure 8: Biomarkers over time.
SOC (blue)- standard of care. NAF (pink)- Nafamostat Mesylate
A-G : 	Dynamic changes of COVID-19 cytokine storm related cytokines IL-1β, CCL-2, IL-10, 	TNF⍺, IL-17A, GM-CSF, amphiregulin
H-P : 	Key clinical biomarkers troponin, SpO_2_/FiO_2_ ratio, white blood cell count, 	neutrophil count, monocyte count, lymphocyte count, platelets count, PT and ferritin.
Data presented as mean with best-fit line and 95% confidence intervals by linear regression. GM-CSF data were mostly below the LLOD, however, raw data was used to plot the graph.

### Slide 7
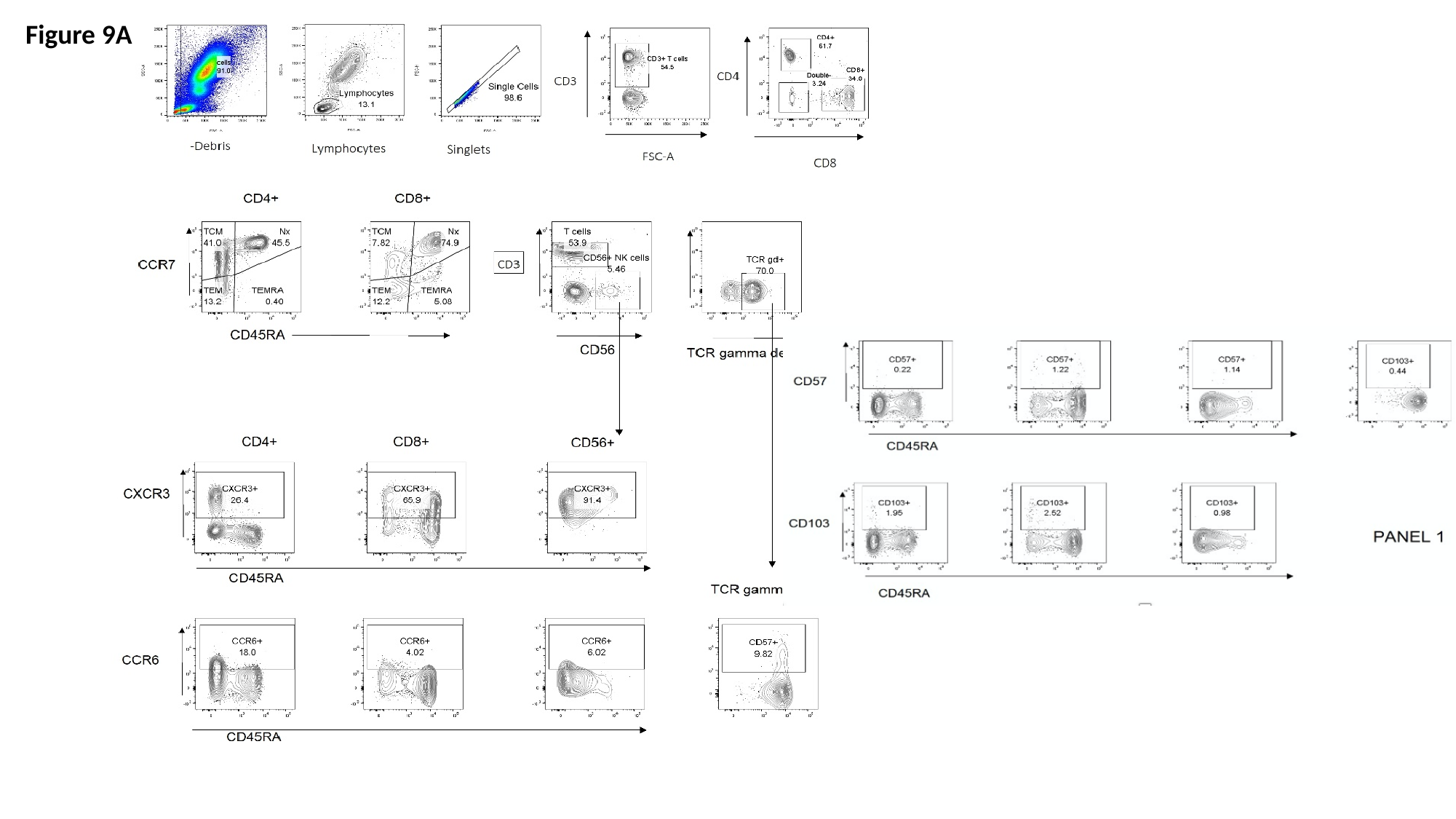

Figure 9A

### Slide 8
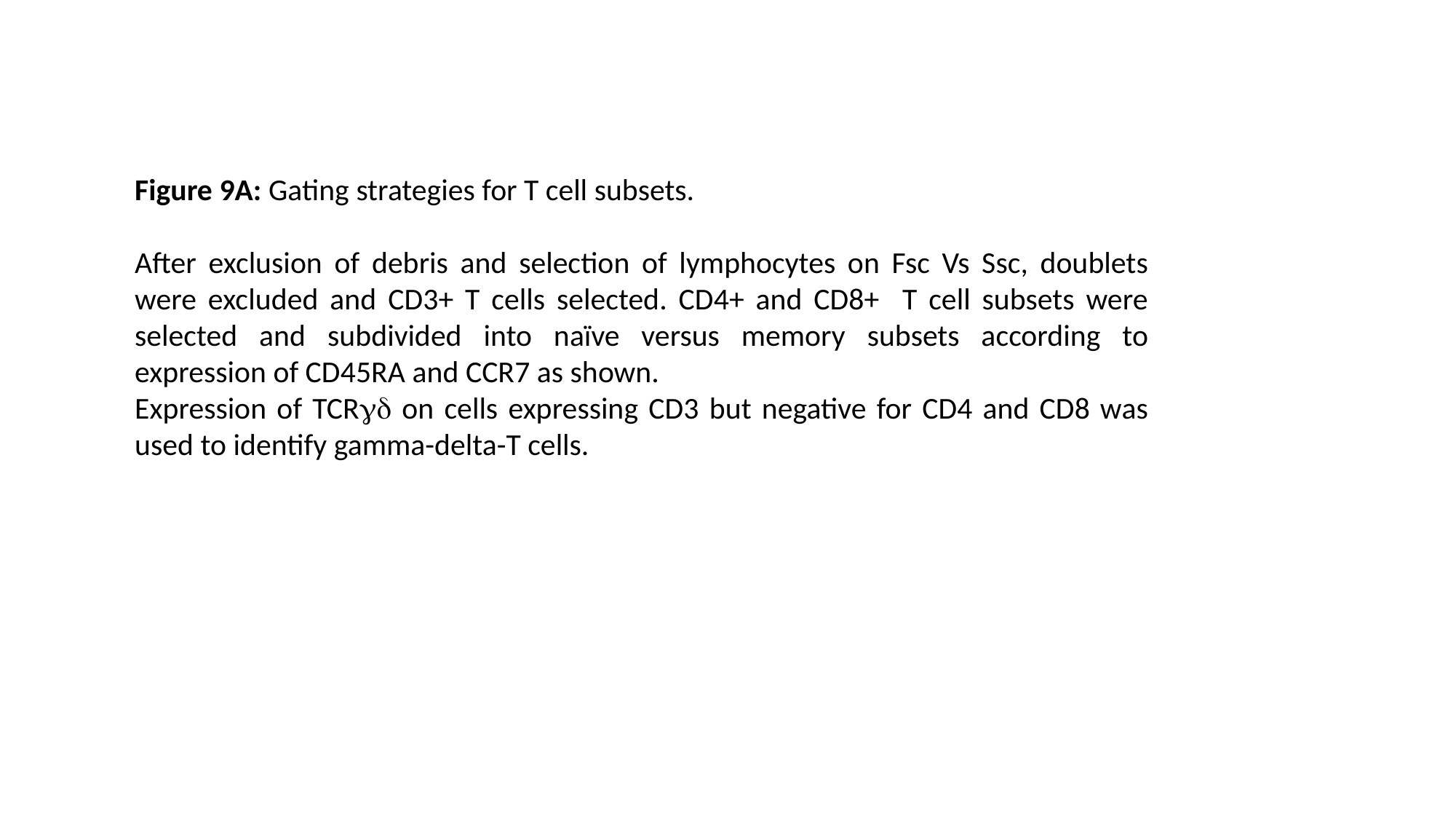

Figure 9A: Gating strategies for T cell subsets.
After exclusion of debris and selection of lymphocytes on Fsc Vs Ssc, doublets were excluded and CD3+ T cells selected. CD4+ and CD8+ T cell subsets were selected and subdivided into naïve versus memory subsets according to expression of CD45RA and CCR7 as shown.
Expression of TCRγδ on cells expressing CD3 but negative for CD4 and CD8 was used to identify gamma-delta-T cells.

### Slide 9
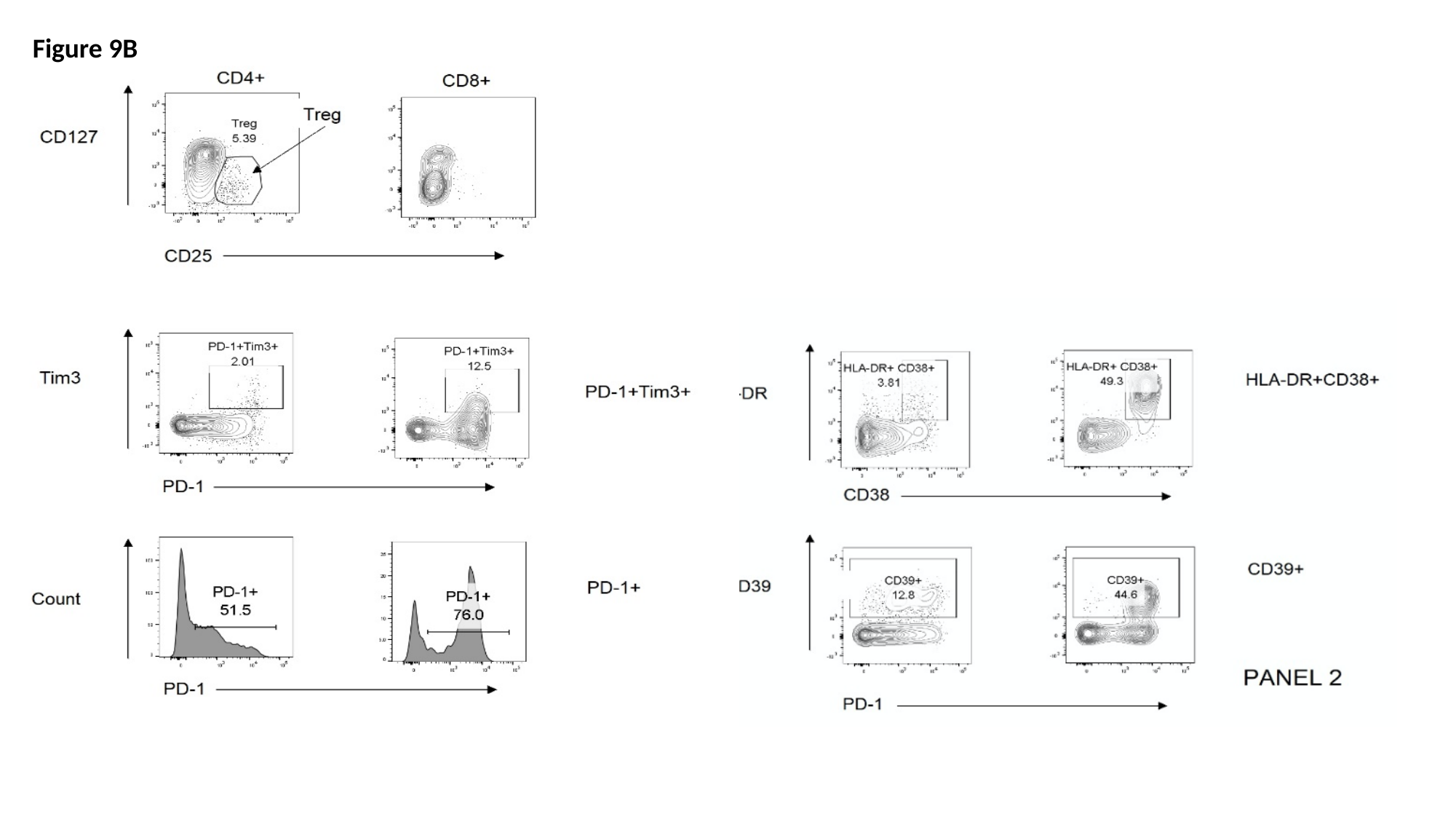

Figure 9B

### Slide 10
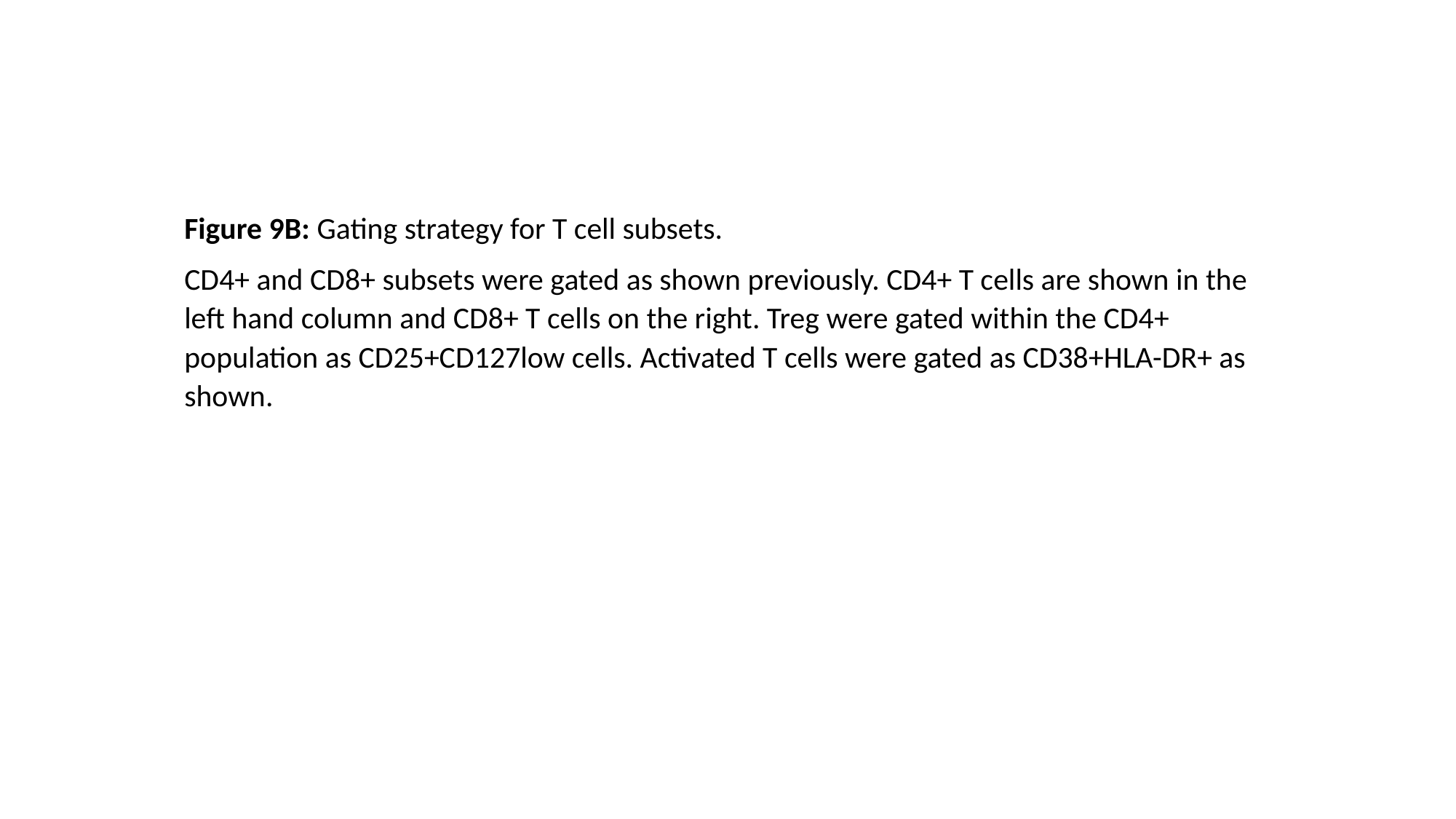

Figure 9B: Gating strategy for T cell subsets.
CD4+ and CD8+ subsets were gated as shown previously. CD4+ T cells are shown in the left hand column and CD8+ T cells on the right. Treg were gated within the CD4+ population as CD25+CD127low cells. Activated T cells were gated as CD38+HLA-DR+ as shown.

### Slide 11
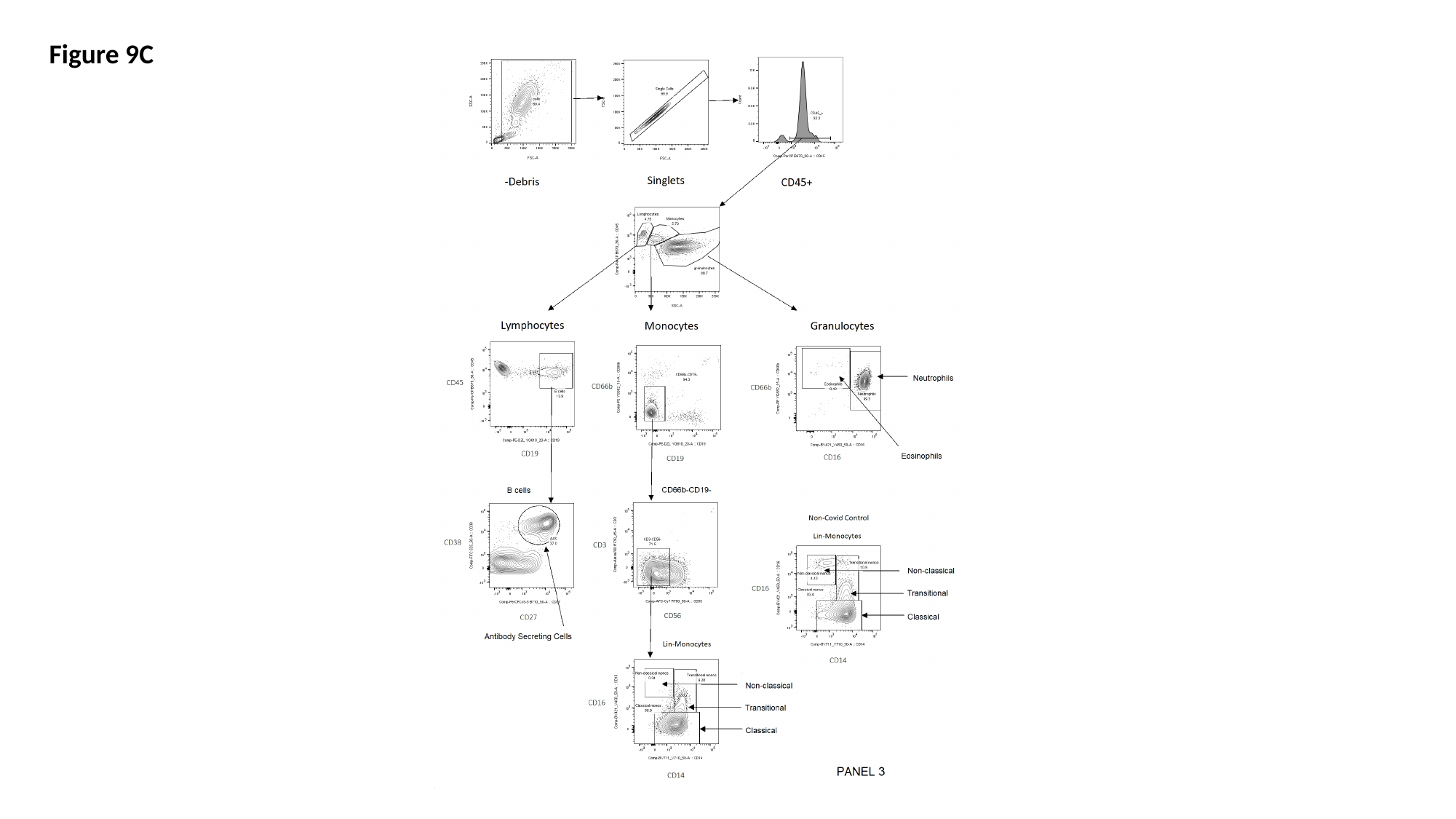

Figure 9C

### Slide 12
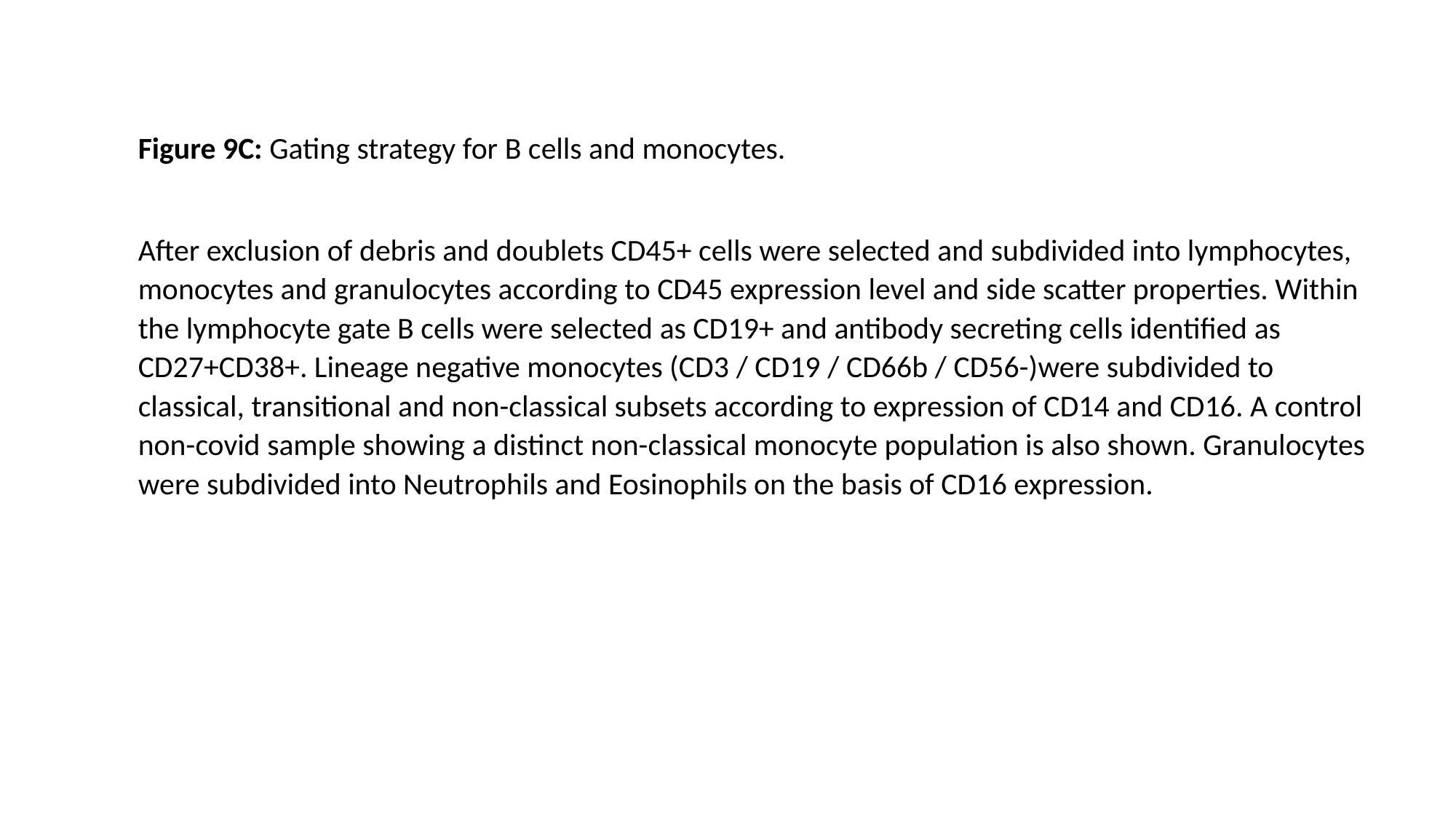

Figure 9C: Gating strategy for B cells and monocytes.
After exclusion of debris and doublets CD45+ cells were selected and subdivided into lymphocytes, monocytes and granulocytes according to CD45 expression level and side scatter properties. Within the lymphocyte gate B cells were selected as CD19+ and antibody secreting cells identified as CD27+CD38+. Lineage negative monocytes (CD3 / CD19 / CD66b / CD56-)were subdivided to classical, transitional and non-classical subsets according to expression of CD14 and CD16. A control non-covid sample showing a distinct non-classical monocyte population is also shown. Granulocytes were subdivided into Neutrophils and Eosinophils on the basis of CD16 expression.

### Slide 13
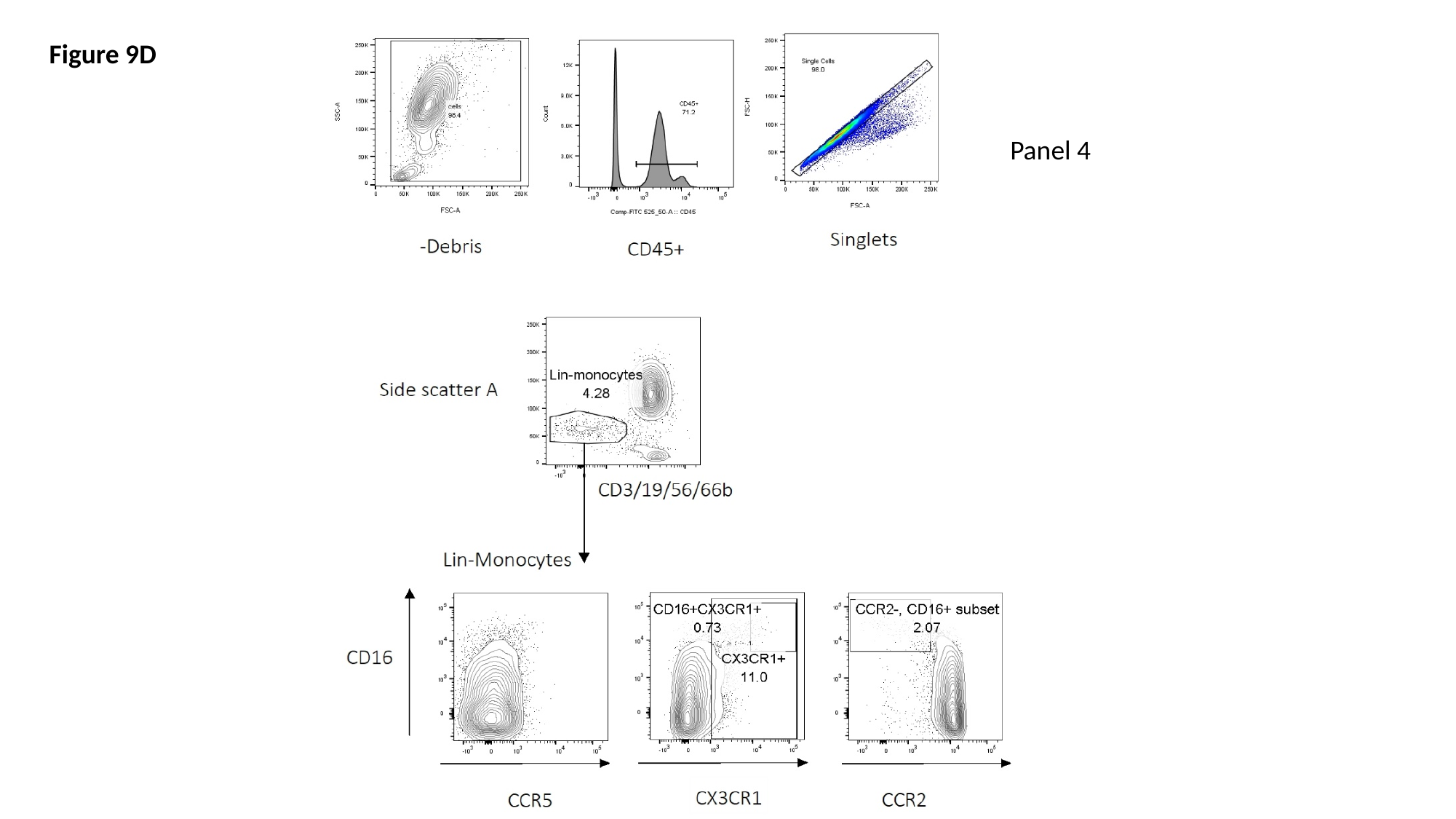

Figure 9D
Panel 4

### Slide 14
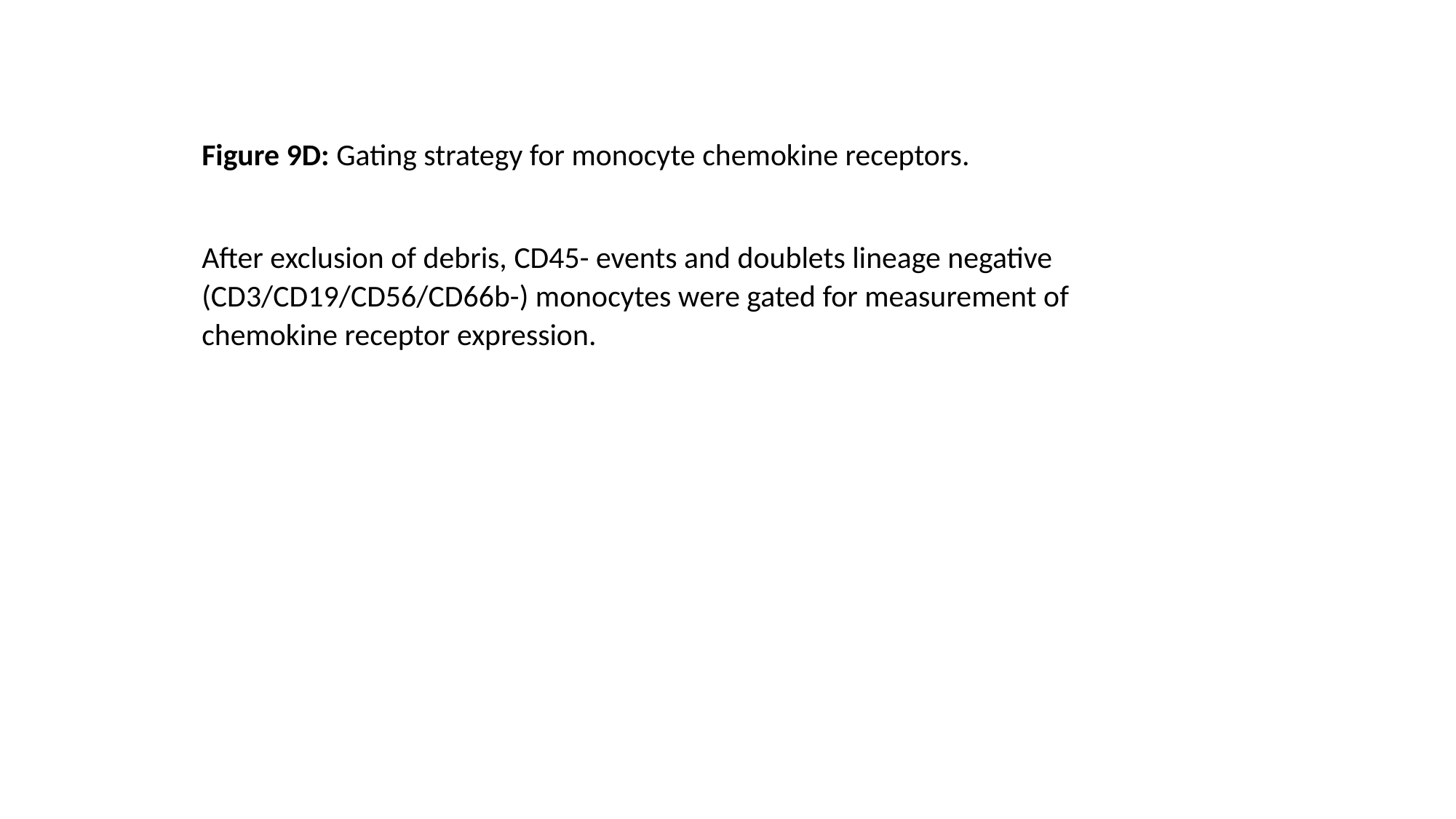

Figure 9D: Gating strategy for monocyte chemokine receptors.
After exclusion of debris, CD45- events and doublets lineage negative (CD3/CD19/CD56/CD66b-) monocytes were gated for measurement of chemokine receptor expression.

### Slide 15
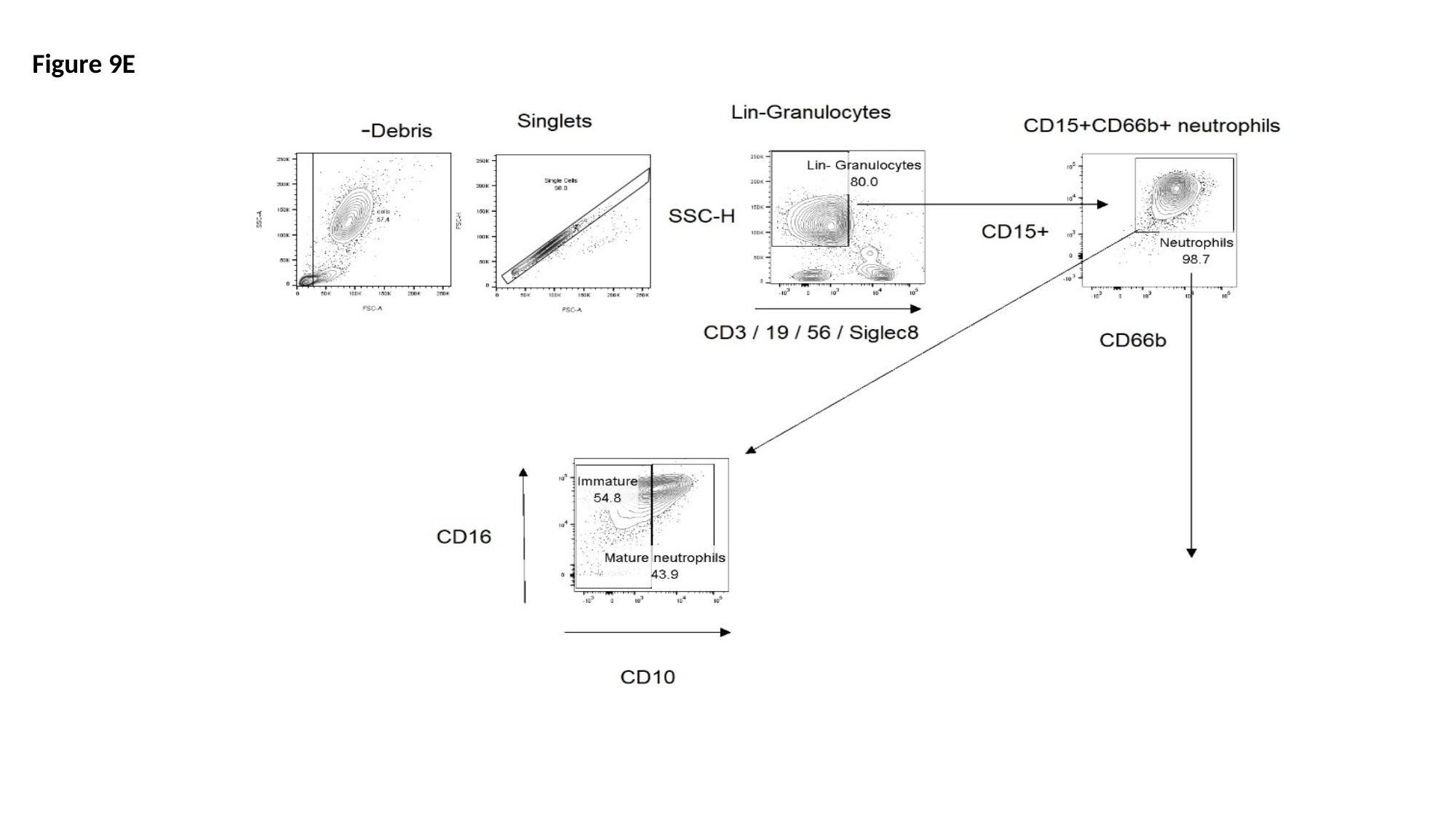

Figure 9E

### Slide 16
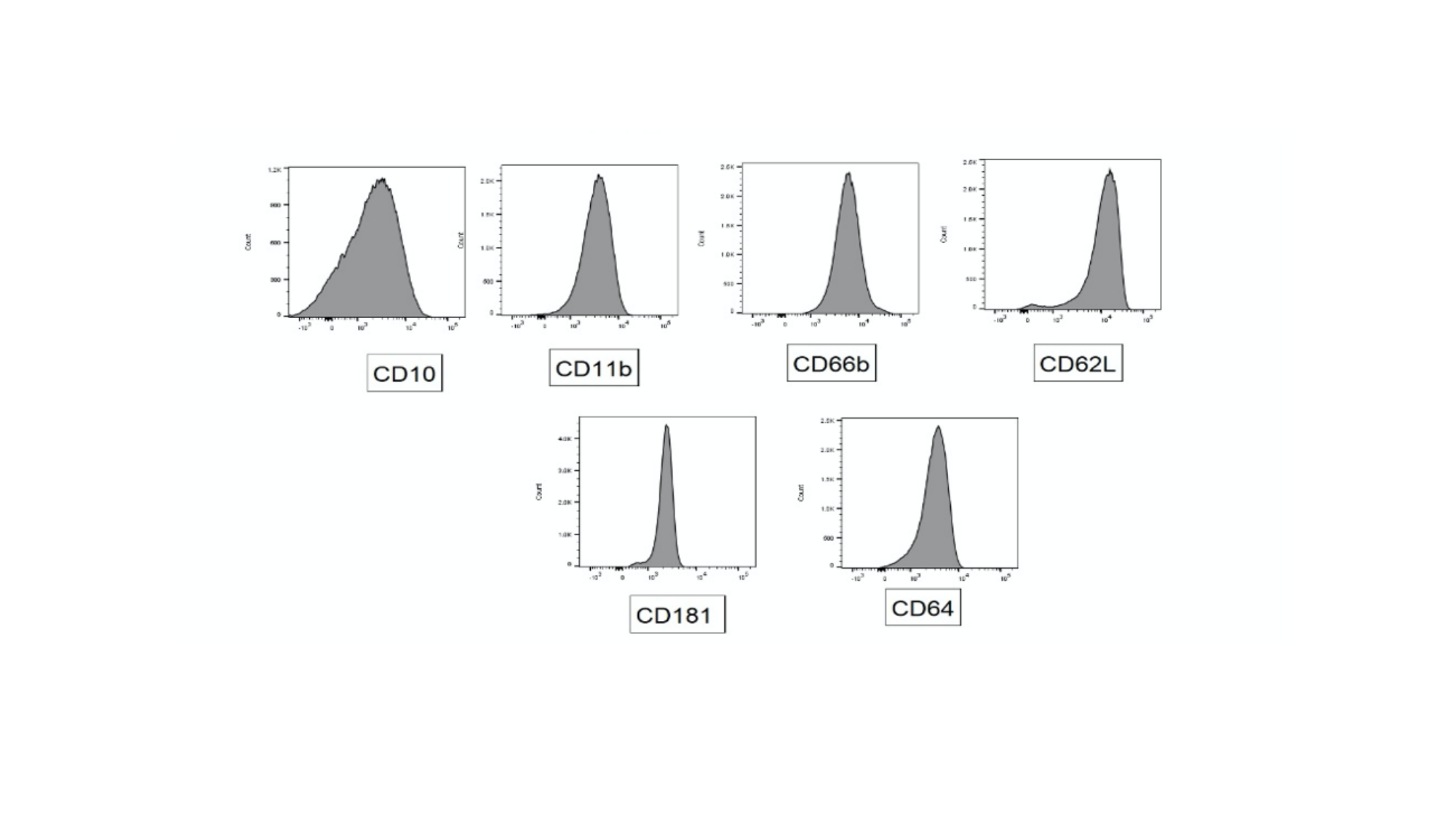

### Slide 17
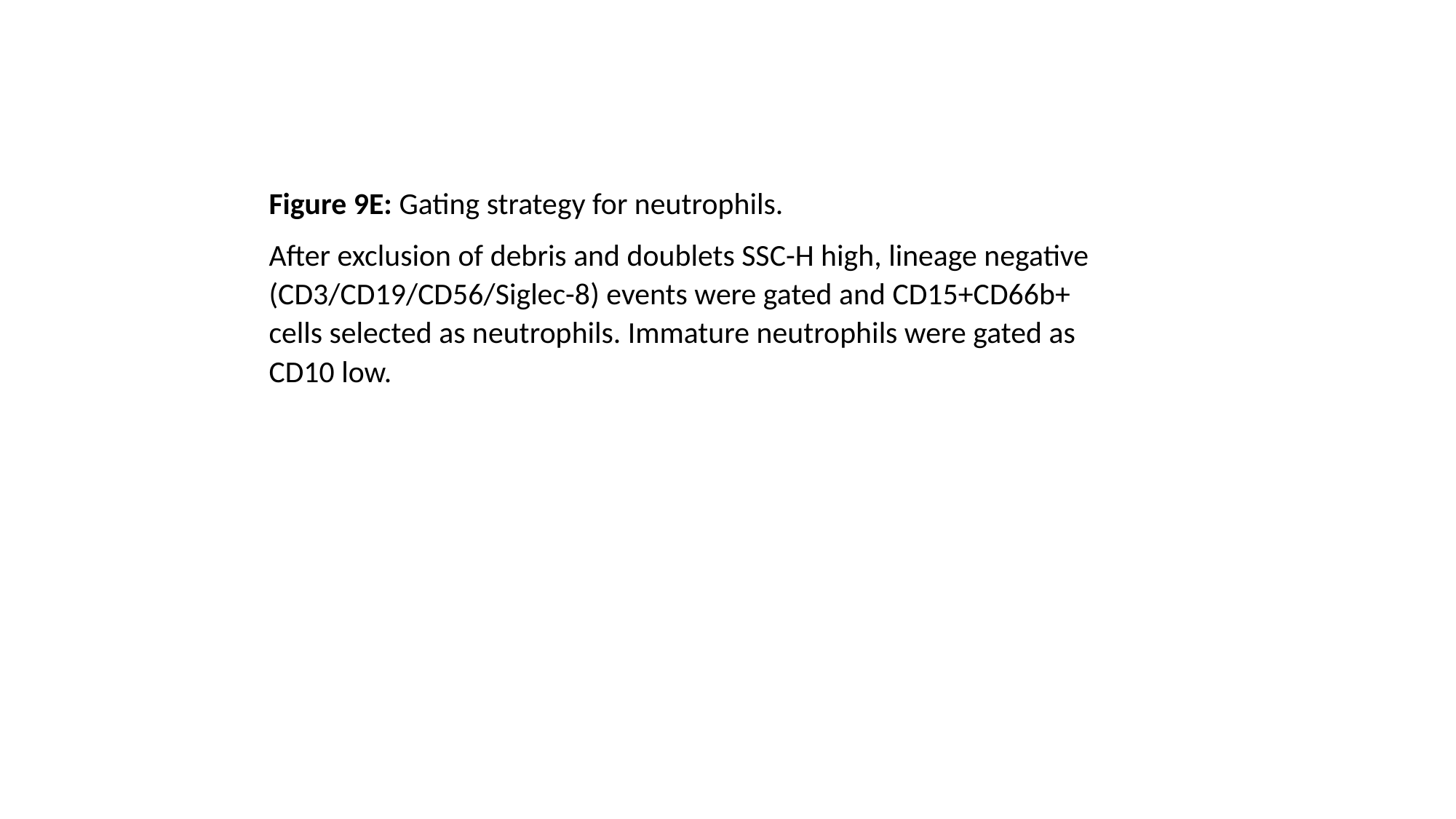

Figure 9E: Gating strategy for neutrophils.
After exclusion of debris and doublets SSC-H high, lineage negative (CD3/CD19/CD56/Siglec-8) events were gated and CD15+CD66b+ cells selected as neutrophils. Immature neutrophils were gated as CD10 low.

### Slide 18
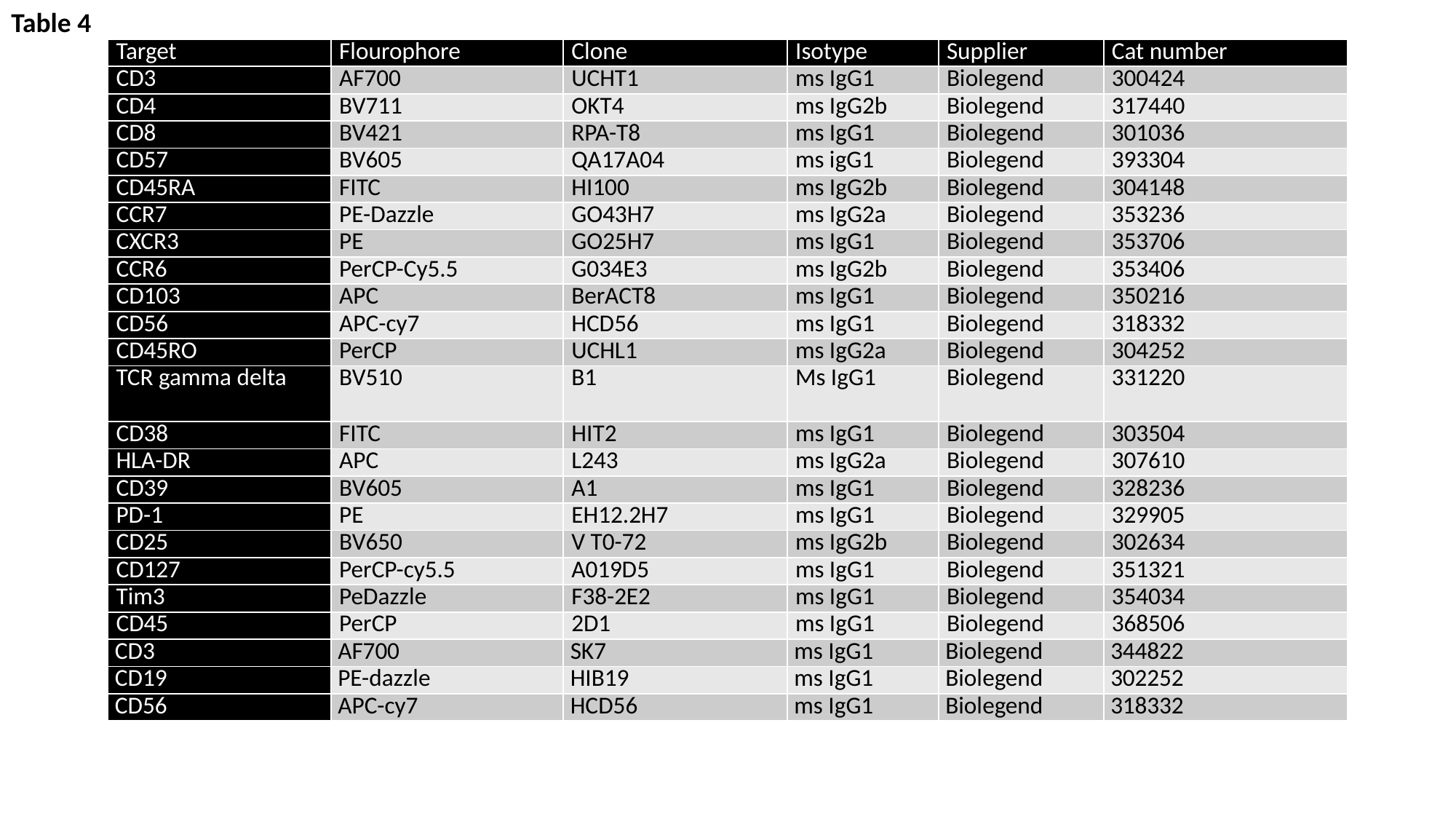

Table 4
| Target | Flourophore | Clone | Isotype | Supplier | Cat number |
| --- | --- | --- | --- | --- | --- |
| CD3 | AF700 | UCHT1 | ms IgG1 | Biolegend | 300424 |
| CD4 | BV711 | OKT4 | ms IgG2b | Biolegend | 317440 |
| CD8 | BV421 | RPA-T8 | ms IgG1 | Biolegend | 301036 |
| CD57 | BV605 | QA17A04 | ms igG1 | Biolegend | 393304 |
| CD45RA | FITC | HI100 | ms IgG2b | Biolegend | 304148 |
| CCR7 | PE-Dazzle | GO43H7 | ms IgG2a | Biolegend | 353236 |
| CXCR3 | PE | GO25H7 | ms IgG1 | Biolegend | 353706 |
| CCR6 | PerCP-Cy5.5 | G034E3 | ms IgG2b | Biolegend | 353406 |
| CD103 | APC | BerACT8 | ms IgG1 | Biolegend | 350216 |
| CD56 | APC-cy7 | HCD56 | ms IgG1 | Biolegend | 318332 |
| CD45RO | PerCP | UCHL1 | ms IgG2a | Biolegend | 304252 |
| TCR gamma delta | BV510 | B1 | Ms IgG1 | Biolegend | 331220 |
| CD38 | FITC | HIT2 | ms IgG1 | Biolegend | 303504 |
| HLA-DR | APC | L243 | ms IgG2a | Biolegend | 307610 |
| CD39 | BV605 | A1 | ms IgG1 | Biolegend | 328236 |
| PD-1 | PE | EH12.2H7 | ms IgG1 | Biolegend | 329905 |
| CD25 | BV650 | V T0-72 | ms IgG2b | Biolegend | 302634 |
| CD127 | PerCP-cy5.5 | A019D5 | ms IgG1 | Biolegend | 351321 |
| Tim3 | PeDazzle | F38-2E2 | ms IgG1 | Biolegend | 354034 |
| CD45 | PerCP | 2D1 | ms IgG1 | Biolegend | 368506 |
| CD3 | AF700 | SK7 | ms IgG1 | Biolegend | 344822 |
| CD19 | PE-dazzle | HIB19 | ms IgG1 | Biolegend | 302252 |
| CD56 | APC-cy7 | HCD56 | ms IgG1 | Biolegend | 318332 |

### Slide 19
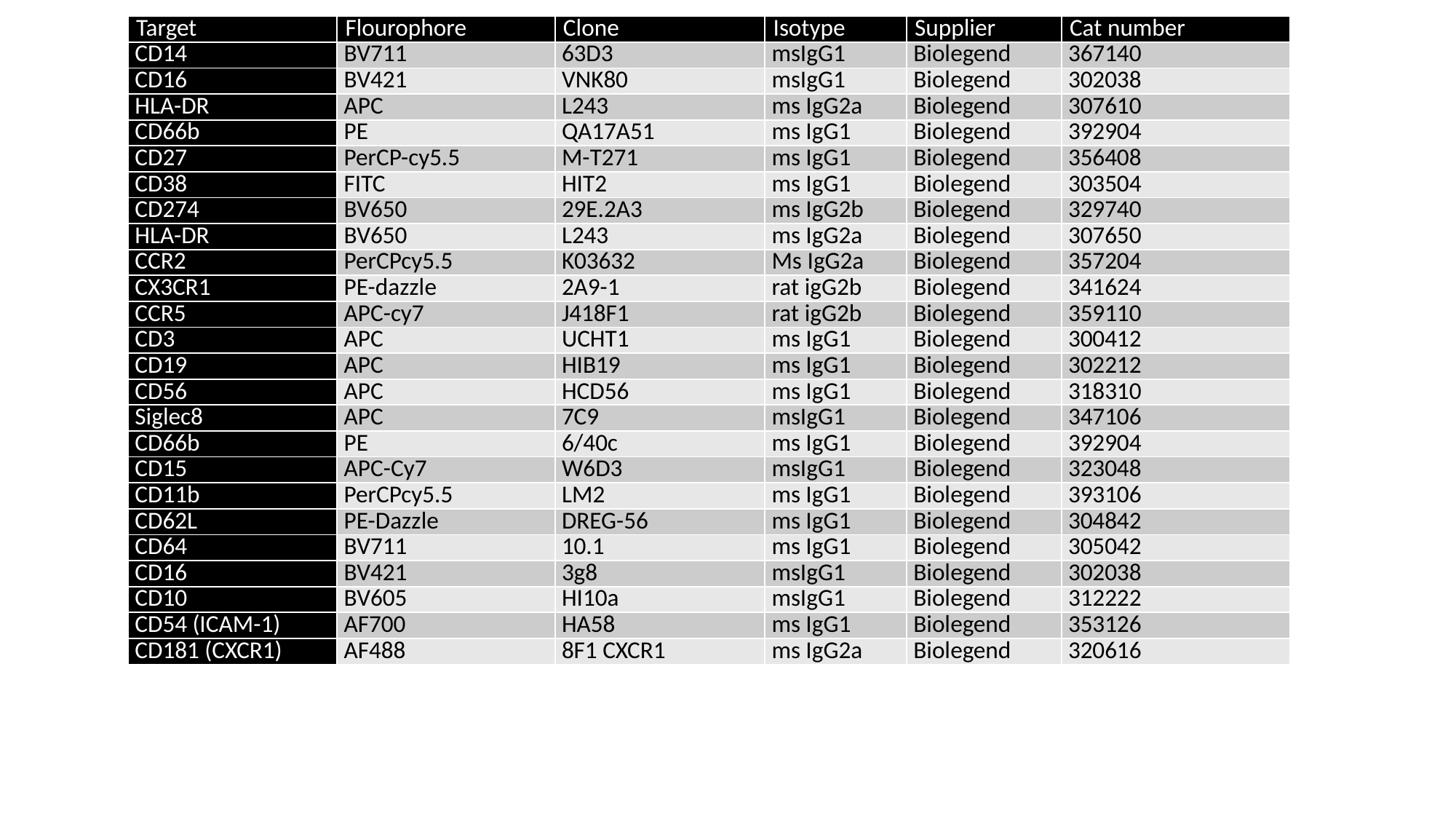

| Target | Flourophore | Clone | Isotype | Supplier | Cat number |
| --- | --- | --- | --- | --- | --- |
| CD14 | BV711 | 63D3 | msIgG1 | Biolegend | 367140 |
| CD16 | BV421 | VNK80 | msIgG1 | Biolegend | 302038 |
| HLA-DR | APC | L243 | ms IgG2a | Biolegend | 307610 |
| CD66b | PE | QA17A51 | ms IgG1 | Biolegend | 392904 |
| CD27 | PerCP-cy5.5 | M-T271 | ms IgG1 | Biolegend | 356408 |
| CD38 | FITC | HIT2 | ms IgG1 | Biolegend | 303504 |
| CD274 | BV650 | 29E.2A3 | ms IgG2b | Biolegend | 329740 |
| HLA-DR | BV650 | L243 | ms IgG2a | Biolegend | 307650 |
| CCR2 | PerCPcy5.5 | K03632 | Ms IgG2a | Biolegend | 357204 |
| CX3CR1 | PE-dazzle | 2A9-1 | rat igG2b | Biolegend | 341624 |
| CCR5 | APC-cy7 | J418F1 | rat igG2b | Biolegend | 359110 |
| CD3 | APC | UCHT1 | ms IgG1 | Biolegend | 300412 |
| CD19 | APC | HIB19 | ms IgG1 | Biolegend | 302212 |
| CD56 | APC | HCD56 | ms IgG1 | Biolegend | 318310 |
| Siglec8 | APC | 7C9 | msIgG1 | Biolegend | 347106 |
| CD66b | PE | 6/40c | ms IgG1 | Biolegend | 392904 |
| CD15 | APC-Cy7 | W6D3 | msIgG1 | Biolegend | 323048 |
| CD11b | PerCPcy5.5 | LM2 | ms IgG1 | Biolegend | 393106 |
| CD62L | PE-Dazzle | DREG-56 | ms IgG1 | Biolegend | 304842 |
| CD64 | BV711 | 10.1 | ms IgG1 | Biolegend | 305042 |
| CD16 | BV421 | 3g8 | msIgG1 | Biolegend | 302038 |
| CD10 | BV605 | HI10a | msIgG1 | Biolegend | 312222 |
| CD54 (ICAM-1) | AF700 | HA58 | ms IgG1 | Biolegend | 353126 |
| CD181 (CXCR1) | AF488 | 8F1 CXCR1 | ms IgG2a | Biolegend | 320616 |

### Slide 20
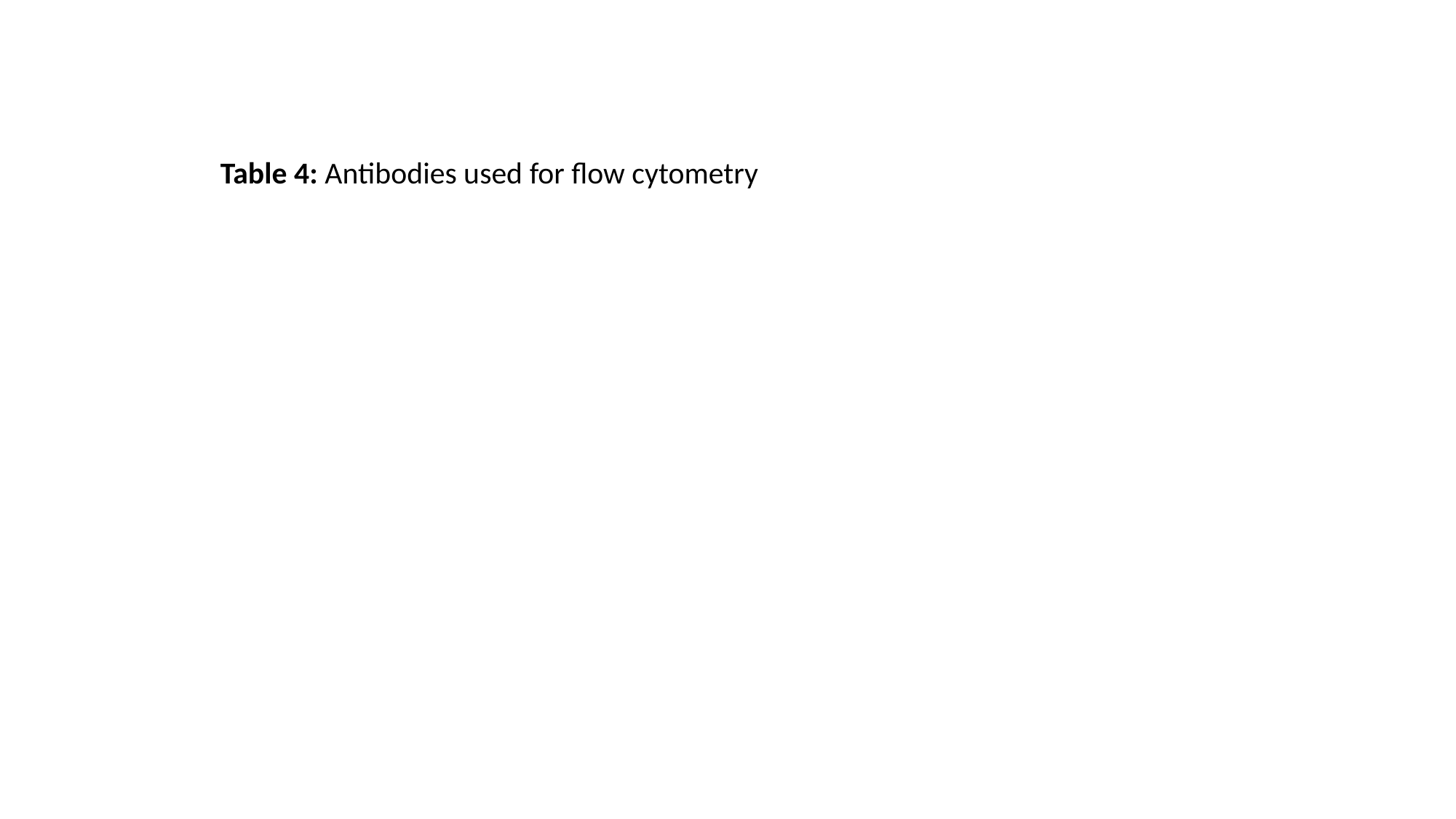

Table 4: Antibodies used for flow cytometry

### Slide 21
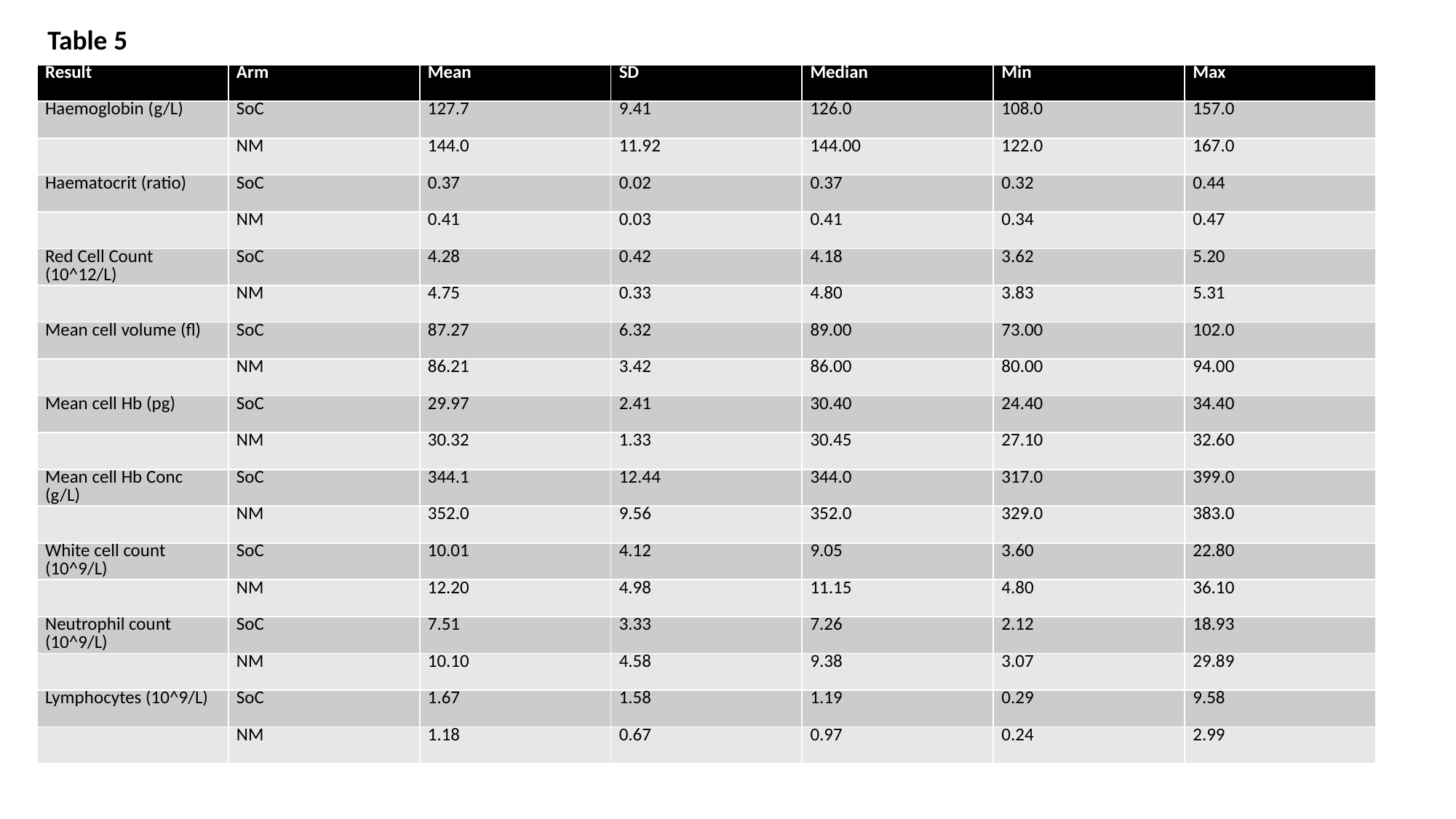

Table 5
| Result | Arm | Mean | SD | Median | Min | Max |
| --- | --- | --- | --- | --- | --- | --- |
| Haemoglobin (g/L) | SoC | 127.7 | 9.41 | 126.0 | 108.0 | 157.0 |
| | NM | 144.0 | 11.92 | 144.00 | 122.0 | 167.0 |
| Haematocrit (ratio) | SoC | 0.37 | 0.02 | 0.37 | 0.32 | 0.44 |
| | NM | 0.41 | 0.03 | 0.41 | 0.34 | 0.47 |
| Red Cell Count (10^12/L) | SoC | 4.28 | 0.42 | 4.18 | 3.62 | 5.20 |
| | NM | 4.75 | 0.33 | 4.80 | 3.83 | 5.31 |
| Mean cell volume (fl) | SoC | 87.27 | 6.32 | 89.00 | 73.00 | 102.0 |
| | NM | 86.21 | 3.42 | 86.00 | 80.00 | 94.00 |
| Mean cell Hb (pg) | SoC | 29.97 | 2.41 | 30.40 | 24.40 | 34.40 |
| | NM | 30.32 | 1.33 | 30.45 | 27.10 | 32.60 |
| Mean cell Hb Conc (g/L) | SoC | 344.1 | 12.44 | 344.0 | 317.0 | 399.0 |
| | NM | 352.0 | 9.56 | 352.0 | 329.0 | 383.0 |
| White cell count (10^9/L) | SoC | 10.01 | 4.12 | 9.05 | 3.60 | 22.80 |
| | NM | 12.20 | 4.98 | 11.15 | 4.80 | 36.10 |
| Neutrophil count (10^9/L) | SoC | 7.51 | 3.33 | 7.26 | 2.12 | 18.93 |
| | NM | 10.10 | 4.58 | 9.38 | 3.07 | 29.89 |
| Lymphocytes (10^9/L) | SoC | 1.67 | 1.58 | 1.19 | 0.29 | 9.58 |
| | NM | 1.18 | 0.67 | 0.97 | 0.24 | 2.99 |

### Slide 22
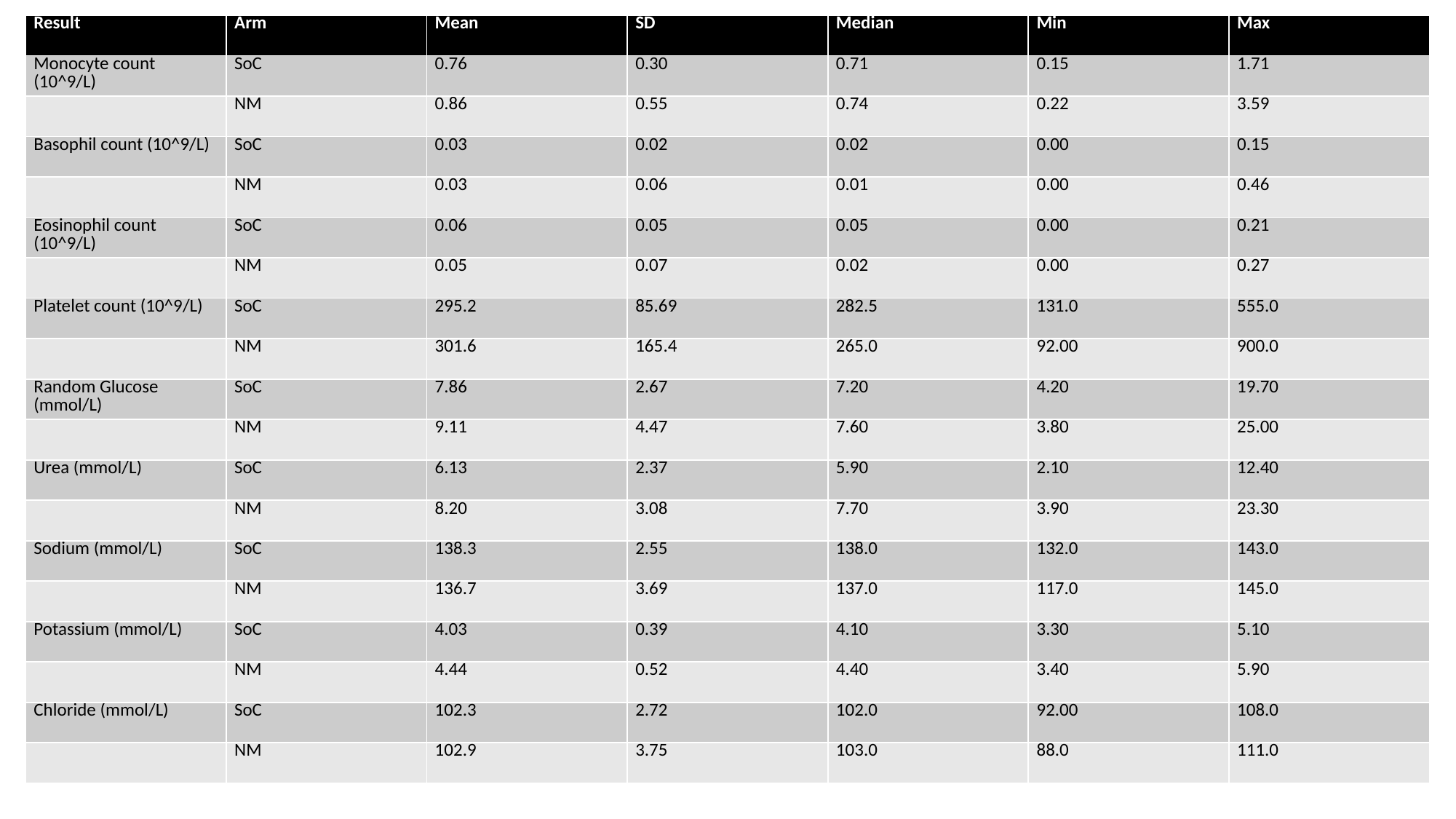

| Result | Arm | Mean | SD | Median | Min | Max |
| --- | --- | --- | --- | --- | --- | --- |
| Monocyte count (10^9/L) | SoC | 0.76 | 0.30 | 0.71 | 0.15 | 1.71 |
| | NM | 0.86 | 0.55 | 0.74 | 0.22 | 3.59 |
| Basophil count (10^9/L) | SoC | 0.03 | 0.02 | 0.02 | 0.00 | 0.15 |
| | NM | 0.03 | 0.06 | 0.01 | 0.00 | 0.46 |
| Eosinophil count (10^9/L) | SoC | 0.06 | 0.05 | 0.05 | 0.00 | 0.21 |
| | NM | 0.05 | 0.07 | 0.02 | 0.00 | 0.27 |
| Platelet count (10^9/L) | SoC | 295.2 | 85.69 | 282.5 | 131.0 | 555.0 |
| | NM | 301.6 | 165.4 | 265.0 | 92.00 | 900.0 |
| Random Glucose (mmol/L) | SoC | 7.86 | 2.67 | 7.20 | 4.20 | 19.70 |
| | NM | 9.11 | 4.47 | 7.60 | 3.80 | 25.00 |
| Urea (mmol/L) | SoC | 6.13 | 2.37 | 5.90 | 2.10 | 12.40 |
| | NM | 8.20 | 3.08 | 7.70 | 3.90 | 23.30 |
| Sodium (mmol/L) | SoC | 138.3 | 2.55 | 138.0 | 132.0 | 143.0 |
| | NM | 136.7 | 3.69 | 137.0 | 117.0 | 145.0 |
| Potassium (mmol/L) | SoC | 4.03 | 0.39 | 4.10 | 3.30 | 5.10 |
| | NM | 4.44 | 0.52 | 4.40 | 3.40 | 5.90 |
| Chloride (mmol/L) | SoC | 102.3 | 2.72 | 102.0 | 92.00 | 108.0 |
| | NM | 102.9 | 3.75 | 103.0 | 88.0 | 111.0 |

### Slide 23
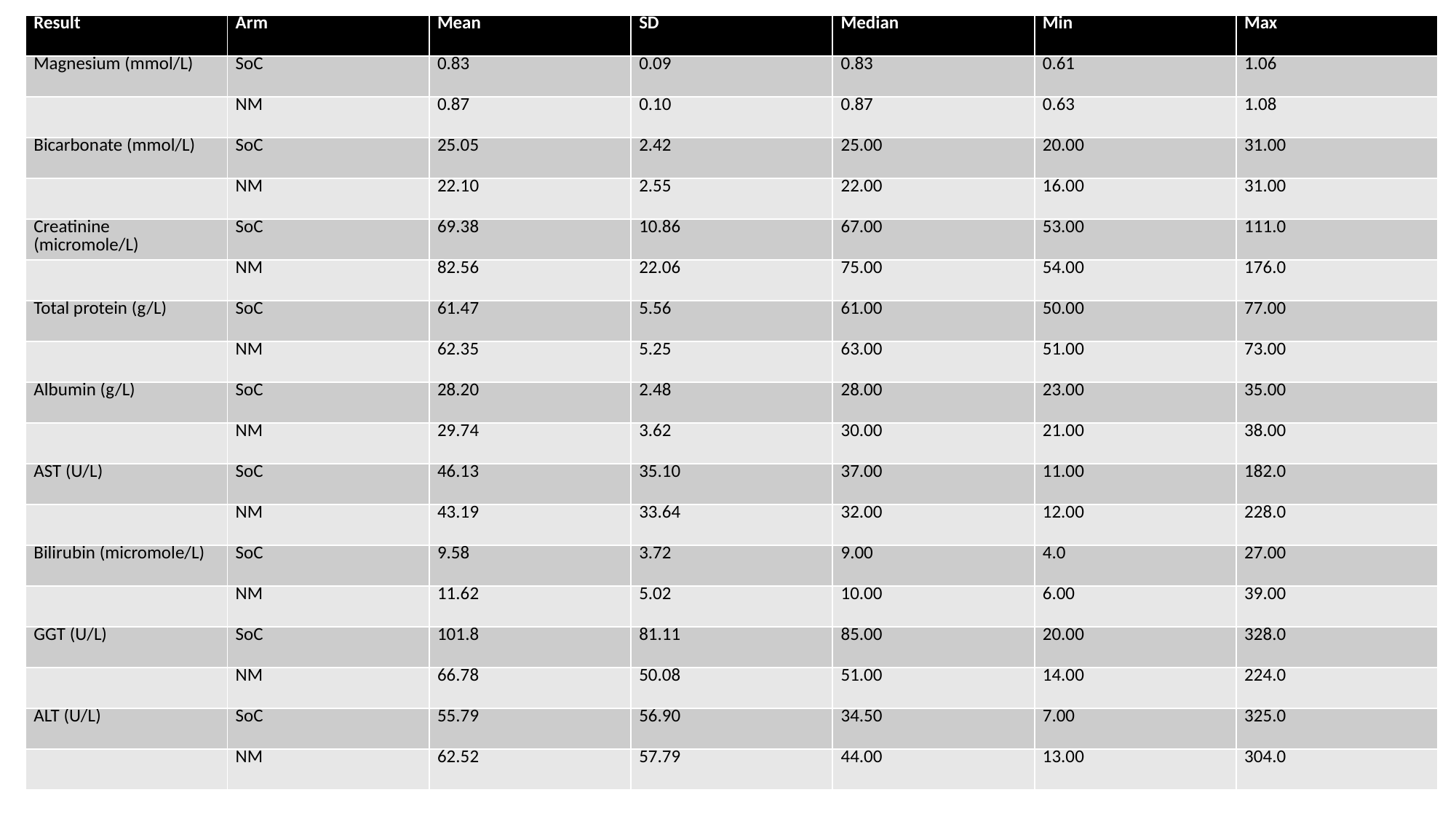

| Result | Arm | Mean | SD | Median | Min | Max |
| --- | --- | --- | --- | --- | --- | --- |
| Magnesium (mmol/L) | SoC | 0.83 | 0.09 | 0.83 | 0.61 | 1.06 |
| | NM | 0.87 | 0.10 | 0.87 | 0.63 | 1.08 |
| Bicarbonate (mmol/L) | SoC | 25.05 | 2.42 | 25.00 | 20.00 | 31.00 |
| | NM | 22.10 | 2.55 | 22.00 | 16.00 | 31.00 |
| Creatinine (micromole/L) | SoC | 69.38 | 10.86 | 67.00 | 53.00 | 111.0 |
| | NM | 82.56 | 22.06 | 75.00 | 54.00 | 176.0 |
| Total protein (g/L) | SoC | 61.47 | 5.56 | 61.00 | 50.00 | 77.00 |
| | NM | 62.35 | 5.25 | 63.00 | 51.00 | 73.00 |
| Albumin (g/L) | SoC | 28.20 | 2.48 | 28.00 | 23.00 | 35.00 |
| | NM | 29.74 | 3.62 | 30.00 | 21.00 | 38.00 |
| AST (U/L) | SoC | 46.13 | 35.10 | 37.00 | 11.00 | 182.0 |
| | NM | 43.19 | 33.64 | 32.00 | 12.00 | 228.0 |
| Bilirubin (micromole/L) | SoC | 9.58 | 3.72 | 9.00 | 4.0 | 27.00 |
| | NM | 11.62 | 5.02 | 10.00 | 6.00 | 39.00 |
| GGT (U/L) | SoC | 101.8 | 81.11 | 85.00 | 20.00 | 328.0 |
| | NM | 66.78 | 50.08 | 51.00 | 14.00 | 224.0 |
| ALT (U/L) | SoC | 55.79 | 56.90 | 34.50 | 7.00 | 325.0 |
| | NM | 62.52 | 57.79 | 44.00 | 13.00 | 304.0 |

### Slide 24
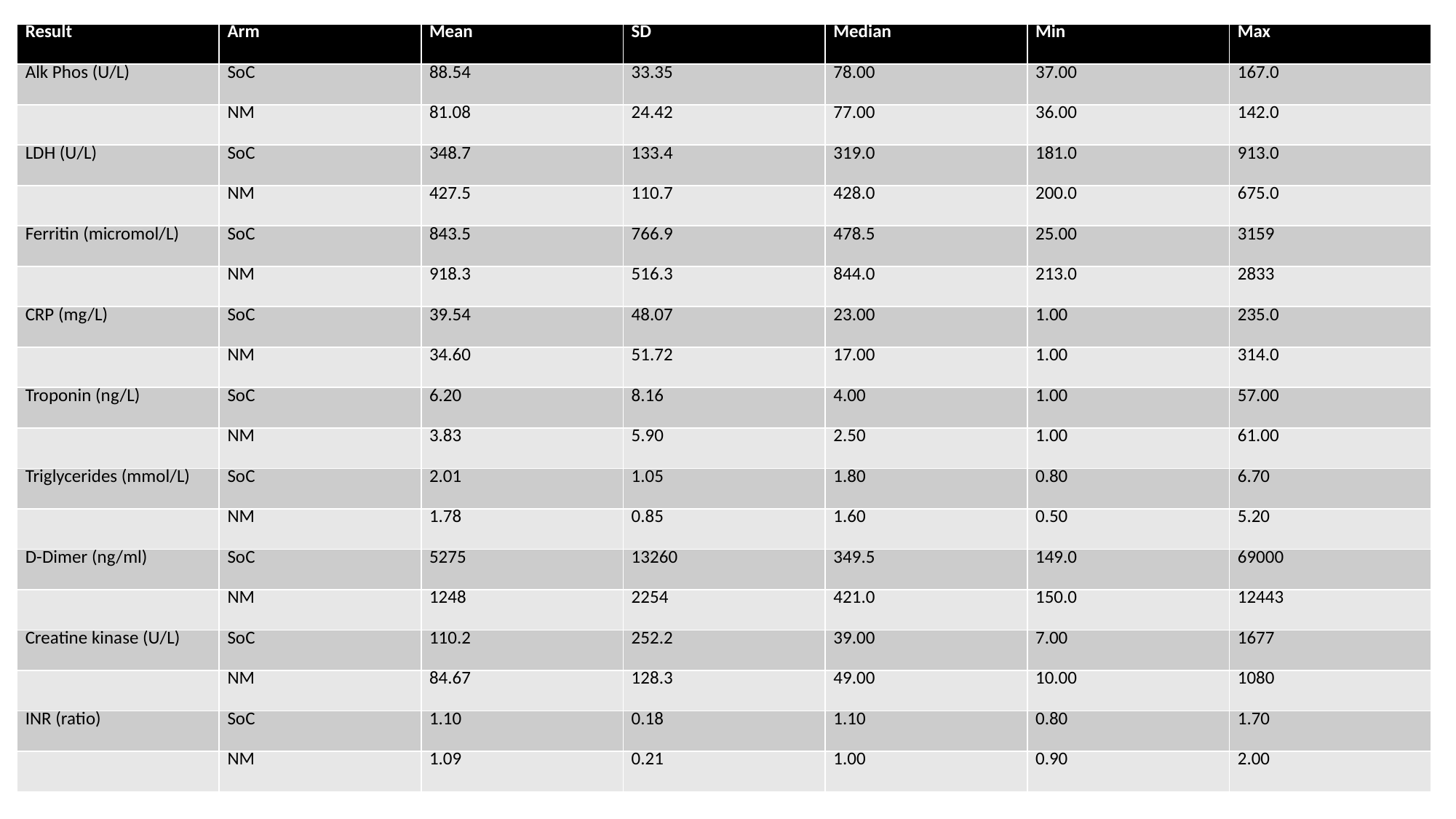

| Result | Arm | Mean | SD | Median | Min | Max |
| --- | --- | --- | --- | --- | --- | --- |
| Alk Phos (U/L) | SoC | 88.54 | 33.35 | 78.00 | 37.00 | 167.0 |
| | NM | 81.08 | 24.42 | 77.00 | 36.00 | 142.0 |
| LDH (U/L) | SoC | 348.7 | 133.4 | 319.0 | 181.0 | 913.0 |
| | NM | 427.5 | 110.7 | 428.0 | 200.0 | 675.0 |
| Ferritin (micromol/L) | SoC | 843.5 | 766.9 | 478.5 | 25.00 | 3159 |
| | NM | 918.3 | 516.3 | 844.0 | 213.0 | 2833 |
| CRP (mg/L) | SoC | 39.54 | 48.07 | 23.00 | 1.00 | 235.0 |
| | NM | 34.60 | 51.72 | 17.00 | 1.00 | 314.0 |
| Troponin (ng/L) | SoC | 6.20 | 8.16 | 4.00 | 1.00 | 57.00 |
| | NM | 3.83 | 5.90 | 2.50 | 1.00 | 61.00 |
| Triglycerides (mmol/L) | SoC | 2.01 | 1.05 | 1.80 | 0.80 | 6.70 |
| | NM | 1.78 | 0.85 | 1.60 | 0.50 | 5.20 |
| D-Dimer (ng/ml) | SoC | 5275 | 13260 | 349.5 | 149.0 | 69000 |
| | NM | 1248 | 2254 | 421.0 | 150.0 | 12443 |
| Creatine kinase (U/L) | SoC | 110.2 | 252.2 | 39.00 | 7.00 | 1677 |
| | NM | 84.67 | 128.3 | 49.00 | 10.00 | 1080 |
| INR (ratio) | SoC | 1.10 | 0.18 | 1.10 | 0.80 | 1.70 |
| | NM | 1.09 | 0.21 | 1.00 | 0.90 | 2.00 |

### Slide 25
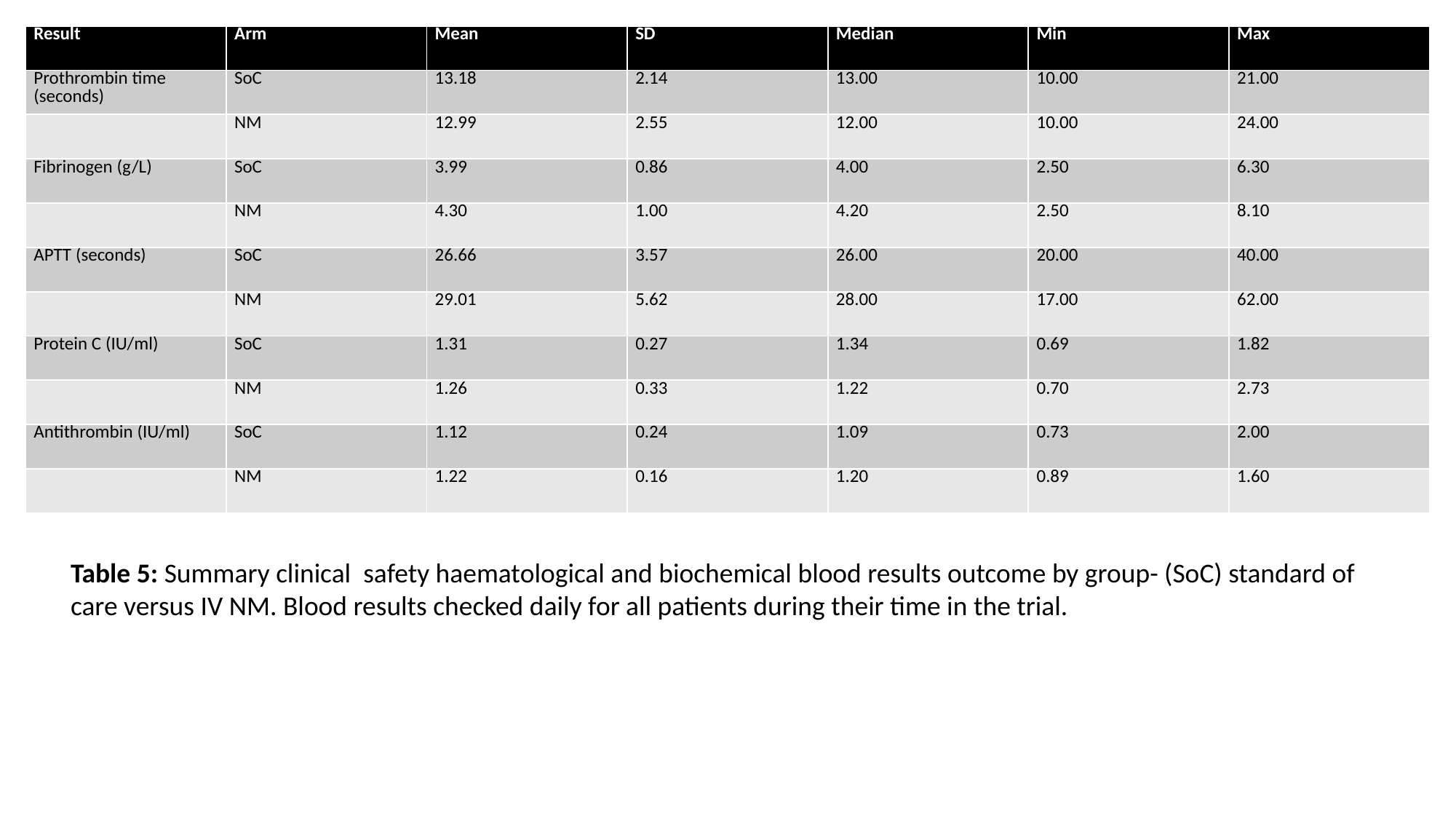

| Result | Arm | Mean | SD | Median | Min | Max |
| --- | --- | --- | --- | --- | --- | --- |
| Prothrombin time (seconds) | SoC | 13.18 | 2.14 | 13.00 | 10.00 | 21.00 |
| | NM | 12.99 | 2.55 | 12.00 | 10.00 | 24.00 |
| Fibrinogen (g/L) | SoC | 3.99 | 0.86 | 4.00 | 2.50 | 6.30 |
| | NM | 4.30 | 1.00 | 4.20 | 2.50 | 8.10 |
| APTT (seconds) | SoC | 26.66 | 3.57 | 26.00 | 20.00 | 40.00 |
| | NM | 29.01 | 5.62 | 28.00 | 17.00 | 62.00 |
| Protein C (IU/ml) | SoC | 1.31 | 0.27 | 1.34 | 0.69 | 1.82 |
| | NM | 1.26 | 0.33 | 1.22 | 0.70 | 2.73 |
| Antithrombin (IU/ml) | SoC | 1.12 | 0.24 | 1.09 | 0.73 | 2.00 |
| | NM | 1.22 | 0.16 | 1.20 | 0.89 | 1.60 |
Table 5: Summary clinical safety haematological and biochemical blood results outcome by group- (SoC) standard of care versus IV NM. Blood results checked daily for all patients during their time in the trial.

### Slide 26
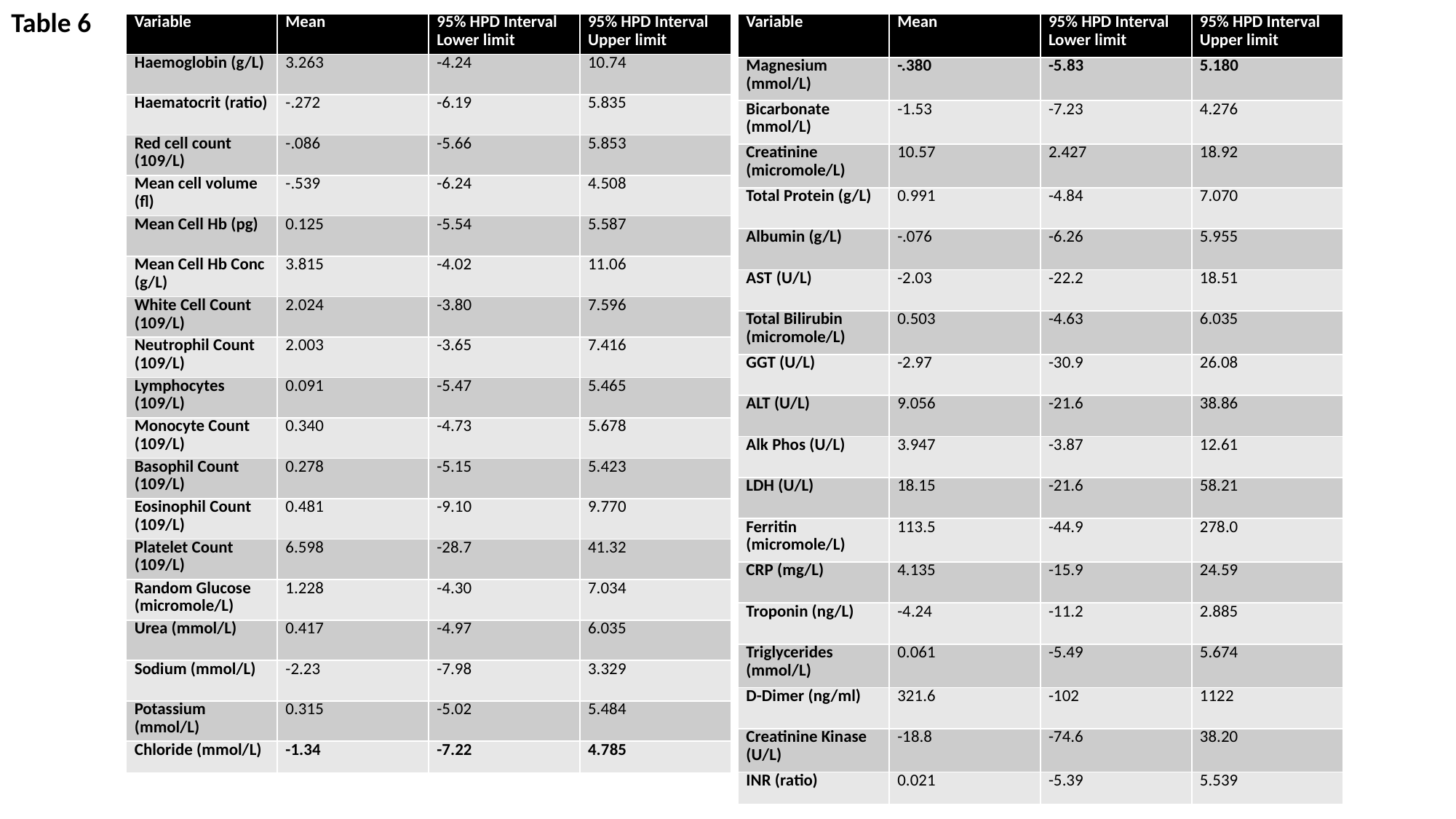

Table 6
| Variable | Mean | 95% HPD Interval Lower limit | 95% HPD Interval Upper limit |
| --- | --- | --- | --- |
| Magnesium (mmol/L) | -.380 | -5.83 | 5.180 |
| Bicarbonate (mmol/L) | -1.53 | -7.23 | 4.276 |
| Creatinine (micromole/L) | 10.57 | 2.427 | 18.92 |
| Total Protein (g/L) | 0.991 | -4.84 | 7.070 |
| Albumin (g/L) | -.076 | -6.26 | 5.955 |
| AST (U/L) | -2.03 | -22.2 | 18.51 |
| Total Bilirubin (micromole/L) | 0.503 | -4.63 | 6.035 |
| GGT (U/L) | -2.97 | -30.9 | 26.08 |
| ALT (U/L) | 9.056 | -21.6 | 38.86 |
| Alk Phos (U/L) | 3.947 | -3.87 | 12.61 |
| LDH (U/L) | 18.15 | -21.6 | 58.21 |
| Ferritin (micromole/L) | 113.5 | -44.9 | 278.0 |
| CRP (mg/L) | 4.135 | -15.9 | 24.59 |
| Troponin (ng/L) | -4.24 | -11.2 | 2.885 |
| Triglycerides (mmol/L) | 0.061 | -5.49 | 5.674 |
| D-Dimer (ng/ml) | 321.6 | -102 | 1122 |
| Creatinine Kinase (U/L) | -18.8 | -74.6 | 38.20 |
| INR (ratio) | 0.021 | -5.39 | 5.539 |
| Variable | Mean | 95% HPD Interval Lower limit | 95% HPD Interval Upper limit |
| --- | --- | --- | --- |
| Haemoglobin (g/L) | 3.263 | -4.24 | 10.74 |
| Haematocrit (ratio) | -.272 | -6.19 | 5.835 |
| Red cell count (109/L) | -.086 | -5.66 | 5.853 |
| Mean cell volume (fl) | -.539 | -6.24 | 4.508 |
| Mean Cell Hb (pg) | 0.125 | -5.54 | 5.587 |
| Mean Cell Hb Conc (g/L) | 3.815 | -4.02 | 11.06 |
| White Cell Count (109/L) | 2.024 | -3.80 | 7.596 |
| Neutrophil Count (109/L) | 2.003 | -3.65 | 7.416 |
| Lymphocytes (109/L) | 0.091 | -5.47 | 5.465 |
| Monocyte Count (109/L) | 0.340 | -4.73 | 5.678 |
| Basophil Count (109/L) | 0.278 | -5.15 | 5.423 |
| Eosinophil Count (109/L) | 0.481 | -9.10 | 9.770 |
| Platelet Count (109/L) | 6.598 | -28.7 | 41.32 |
| Random Glucose (micromole/L) | 1.228 | -4.30 | 7.034 |
| Urea (mmol/L) | 0.417 | -4.97 | 6.035 |
| Sodium (mmol/L) | -2.23 | -7.98 | 3.329 |
| Potassium (mmol/L) | 0.315 | -5.02 | 5.484 |
| Chloride (mmol/L) | -1.34 | -7.22 | 4.785 |

### Slide 27
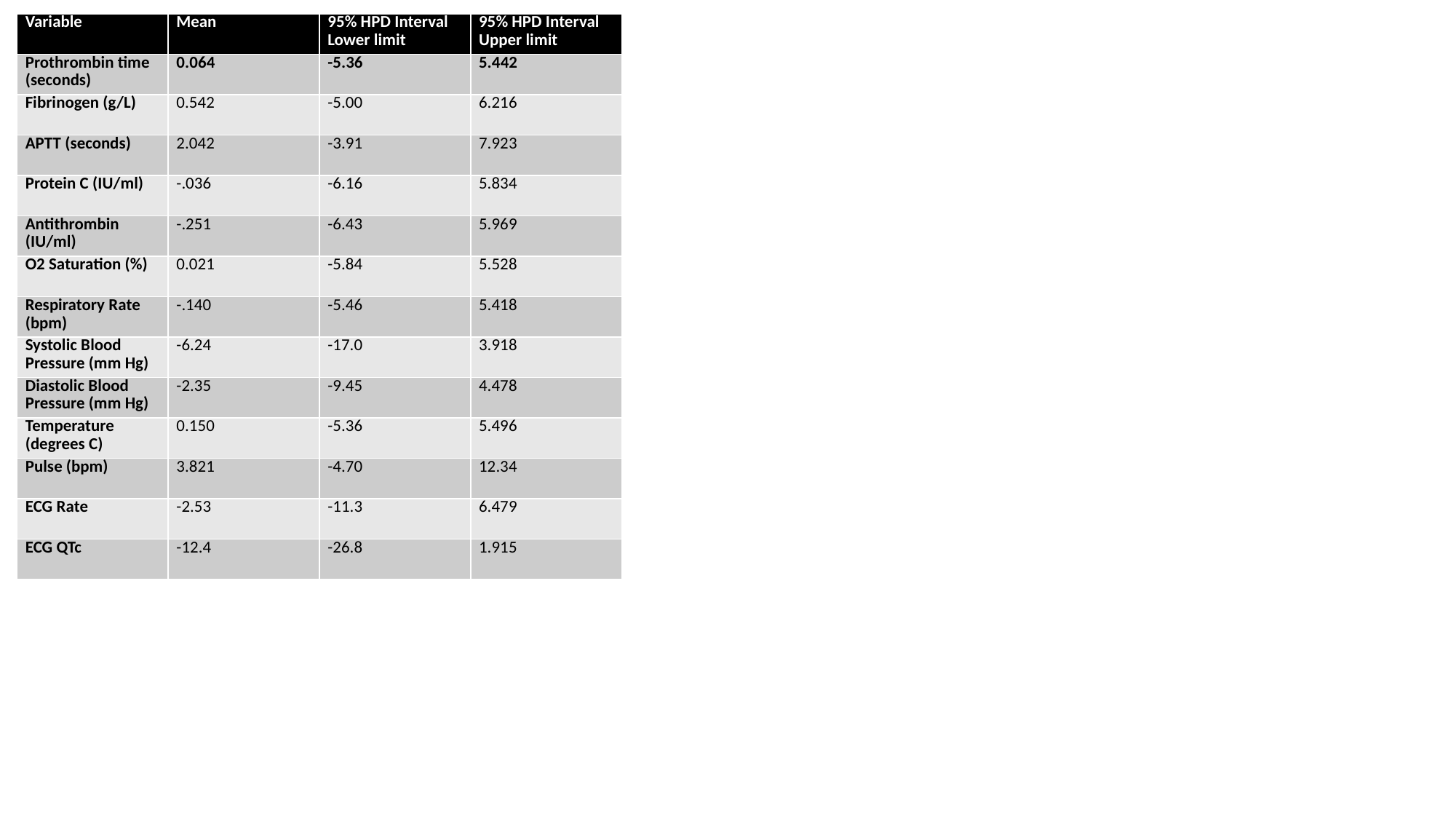

| Variable | Mean | 95% HPD Interval Lower limit | 95% HPD Interval Upper limit |
| --- | --- | --- | --- |
| Prothrombin time (seconds) | 0.064 | -5.36 | 5.442 |
| Fibrinogen (g/L) | 0.542 | -5.00 | 6.216 |
| APTT (seconds) | 2.042 | -3.91 | 7.923 |
| Protein C (IU/ml) | -.036 | -6.16 | 5.834 |
| Antithrombin (IU/ml) | -.251 | -6.43 | 5.969 |
| O2 Saturation (%) | 0.021 | -5.84 | 5.528 |
| Respiratory Rate (bpm) | -.140 | -5.46 | 5.418 |
| Systolic Blood Pressure (mm Hg) | -6.24 | -17.0 | 3.918 |
| Diastolic Blood Pressure (mm Hg) | -2.35 | -9.45 | 4.478 |
| Temperature (degrees C) | 0.150 | -5.36 | 5.496 |
| Pulse (bpm) | 3.821 | -4.70 | 12.34 |
| ECG Rate | -2.53 | -11.3 | 6.479 |
| ECG QTc | -12.4 | -26.8 | 1.915 |

### Slide 28
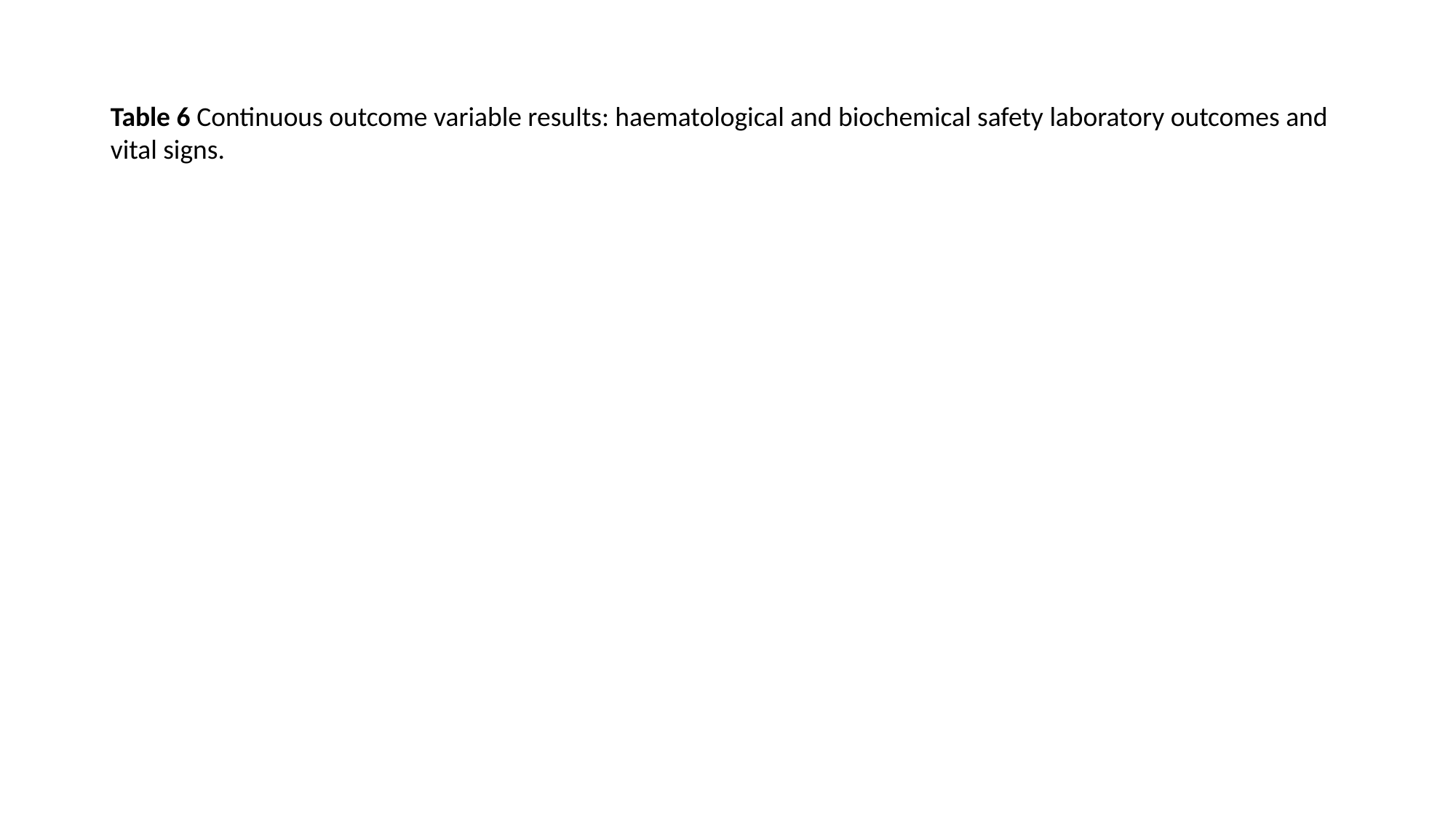

Table 6 Continuous outcome variable results: haematological and biochemical safety laboratory outcomes and vital signs.

### Slide 29
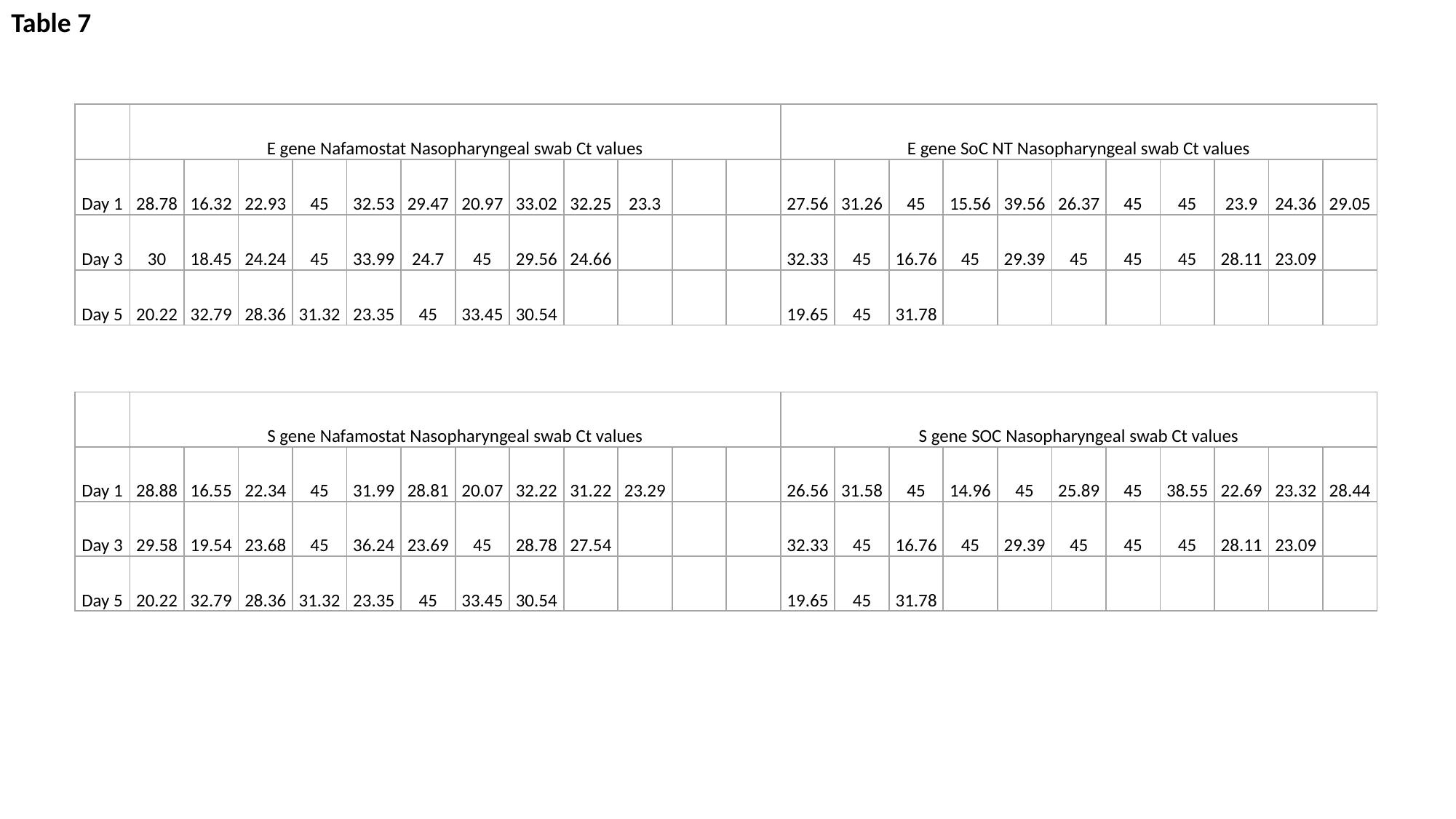

Table 7
| | E gene Nafamostat Nasopharyngeal swab Ct values | | | | | | | | | | | | E gene SoC NT Nasopharyngeal swab Ct values | | | | | | | | | | |
| --- | --- | --- | --- | --- | --- | --- | --- | --- | --- | --- | --- | --- | --- | --- | --- | --- | --- | --- | --- | --- | --- | --- | --- |
| Day 1 | 28.78 | 16.32 | 22.93 | 45 | 32.53 | 29.47 | 20.97 | 33.02 | 32.25 | 23.3 | | | 27.56 | 31.26 | 45 | 15.56 | 39.56 | 26.37 | 45 | 45 | 23.9 | 24.36 | 29.05 |
| Day 3 | 30 | 18.45 | 24.24 | 45 | 33.99 | 24.7 | 45 | 29.56 | 24.66 | | | | 32.33 | 45 | 16.76 | 45 | 29.39 | 45 | 45 | 45 | 28.11 | 23.09 | |
| Day 5 | 20.22 | 32.79 | 28.36 | 31.32 | 23.35 | 45 | 33.45 | 30.54 | | | | | 19.65 | 45 | 31.78 | | | | | | | | |
| | S gene Nafamostat Nasopharyngeal swab Ct values | | | | | | | | | | | | S gene SOC Nasopharyngeal swab Ct values | | | | | | | | | | |
| --- | --- | --- | --- | --- | --- | --- | --- | --- | --- | --- | --- | --- | --- | --- | --- | --- | --- | --- | --- | --- | --- | --- | --- |
| Day 1 | 28.88 | 16.55 | 22.34 | 45 | 31.99 | 28.81 | 20.07 | 32.22 | 31.22 | 23.29 | | | 26.56 | 31.58 | 45 | 14.96 | 45 | 25.89 | 45 | 38.55 | 22.69 | 23.32 | 28.44 |
| Day 3 | 29.58 | 19.54 | 23.68 | 45 | 36.24 | 23.69 | 45 | 28.78 | 27.54 | | | | 32.33 | 45 | 16.76 | 45 | 29.39 | 45 | 45 | 45 | 28.11 | 23.09 | |
| Day 5 | 20.22 | 32.79 | 28.36 | 31.32 | 23.35 | 45 | 33.45 | 30.54 | | | | | 19.65 | 45 | 31.78 | | | | | | | | |

### Slide 30
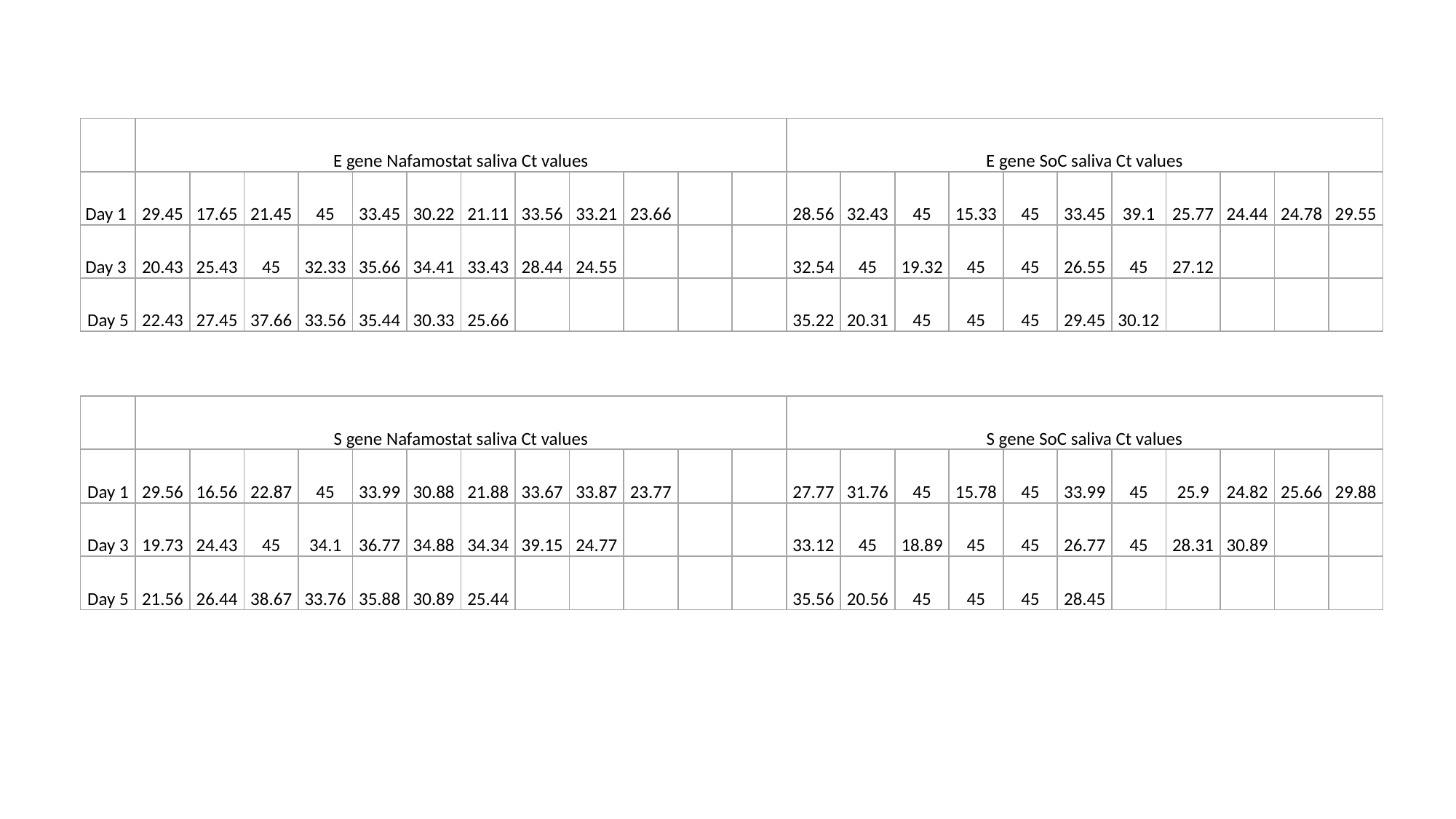

| | E gene Nafamostat saliva Ct values | | | | | | | | | | | | E gene SoC saliva Ct values | | | | | | | | | | |
| --- | --- | --- | --- | --- | --- | --- | --- | --- | --- | --- | --- | --- | --- | --- | --- | --- | --- | --- | --- | --- | --- | --- | --- |
| Day 1 | 29.45 | 17.65 | 21.45 | 45 | 33.45 | 30.22 | 21.11 | 33.56 | 33.21 | 23.66 | | | 28.56 | 32.43 | 45 | 15.33 | 45 | 33.45 | 39.1 | 25.77 | 24.44 | 24.78 | 29.55 |
| Day 3 | 20.43 | 25.43 | 45 | 32.33 | 35.66 | 34.41 | 33.43 | 28.44 | 24.55 | | | | 32.54 | 45 | 19.32 | 45 | 45 | 26.55 | 45 | 27.12 | | | |
| Day 5 | 22.43 | 27.45 | 37.66 | 33.56 | 35.44 | 30.33 | 25.66 | | | | | | 35.22 | 20.31 | 45 | 45 | 45 | 29.45 | 30.12 | | | | |
| | S gene Nafamostat saliva Ct values | | | | | | | | | | | | S gene SoC saliva Ct values | | | | | | | | | | |
| --- | --- | --- | --- | --- | --- | --- | --- | --- | --- | --- | --- | --- | --- | --- | --- | --- | --- | --- | --- | --- | --- | --- | --- |
| Day 1 | 29.56 | 16.56 | 22.87 | 45 | 33.99 | 30.88 | 21.88 | 33.67 | 33.87 | 23.77 | | | 27.77 | 31.76 | 45 | 15.78 | 45 | 33.99 | 45 | 25.9 | 24.82 | 25.66 | 29.88 |
| Day 3 | 19.73 | 24.43 | 45 | 34.1 | 36.77 | 34.88 | 34.34 | 39.15 | 24.77 | | | | 33.12 | 45 | 18.89 | 45 | 45 | 26.77 | 45 | 28.31 | 30.89 | | |
| Day 5 | 21.56 | 26.44 | 38.67 | 33.76 | 35.88 | 30.89 | 25.44 | | | | | | 35.56 | 20.56 | 45 | 45 | 45 | 28.45 | | | | | |

### Slide 31
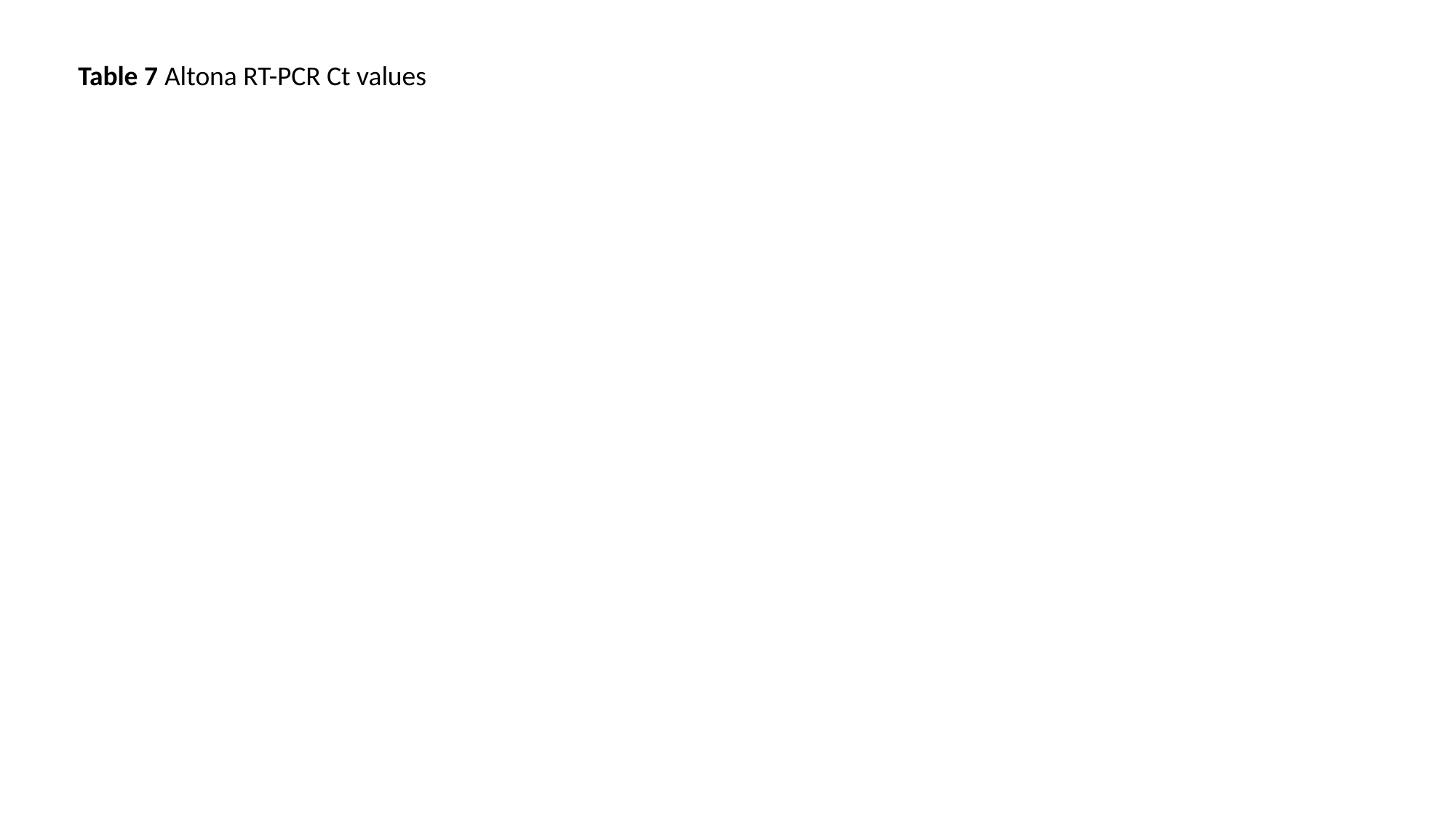

Table 7 Altona RT-PCR Ct values

### Slide 32
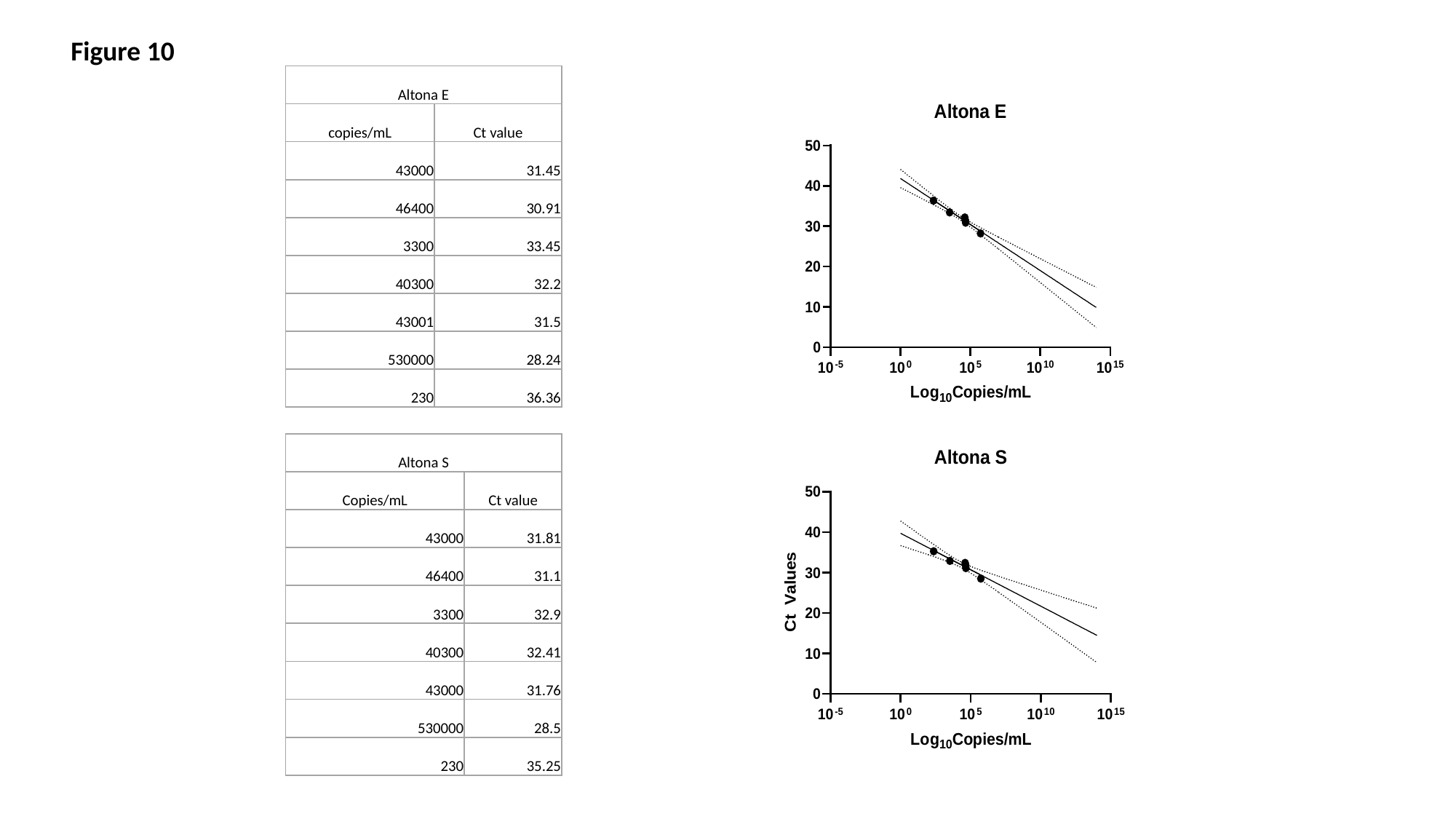

Figure 10
| Altona E | |
| --- | --- |
| copies/mL | Ct value |
| 43000 | 31.45 |
| 46400 | 30.91 |
| 3300 | 33.45 |
| 40300 | 32.2 |
| 43001 | 31.5 |
| 530000 | 28.24 |
| 230 | 36.36 |
| Altona S | |
| --- | --- |
| Copies/mL | Ct value |
| 43000 | 31.81 |
| 46400 | 31.1 |
| 3300 | 32.9 |
| 40300 | 32.41 |
| 43000 | 31.76 |
| 530000 | 28.5 |
| 230 | 35.25 |

### Slide 33
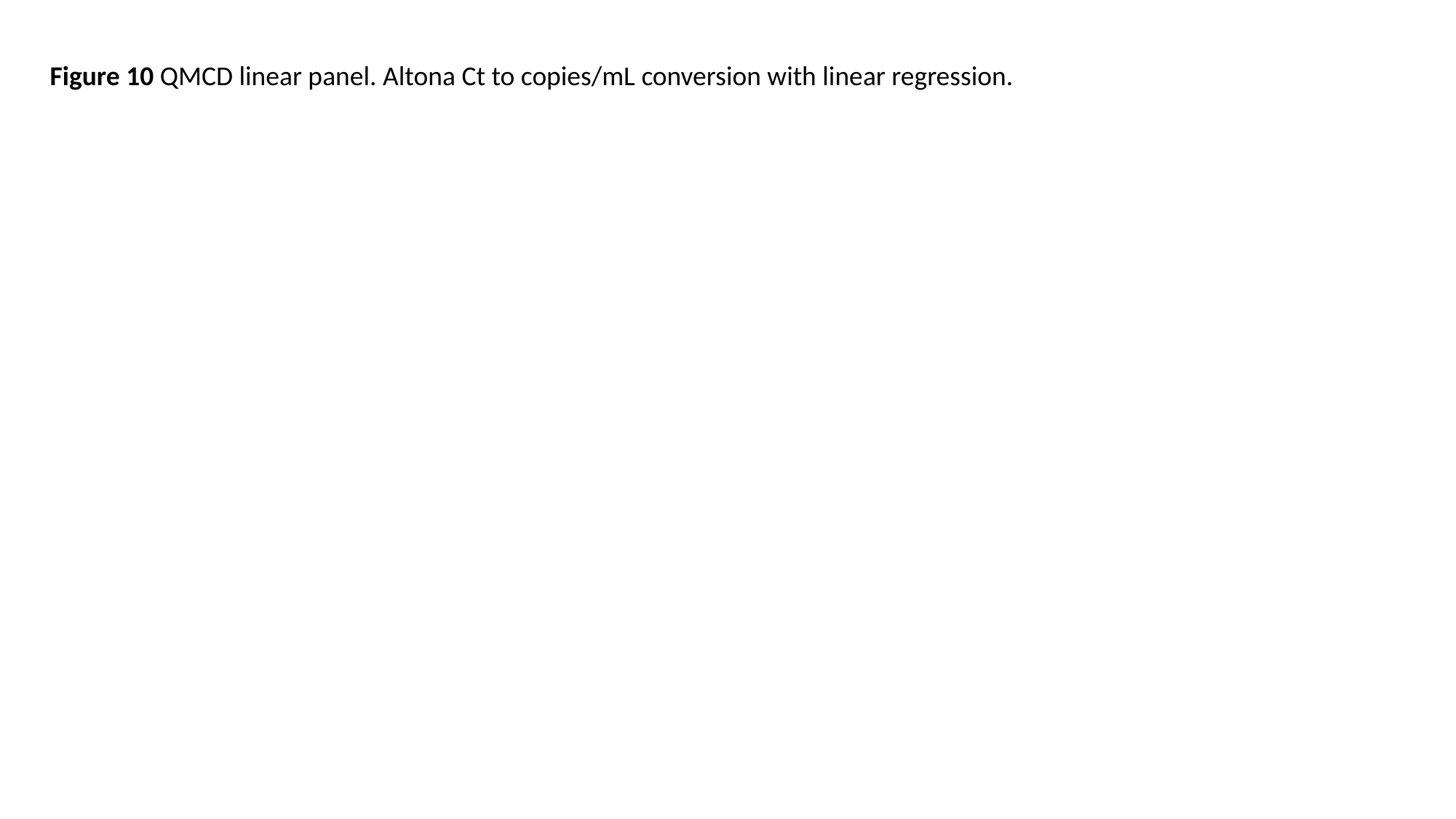

Figure 10 QMCD linear panel. Altona Ct to copies/mL conversion with linear regression.
